# Integrative pan-cancer genomic and transcriptomic analyses of refractory metastatic cancer

**DOI:** 10.1101/2022.11.08.22282064

**Authors:** Yoann Pradat, Julien Viot, Konstantin Gunbin, Andrey Yurchenko, Luigi Cerbone, Marc Deloger, Guillaume Grisay, Loic Verlingue, Véronique Scott, Ismael Padioleau, Leonardo Panunzi, Stefan Michiels, Antoine Hollebecque, Gérôme Jules-Clément, Laura Mezquita, Antoine Lainé, Yohann Loriot, Benjamin Besse, Luc Friboulet, Fabrice André, Paul-Henry Cournède, Daniel Gautheret, Sergey Nikolaev

## Abstract

Metastatic relapse after treatment is the leading cause of cancer mortality, and known resistance mechanisms are missing for most treatments administered to patients. To bridge this gap, we analyze a pan-cancer cohort (META-PRISM) of 1,031 refractory metastatic tumors profiled via whole-exome and transcriptome sequencing. META-PRISM tumors, particularly prostate, bladder, and pancreatic types, displayed the most transformed genomes compared to primary untreated tumors. Standard-of-care resistance biomarkers were identified only in lung and colon cancers - 9.3% of META-PRISM tumors, indicating that too few resistance mechanisms have received clinical validation. In contrast, we verified the enrichment of multiple investigational and hypothetical resistance mechanisms in treated compared to non-treated patients, thereby confirming their putative role in treatment resistance. Additionally, we demonstrated that molecular markers improve six-month survival prediction, particularly in patients with advanced breast cancer. Our analysis establishes the utility of the META-PRISM cohort for investigating resistance mechanisms and performing predictive analyses in cancer.

**Statement of significance:** This study highlights the paucity of standard-of-care markers that explain treatment resistances and the promises of investigational and hypothetical markers awaiting further validation. It also demonstrates the utility of molecular profiling in advanced-stage cancers, particularly breast cancer, to improve the survival prediction and assess eligibility to phase I clinical trial.

## Introduction

Primary untreated tumors have been extensively studied through genomic and transcriptomic profiling, yielding valuable insights into the heterogeneity of tumors and demonstrating the utility of molecular profiling for precision oncology ^1^. Recently, several studies further investigated the genomic landscape of pan-cancer^2 3 4 5^ or tumor-type specific ^6 7 8^ metastatic cohorts. Some widely used antineoplastic drugs, such as cisplatin, induce mutations per se in cancer cells. Additionally, the selective pressure imposed by the cancer therapies leads to tumor evolution and the acquisition of resistance ^9 10 11^. Consequently, the genomes of metastatic tumors that underwent several lines of therapy are expected to contain the genomic footprints of molecular evolution ^12^. Patients with heavily pretreated metastatic tumors also often have no other approved therapies and dismal prognosis. However, these patients might benefit from phase I clinical trials aiming to find new treatment options that do not cause severe side effects. An accurate estimation of the expected survival time is vital to determine patients’ eligibility for these clinical trials.

Currently, survival predictions are based on objective risk clinical markers such as lactate dehydrogenase levels (LDH), serum albumin, neutrophil-to-lymphocyte ratio, or the number of metastatic sites ^13 14^. However, these predictions do not consider tumor molecular markers or characteristics of the tumor microenvironment ^15 16^, which also can be associated with the patient’s survival ^17 18 19 20 21 22 23 24 25^.

This work introduces META-PRISM, a pan-cancer cohort of 1,031 tumors that progressed under at least one line of treatment or have no approved therapy options. Somatic variations, germline mutations, and tumor microenvironments were analyzed via whole-exome sequencing (WES) and RNA sequencing (RNAseq). Identified genetic markers were compared to tumor-type matched untreated primary tumors from The Cancer Genome Atlas (TCGA^26^), focusing on functional pathogenic variants associated with resistance, and validated using metastatic tumors from MET500 cohort^3^. We further investigated the utility of genomic and transcriptomic markers to improve the prediction of survival time based on objective clinical variables in the META-PRISM cohort.

## Results

### 1. META-PRISM cohort overview

The META-PRISM cohort is composed of 1,031 adult patients that were biopsied at entry into precision medicine trials (MOSCATO^27^, MATCH-R^28^) and subjected to whole-exome sequencing (571 biopsies-blood pairs) and transcriptome sequencing (947 biopsies; Fig. 1A, Fig. 1B). The clinical history of patients, tumor characteristics, and other clinical parameters derived from blood testing and physiological assessments were tabulated (Supplementary Table S1), as were details about the library preparation and sequencing quality of the analyzed samples (Supplementary Table S2). The median age of the patients was 59.8 years, ranging from 43.1 to 68.9 years per tumor type (Fig. 1C). The cohort included 39 cancer types (Supplementary Table S3), with 5 of them represented by more than 50 tumors: 192 lung adenocarcinomas (LUAD), 98 breast carcinomas (BRCA), 95 prostate adenocarcinomas (PRAD), 76 bladder urothelial carcinomas (BLCA) and 61 pancreatic adenocarcinomas (61 PAAD) (Fig. 1A, 1B). Moreover, 163 tumors were of rare subtypes or unknown primary (Fig. 1B) and included 42 tumors with neuroendocrine differentiation (FS1). We focused our genetic analysis on the tumor types (excluding tumors of unknown origin) that were represented by at least ten sequenced tumors, resulting in ten types for WES (META-PRISM WES, 84.1% of all DNA samples), ten types for WES and RNAseq (META-PRISM WES & RNAseq, 82.3% of all DNA & RNA samples), and 20 for RNAseq (META-PRISM RNAseq, 84.3% of all RNA samples; Supplementary Table S2; Fig. 1B).

**Fig 1.**
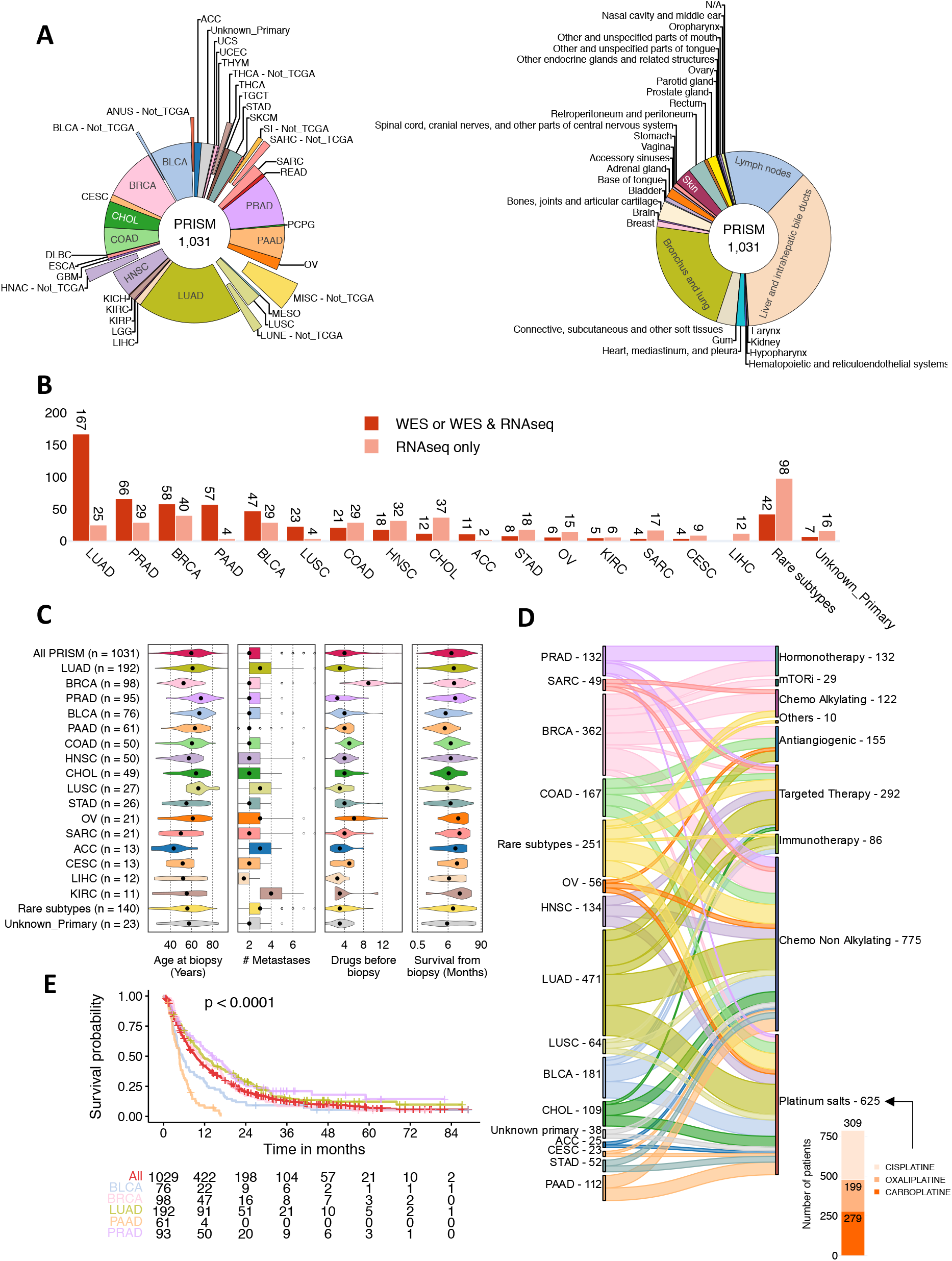
Clinical characteristics, sequencing data, treatment history, and outcomes of the META-PRISM cohort. **A**. Pie chart of the tumor types according to TCGA classification (left) and of the biopsy sites according to ICD-O-3 nomenclature (right). Tumors that could not fit into TCGA classification were classified into tumor types suffixed by “Not_TCGA” and are shown as exploded slices. **B**. Number of tumors per tumor type with either RNAseq only, WES only, or WES and RNAseq. Tumors with WES only or WES and RNAseq were grouped. Only tumor types represented by at least ten samples in META-PRISM WES are shown. **C**. Violin plots describing the age distribution, the number of detected metastatic sites at the time of the biopsy, the number of treatments received before the biopsy, and the survival time from the biopsy date for each tumor type. Only treatments for which resistance was diagnosed before or shortly after the biopsy are listed. Only the survival times of deceased patients are used in the violins (909/1,031). **D**. River plot showing the number of treatment-patient associations per tumor type and drug family. Only relationships involving at least ten tumors are displayed for clarity. Additional details are provided in the side bar plot about the different platinum treatments used. A panel of medical experts established the drug families. **E**. Kaplan-Meier survival curves of the whole META-PRISM cohort and the five most represented tumor types. The p-value was calculated using a log-rank test.

At the time of the biopsy, patients had a median number of two metastases distributed at 35 different sites (Fig. 1C, Supplementary Fig. S2). The most frequently biopsied metastatic sites were liver (37%), lung (22%), and lymph nodes (15%) (Fig. 1A, Supplementary Fig. S2). Before the biopsy, patients had received a median number of four treatments, with BRCA and ovarian carcinomas (OV) being the most pretreated tumors (medians of six and nine treatments, respectively). Most treatments were tumor-type specific such as EGFR inhibitors in LUAD or hormone therapies in BRCA or PRAD. Other therapies, particularly platinum-based drugs, were applied to multiple tumor types (Fig. 1D).

The median survival time from the biopsy date was 7.8 months (n=909 deceased patients out of 1,029) (Fig. 1C) and was highly heterogeneous across tumor types. PAAD, BLCA, carcinomas of unknown primary and lung squamous cell carcinomas (LUSC) had very poor prognosis (<6 months median survival), while cervical squamous cell carcinomas and endocervical adenocarcinomas (CESC), OV, sarcomas (SARC), and kidney renal clear cell carcinomas (KIRC) had longer survival (>12 months; Fig. 1C, 1E).

### 2. Genomic and transcriptomic description of META-PRISM

In order to provide a comparative assessment of the types and extent of genetic variations in META-PRISM, we took advantage of the TCGA cohort representing primary untreated tumors (Supplementary Fig. S3A, S3B). Furthermore, we reanalyzed the previously published MET500 pan-cancer cohort of metastatic tumors to validate discoveries made on META-PRISM (Supplementary Figs. S3C, S3D).

#### a. Variations derived from WES

##### i. Somatic mutations: Tumor mutational burden (TMB) and mutational signatures

WES analysis revealed a significant increase of somatic mutations in META-PRISM vs. TCGA for six of ten studied tumor types. Tumors from the MET500 cohort demonstrated a tumor mutational burden (TMB) similar to that of META-PRISM tumors. We observed the most significant increase of TMB in low-burden tumor types BRCA (2.3x fold in META-PRISM, 3.0x fold in MET500), PRAD (2.1x fold in META-PRISM, 2.2x in MET500) and PAAD (1.7x fold in META-PRISM); while no such difference was observed in high-burden tumor types, namely BLCA, LUAD, and LUSC (Fig. 2A).

**Fig 2.**
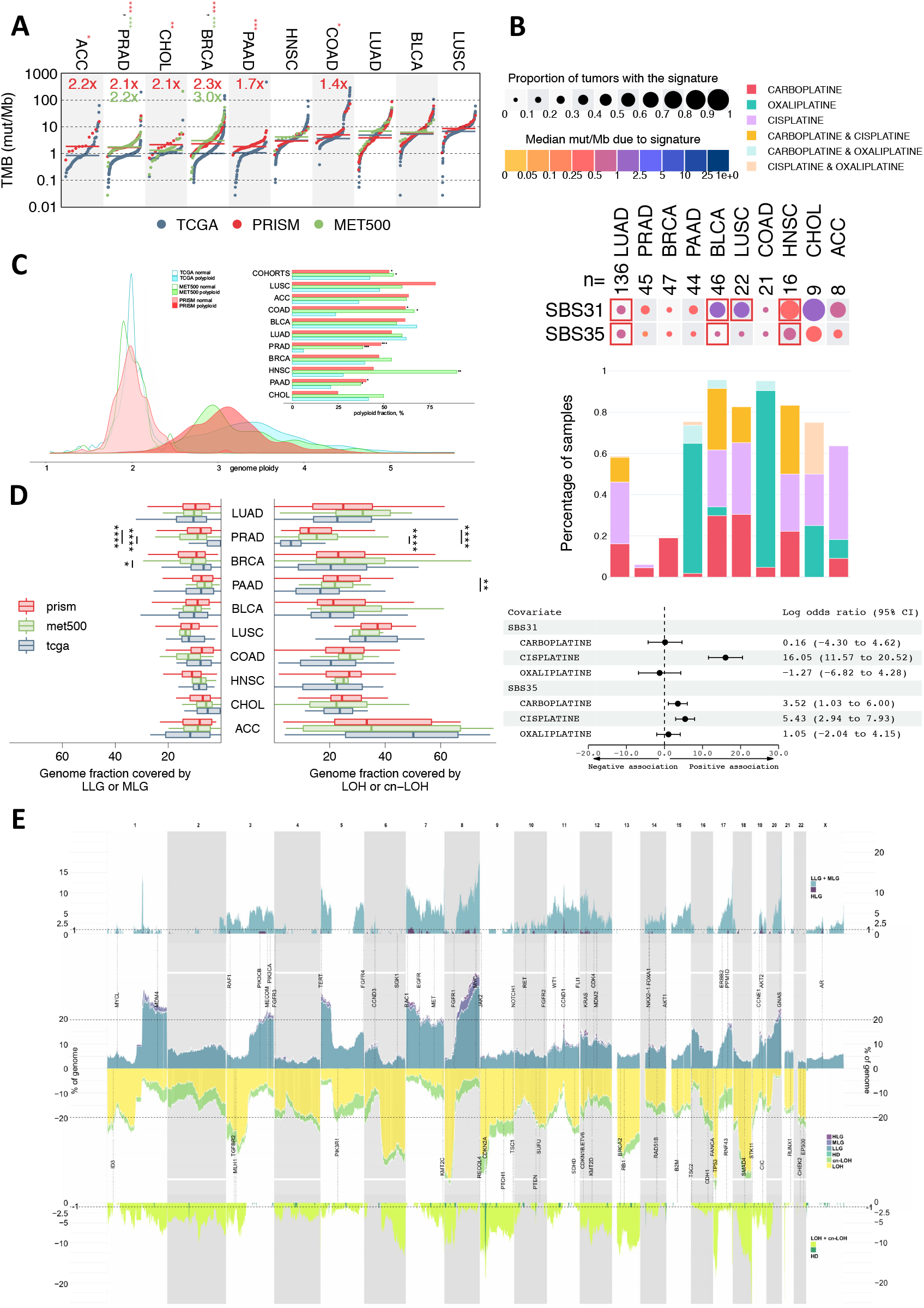
The genomic landscape of META-PRISM tumors. **A.** Distribution of the mutational burden for each cohort and each tumor type of the META-PRISM WES subcohort (red), MET500 (green), and TCGA (blue) cohorts. The fold changes and the p-values from Mann-Whitney U tests represent comparisons between mutational burdens of META-PRISM and TCGA (red), and MET500 and TCGA (green; * p < 0.05, ** p < 0.01, *** p < 0.001, Supplementary Methods). **B**. The upper panel depicts the detection level of mutational signatures (rows) in tumor types (columns). Colors indicate the median number of mutations contributed by the signature in samples harboring the signature. Red frames indicate significant Mann-Whitney U tests comparing META-PRISM with TCGA. The middle panel provides information about the platinum drugs used in each tumor type. The lower panel shows the log odds ratio from two logistic regression models predicting the presence or absence of SBS31 and SBS35 in all WES samples from META-PRISM with at least 50 somatic mutations (Supplementary Methods). **C**. Density plot depicting the distribution of the estimated average ploidy in META-PRISM, MET500, and TCGA tumors for samples with and without WGD. The bar plot shows the proportion of polyploid tumors in the full cohorts and per tumor type. **D**. Double box plot describing the genome fraction covered by gains (low and middle level; left) and losses (loss of heterozygosity – LOH – and copy neutral – cn-LOH; right) per tumor type. Comparisons of META-PRISM vs. TCGA and MET500 vs. TCGA were performed using Mann-Whitney U tests (* p < 0.05, ** p < 0.01, *** p < 0.001, **** p < 0.0001). **E**. The middle panel describes the fraction of tumors harboring different types of copy gains and losses across the genome (Supplementary Methods). The upper and lower panels show the excess of copy gains and losses in META-PRISM compared to TCGA. SCNAs were classified into three types of copy gains: low, middle, and high-level, and three types of copy losses: LOH, cn-LOH, and homozygous deletions (HD). The vertical dotted lines align to the loci where selected oncogenes (above) and tumor suppressors (below) are located. Only tumor types from the META-PRISM WES subcohort (ten tumor types) are represented in this figure. All p-values were adjusted for multiple testing using the Benjamini-Hochberg procedure. P-values in panel **B.** were adjusted by considering all the tests on the complete list of deconvoluted signatures (Supplementary Fig. S4 and Methods).

Deconvolution of mutational signatures revealed similar signature compositions between metastatic META-PRISM, MET500, and primary TCGA cancers in all studied tumor types (Supplementary Fig. S4 and Methods). However, a notable and consistent difference in META-PRISM vs. TCGA across tumor types was the presence of signatures associated with platinum treatments (SBS31 and SBS35), reflecting that the majority of META-PRISM tumors (691 of 1,011 with known drug history) were pretreated with platinum compound therapies. Signatures SBS31 and SBS35 were detected in more than 50% of tumors in six META-PRISM tumor types (LUAD, BLCA, LUSC, HNSC, cholangiocarcinomas – CHOL –, and adrenocortical carcinomas – ACC) and contributed significantly more mutations compared to TCGA in four of them (Fig. 2B). Tumor types varied in frequency and types of received platinum drugs. For example, BRCA and PRAD rarely received platinum treatments and concordantly demonstrated very low platinum signature. We then investigated the association of SBS31 and SBS35 with three main platinum drugs in our cohort: cisplatin, carboplatin, and oxaliplatin. Among these drugs, cisplatin had the strongest association with SBS31 and had an association with SBS35 that was comparable to carboplatin, as revealed by logistic regression (Fig. 2B). More specifically, among tumors harboring at least 50 somatic mutations (Supplementary Methods) and treated with cisplatin and no other platinum compound (n=95), 40% harbored SBS31 and 17% SBS35, while in patients treated only with carboplatin (n=80), 17% had a detectable activity of SBS35. In contrast, these two signatures were rarely detectable in tumor types (COAD and PAAD) predominantly treated by oxaliplatin (Fig. 2B).

##### ii. Somatic Copy Number Alterations (SCNA)

Whole-genome duplication (WGD) is a frequent event in cancer involving doubling the chromosome complement. We detected WGDs in 52.2% of META-PRISM WES tumors, which was comparable to MET500 (51.5%) but significantly higher than in TCGA (42.5%; p=0.01, U-test; see Methods) (Fig. 2C). The fraction of WGDs varied between tumor types ranging from 25.0% in CHOL to 78.3% in LUSC in META-PRISM. The most striking increase of WGD events in metastatic cancers compared to TCGA was observed in PRAD (META-PRISM 48.5%, 7.5x fold increase, p < 0.001; MET500 38.6%, 6x fold increase, p < 0.001; Mann-Whitney U tests) (Fig. 2C).

We next investigated the landscape of somatic copy number alterations (SCNA) in META-PRISM WES tumors without WGD. Using FACETS ^29^, we categorized SCNAs in META-PRISM, MET500, and TCGA as copy gains and losses. Copy gains were subdivided into either low (LLG), middle (MLG), or high-level (HLG) gains. Copy losses included loss of heterozygosity (LOH), copy-neutral LOH (cn-LOH), and focal homozygous deletions (HD). LLGs, MLGs, LOH, and cn-LOH often spanned large regions or covered full chromosome arms, while HD and HLGs were almost always focal. The fraction of the genome covered by low- to medium-level gains or LOH was similar across most cancer types except for PRAD tumors, which had an increase in both types of copy-number changes in metastatic tumors compared to TCGA (Fig. 2D). The SCNA landscape of META-PRISM was similar to that of TCGA for both the full tumor-type matched dataset (Fig. 2E) and specific tumor types (Supplementary Figs. S5-S9). The frequency of the majority of large SCNAs did not differ significantly.

In META-PRISM WES tumors without WGD, an average of 7.3% and 19.5% of the genome was covered by copy gains and copy losses, respectively. No significant increase in this type of instability was observed in most studied tumor types except for PRAD, which demonstrated a dramatic increase in metastatic tumors compared to primary tumors, and for PAAD, where the increase was limited to copy losses (Fig. 2D, Supplementary Figs. S8, S9). Three chromosome arm gains (5p, 8q, 7p) and six losses (6q, 8p, 9p, 13q, 17p, 18q) were observed in more than 20% of the non-WGD META-PRISM cohort. However, their frequency was not significantly different from TCGA (Supplementary Fig. S10). The majority of these chromosome regions enriched with gains and losses events were shared between several tumor types, while some chromosome regions were tumor type specific. For instance, +16p, -16q, -22q in BRCA, +20p, +20q, -9q, -11p in BLCA (Supplementary Fig. S10). META-PRISM tumors with WGD had a higher fraction of chromosome arm events but a lower level of homozygous deletions than tumors without WGD (Supplementary Fig. S11A). In contrast, the number of driver mutations in oncogenes or tumor suppressor genes was not influenced by WGD events (Supplementary Fig. S11B).

High-level amplifications and homozygous deletions were rare in the META-PRISM WES tumors, spanning an average of 0.20% and 0.04% of the genome, respectively. However, highly amplified and homozygously deleted genes were detected on average 35 and 12 times per tumor. The most frequent of these events included amplification of *CCND1* (8.0 %), *AR* (3.5%), 19q13 genes (2.9%), *EGFR* (2.7%), *KRAS* (2.7%), *MYC* (2.7%), and losses of *CDKN2A* (13.1%), *FAM106A / LGALS9C* (4.8%), Killer Ig-like receptors genes (4.6%), *PTEN* (4.0%), GSTT1 (3.8%), *RHD / RSRP1* (3.3%) and were enriched in META-PRISM (and in MET500) compared to TCGA (Supplementary Fig. S11C).

##### iii. Mutations and SCNAs in driver genes

The discovery of significantly mutated genes with the Mutpanning tool ^30^ confirmed previously reported cancer drivers (Supplementary Table S4). We next selected a list of 360 cancer genes by intersecting COSMIC census ‘Tier 1’ and ‘Tier 2’ (v92) and OncoKB-annotated genes ^31^ (OncoKB 07/2021; Supplementary Table S5) and created a catalog of driver mutations and SCNAs (point mutations, small indels, amplifications, and deletions) using OncoKB annotations on these genes only (Supplementary Tables S6, S7). These types of driver events were observed in 95% of the META-PRISM WES tumors. The most frequently altered driver genes in META-PRISM were *TP53* (55% of samples), *KRAS* (25%), *CDKN2A* (17%), and *EGFR* (14%) (Fig. 3A). On the whole cohort level, 15 oncogenes and two tumor suppressor genes were significantly enriched compared to TCGA; 73% and 50% of those genes, respectively, were also enriched in MET500. Some driver genes were significantly enriched in specific tumor types, for example, *EGFR* and *CTNNB1* in LUAD; *TP53*, *AR*, *PTEN*, *FOXA1*, *RB1*, and *MYC* in PRAD; *ESR1* and *CCND1* in BRCA; *KRAS* in PAAD; *FGFR3*, *CCND1, MDM2* and *UBR5* in BLCA (Fig. 3A, Fisher-Boschloo tests, Benjamini-Hochberg correction; Supplementary Methods). The number of WES-derived driver events (mutations and SCNAs) was significantly higher in metastatic tumors as compared to primary tumors at the cohort level (medians of four and three in META-PRISM and MET500 vs. two in TCGA, p < 0.0001) and for five of ten tumor types included in the META-PRISM WES subcohort (Fig. 3B).

**Fig 3.**
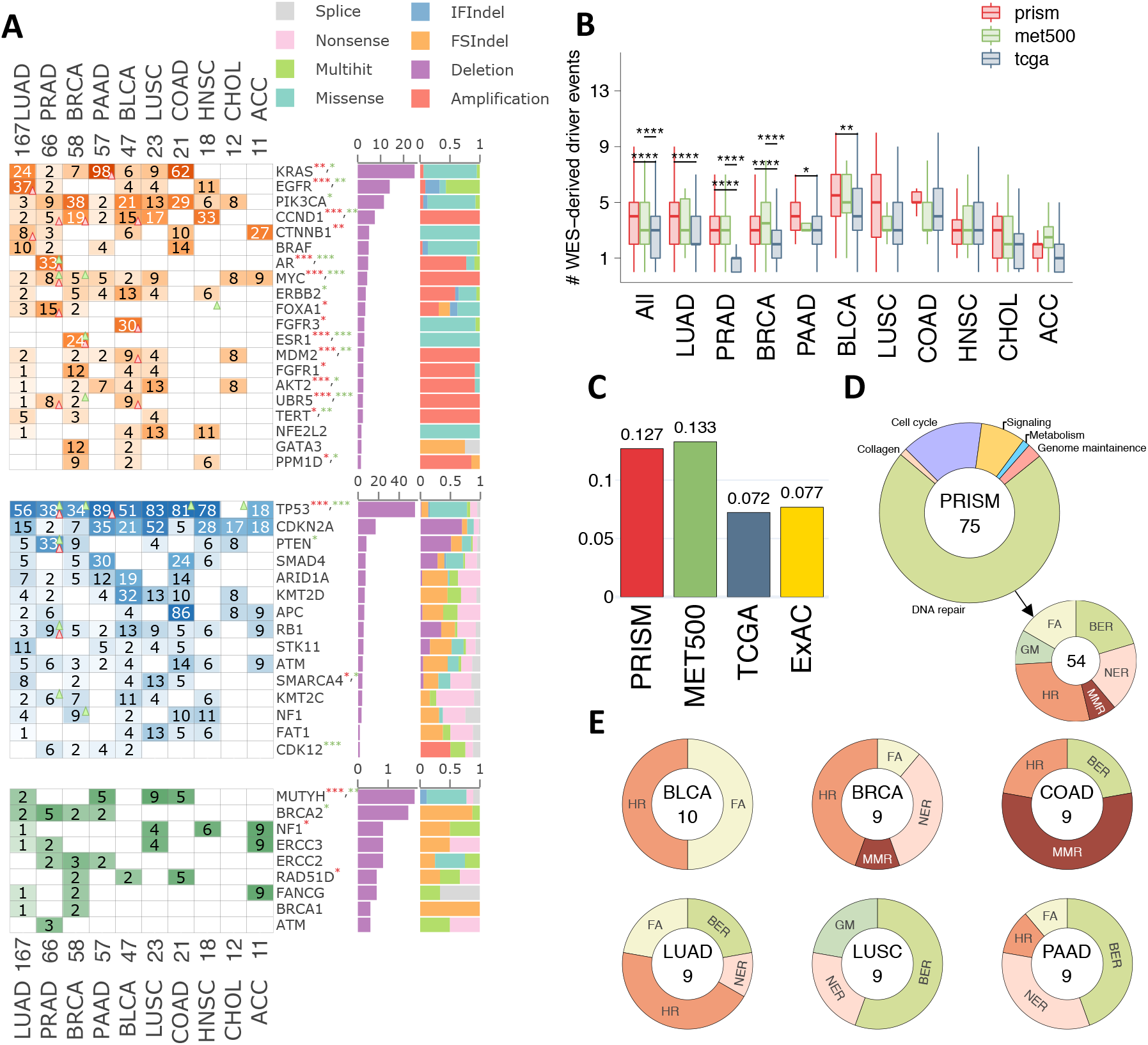
The landscape of cancer-associated somatic and germline mutations and SCNAs in META-PRISM tumors. **A**. Heatmaps depicting the percentage of tumors harboring driver events (point mutations, small indels, SCNAs) in top oncogenes (upper panel), top tumor suppressor genes (middle panel), and top cancer-predisposing genes (lower panel). Triangles’ orientations (increase – triangle points up, decrease – points down) and colors (red for META-PRISM vs. TCGA, green for MET500 vs. TCGA) highlight significant changes in frequency. Similarly, stars next to the gene names represent significant changes at the cohort level using the same color code as for triangles (* p < 0.05, ** p < 0.01, *** p < 0.001). The absolute bar plots show the percentage of tumors in META-PRISM harboring the alteration, while the relative bar plots show the breakdown of these alterations into different categories. Adjusted p-values per tumor type are from Fischer-Boschloo tests, whereas p-values across the cohort are from Cochran-Mantel-Haenszel tests. **B.** Box plot of the number of driver events in META-PRISM, MET500, and TCGA. Adjusted p-values shown in the box plot are derived from Mann-Whitney U tests. **C.** Bar plot representing the incidence of cancer-risk germline variants in META-PRISM, MET500, TCGA cancer patients, and in ExAC European (non-Finnish) population. Stratified Cochran–Mantel–Haenszel test is used to account for tumor type compositions in the cohorts except for comparison with ExAC where the standard Fisher test is used. **D.** Pie charts representing the distribution of cancer-risk variants by pathways in META-PRISM tumors. **E.** Pie charts representing the distribution of cancer-risk variants in DNA repair pathways for each of the six tumor types harboring the most germline events. FA: Fanconi anemia, GM: genome maintenance, BER: base excision repair, NER: nucleotide excision repair, MMR: mismatch repair, HR: homologous repair. All p-values are adjusted for multiple testing using the Benjamini-Hochberg procedure. Only tumor types represented in the META-PRISM WES subcohort are shown in this figure.

Major tumor suppressor genes frequently underwent biallelic inactivation. Such inactivations were observed in 23% of *TP53*-hit tumors, 73% for *CDKN2A*, 63% for *PTEN*, 31% for *SMAD4*, 23% for *ARID1A*, 34% for *APC*, 50% for *RB1*. However, the predominant mechanisms of biallelic inactivation differed from one gene to another: in *TP53*, it was mutation followed by LOH; in *CDKN2A* and *PTEN*, it was homozygous deletions; other genes demonstrated a combination of mechanisms (Supplementary Fig. S12A). Few oncogenes also underwent multi-hit events, most notably *EGFR* (70%), *KRAS* (17%), and *PIK3CA* (23%). Multi-hit events in *EGFR* in META-PRISM were significantly more frequent than in TCGA, likely reflecting the effect of EGFR inhibitors in LUAD (Supplementary Fig. S12B).

##### iv. Germline cancer risk variants

WES of germline DNA from 571 META-PRISM patients was used to identify pathogenic cancer-predisposing variants. We focused on the germline variants annotated as pathogenic or likely pathogenic in the ClinVar database or protein-disrupting and residing in genes strongly associated with cancer predisposition^32^ (Huang et al. Cell 2018; Supplementary Methods). We identified 75 patients in META-PRISM (13.1%) harboring at least one such variant. The fraction of patients with cancer-predisposing variants was similar to that in MET500 patients and was 1.75-times higher than in TCGA (8 cancer types, p=0.0012, Fisher’s exact test) or in ExAC Non-Finish Europeans (OR=1.75, p=0.0002; Fig. 3C). 73% of variants were in DNA repair pathway genes (Fig. 3D). The most frequent genes with cancer-predisposing variants in META-PRISM patients were: *MUTYH* (1.9%), *BRCA2* (1.8%), *NF1* (0.9%), *ERCC2* (0.9%), *ERCC3* (0.9%), *RAD51D* (0.7%) and *FANCG* (0.7%) (Fig. 3A). *MUTYH*, *NF1,* and *RAD51D* were also significantly enriched compared to TCGA. We detected an increase of germline cancer-risk variants in most cancer types in META-PRISM. However, it reached significance only for PRAD (p=0.03) and LUSC (p=0.004) cancer types. Mutations in the homologous recombination pathway were the most frequent in BRCA, PRAD, LUAD, and BLCA; base excision repair pathway – in PAAD and LUSC; mismatch repair pathway – in COAD (Fig. 3E). 37% of genes with germline cancer-risk variants harbored a somatic second-hit event, including somatic mutations in 9% and LOH resulting in the retention of the pathogenic variant in 27%.

#### b. Variations derived from RNAseq

##### i. Tumor Microenvironment (TME)

The tumor microenvironment (TME) plays a significant role in clinical outcomes and response to therapy ^17 18 33 34^. Tumors from META-PRISM, MET500, and TCGA were classified into four tumor microenvironment subtypes – immune-enriched, fibrotic (IE/F); immune-enriched, non-fibrotic (IE); fibrotic (F); and immune-depleted (D) – using functional gene expression signatures scores as described in Bagaev et al. ^16^ (Supplementary Methods). No consistent difference in the distribution of TMEs across tumor types was observed when comparing META-PRISM to TCGA. However, the TME subtypes in some individual tumor types showed striking variation between the cohorts. For instance, immunosuppressive subtypes D and F were significantly increased in PRAD and BLCA, respectively, in META-PRISM compared to TCGA (63% vs. 41%, p < 0.001; 33% vs. 17%, p=0.005; p-values are not adjusted due to the lack of independence Fig. 4A). Enrichment of the D subtype in PRAD was also significant in MET500 vs. TCGA (78% vs. 41%, p < 0.001).

**Fig 4.**
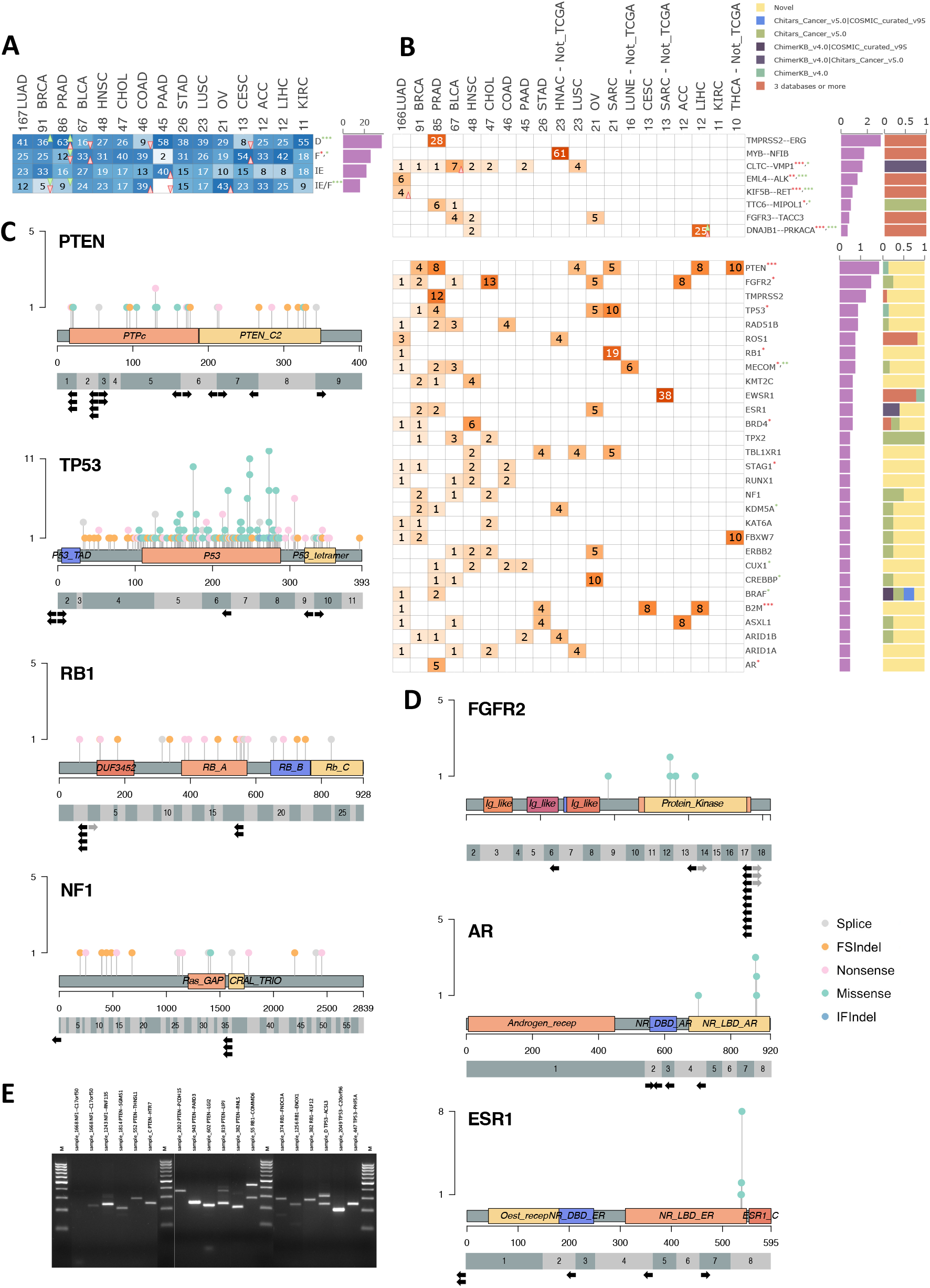
The landscape of tumor micro-environments and cancer-associated gene fusions. **A.** Heatmap depicting the percentage of tumors classified in each TME subtype for each tumor type of the META-PRISM RNAseq subcohort (non-TCGA tumor types and SARC were excluded, Supplementary Methods). **B.** Heatmaps depicting the percentage of tumors harboring gene fusions for known cancer gene fusions (upper panel) or known cancer drivers (lower panel). Only known oncogenic gene fusions seen in at least four samples among META-PRISM RNAseq tumors are shown in the upper panel. The lower panel considers all known oncogenic gene fusions implicating drivers except for the fusions shown in upper plot which are excluded. Likewise, only drivers involved in fusions at least four samples among META-PRISM RNAseq tumors are shown. **C.** Fusions involving tumor suppressor genes *PTEN*, *TP53*, *RB1*, and *NF1*. The top part of each panel indicates the protein domains (amino acid numbering) and locations and recurrence of driver somatic mutations categorized into splice site, frameshift, inframe, nonsense, and missense mutations. The bottom part shows fusion event breakpoints on the exonic structure. Only coding exons are shown. Black arrows indicate fusions breakpoints with the driver gene transcript located 5’ (left arrow) or 3’ (right arrow) of the breakpoint. Grey arrows indicate secondary fusions for which a principal fusion (with higher coverage) is found in the same patient and involves the same gene. **D.** Identical to **C.** considering oncogenes *FGFR2*, *AR*, and *ESR1*. **E.** RT-PCR validation of 23 fusions in *TP53*, *RB1*, *PTEN*, *NF1*, *AR* (Supplementary Methods). **A-B.** Triangles’ orientations (increase – triangle points up, decrease – points down) and colors (red for META-PRISM vs. TCGA, green for MET500 vs.TCGA) highlight significant changes in subtype frequency. Similarly, stars next to the gene names represent significant changes at the cohort level using the same color code as for triangles (* p < 0.05, ** p < 0.01, *** p < 0.001). P-values per tumor type are from Fischer-Boschloo tests, while p-values across the cohort are from Cochran-Mantel-Haenszel tests. **A.** p-values were not corrected for multiple testing due to the lack of independence between the tests performed within each tumor type.

##### ii. Known and novel driver gene fusions

We next investigated gene fusions in the 796 META-PRISM RNAseq tumors that were successfully processed by the fusion pipeline. The calling of fusions was restricted to known oncogenic fusions and fusions involving a cancer driver as only these two types of fusions are most likely relevant to cancer and could reliably be detected in all three cohorts (Supplementary Methods).

A total of 437 known oncogenic gene fusions (Chimer KB v4.0 ^35^; Chitars v5.0 ^36^ ; COSMIC v95; TIC v3.3 ^37^) were identified in META-PRISM RNAseq tumors (34%, Supplementary Table S8). As previously described, well-known oncogenic gene fusions were often tumor-type specific – *TMPRSS2*-*ERG* 28% in PRAD, *EML4*-*ALK* 6% in LUAD, *DNAJB1*-*PRKACA* 25% in LIHC, *MYB*-*NFIB* 61% in head and neck adenoid cystic carcinomas (HNAC, rare subtype) – and some were significantly enriched in META-PRISM vs. TCGA (Fig. 4B). Additionally to known oncogenic fusions, we identified 330 fusions involving cancer driver genes and promiscuous partners (29% of META-PRISM RNAseq tumors; Supplementary Table S8).

Among the most recurrently fused driver genes, we identified tumor suppressors *PTEN* (1.9%), *TP53* (0.9%), *RB1* (0.8%), and *NF1* (0.5%). In these genes, 67% of fusion breakpoints were recurrent (Fig. 4C). *PTEN* fusions were observed in several tumor types in META-PRISM, but they were most prevalent in PRAD (Fig. 4B). The oncogene most frequently involved in fusions across different tumor types was *FGFR2* (1.5%). Interestingly, *ESR1* and *AR*, both known to be important factors of hormone therapy resistance in BRCA and PRAD, were involved in fusions in 2.2% and 4.7% of the respective tumor types in META-PRISM (Fig. 4B, 4D). 16 out of 18 tested fusions (89%) were validated through RT-PCR and Sanger sequencing (Fig. 4E, Supplementary Table S9).

### 3. Markers of resistance to treatment and cancer aggressiveness

#### a. Genomic variations relevant to treatment

##### i. Actionable variants: markers of resistance and sensitivity to treatments

Biomarkers of resistance and response to anticancer therapies were annotated in META-PRISM, MET500, and TCGA by mapping somatic events to the OncoKB and CIViC databases ^31 38^ (Supplementary Tables S6-S8). ESCAT guidelines ^39^ were used to classify biomarkers as Tier 1, Tier 2, or Tier 3. Tier 1 is standard-of-care (SOC) markers that are used in routine clinical practice for treatment indications; Tier 2 is investigational markers supported by clinical trials but for which additional data are needed; and Tier 3 is hypothetical targets supported by scarce or off-label data.

We observed SOC resistance and sensitivity biomarkers in 9.3% and 46.6% of META-PRISM WES & RNAseq tumors, respectively (Fig. 5A-B), while biomarkers of any level were found in 73.4% and 87.2% of these tumors, respectively. Tier 1 resistance biomarkers were detected in only three tumor types: LUAD (*EGFR* 17%), LUSC (*EGFR* 5%), and COAD (*KRAS* 65%, *NRAS* 6%); while investigational Tier 2 and hypothetical Tier 3 biomarkers were rather frequent in the majority of tumor types (Fig. 5A). META-PRISM WES & RNAseq tumors harbored significantly more resistance biomarkers of all three tiers compared to TCGA: Tier 1, common odds ratio (cOR) 7.3 (CI 95% [3.6, 14.7], CMH test stratified by tumor type); Tier 2, cOR 1.9 (CI 95% [1.5, 2.5]); and Tier 3, cOR 2.2 (CI 95% [1.7, 2.8]). The increase was also replicated in MET500 for all three tiers (Tier1 cOR 4.6 – CI 95% [1.2, 16.8] –, Tier2 cOR 2.0 – CI 95% [1.4, 2.9] –, Tier3 cOR 3.5 – CI 95% [2.6, 4.6]).

**Fig 5.**
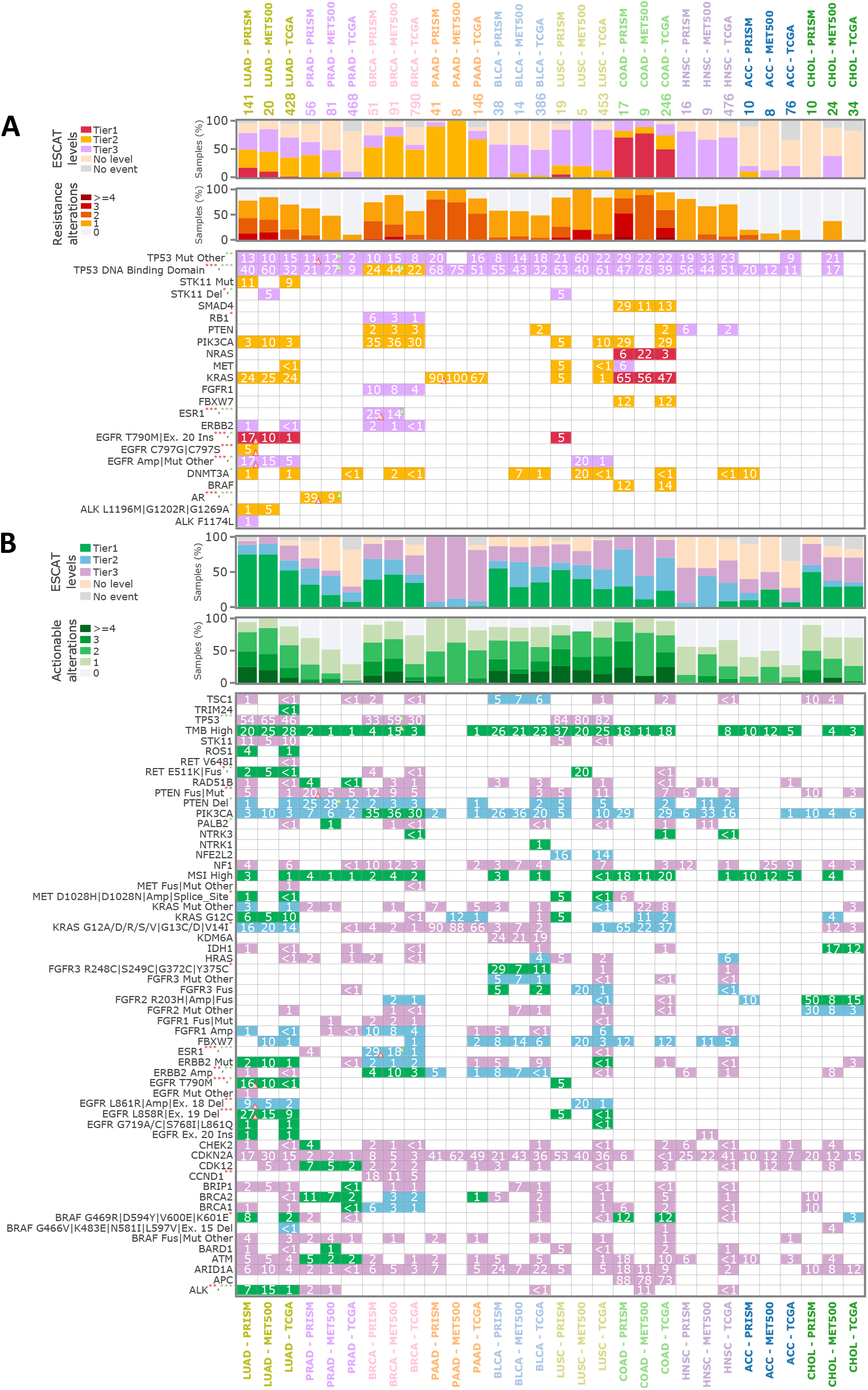
Known genetic markers of treatment resistance and sensitivity in META-PRISM. **A.** Set of panels showing the prevalence of known genetic markers of resistance to treatment in META-PRISM, MET500, and TCGA by tumor type. Upper panel – fractions of tumors harboring resistance markers split by tier (only the best tier is shown for each tumor). Middle panel – fractions of tumors with multiple resistance markers. Lower panel – heatmap showing the most frequent resistance-associated variants. Triangles’ orientations (increase – triangle points up, decrease – points down) and colors (red for META-PRISM vs. TCGA, green for MET500 vs. TCGA) highlight significant changes in prevalence. Similarly, stars next to the gene alterations represent significant changes at the cohort level using the same color code as for triangles (* p < 0.05, ** p < 0.01, *** p < 0.001). P-values per tumor type are from Fischer-Boschloo tests, and p-values across the cohort are from Cochran-Mantel-Haenszel tests. All p-values were adjusted for multiple testing using the Benjamini-Hochberg procedure. **B.** Identical to **A.** but considering markers of sensitivity to treatments.

Some common sensitivity biomarkers were frequent in META-PRISM and often enriched vs. TCGA, most notably: *EGFR* L858R/Exon 19 del (first- and second-generation *EGFR* inhibitors), *EGFR* T790M (third-generation *EGFR* inhibitors), and *ALK* oncogenic mutations or fusions (ALK inhibitors) in LUAD; *PTEN* fusions or loss-of-function mutations (mTOR inhibitors) in PRAD; *FGFR3* p.R248C, p.S249C, p.G372C, p.Y375C mutations (*FGFR* inhibitors) in BLCA. On top of that, we detected 13% of hypermutated tumors (>10M/Mb) and 3% of microsatellite-unstable tumors in META-PRISM, which are indications for treating patients with immunotherapies (Fig. 5B).

##### ii. Association of treatment with on-label markers of resistance

We next used the alterations described above to uncover the associations between ESCAT resistance markers and the treatments received before the biopsy in META-PRISM WES & RNAseq tumors. Additionally, we retrieved ‘novel’ resistance markers that are supported by the literature but are not yet included in the versions of OncoKB and CIViC databases used for this analysis (Supplementary Table S10).

We could only identify resistance mechanisms for a minority of treatments. SOC resistance events were detected only for first and second-generation *EGFR* inhibitors in LUAD for 34% of treated patients and *EGFR* antibodies in COAD for one in five treated patients (Fig. 6A, 6F). In contrast, emerging resistance markers substantially increase the fraction of observed treatment resistances that may be explained. In LUAD, Tier 2&3 resistance mutations were identified in three of 17 patients that progressed on first- or second-generation *ALK* inhibitors and five of 28 patients that progressed on immunotherapies (Fig. 6A). BRCA patients treated with hormone therapy harbored Tier 3 resistances through *ESR1* mutations in 32% of cases (Fig. 6B). PRAD patients treated with hormone therapy harbored Tier 2 resistances through *AR* alterations in 31% of cases (Fig. 6C). In COAD, we also observed Tier 2 markers of resistance to EGFR antibodies (Fig 6D). In PAAD, we did not identify any resistance markers for the drugs patients received, and in general, no mechanism of resistance was identified in our cohort for chemotherapies or antiangiogenic treatments.

**Fig 6.**
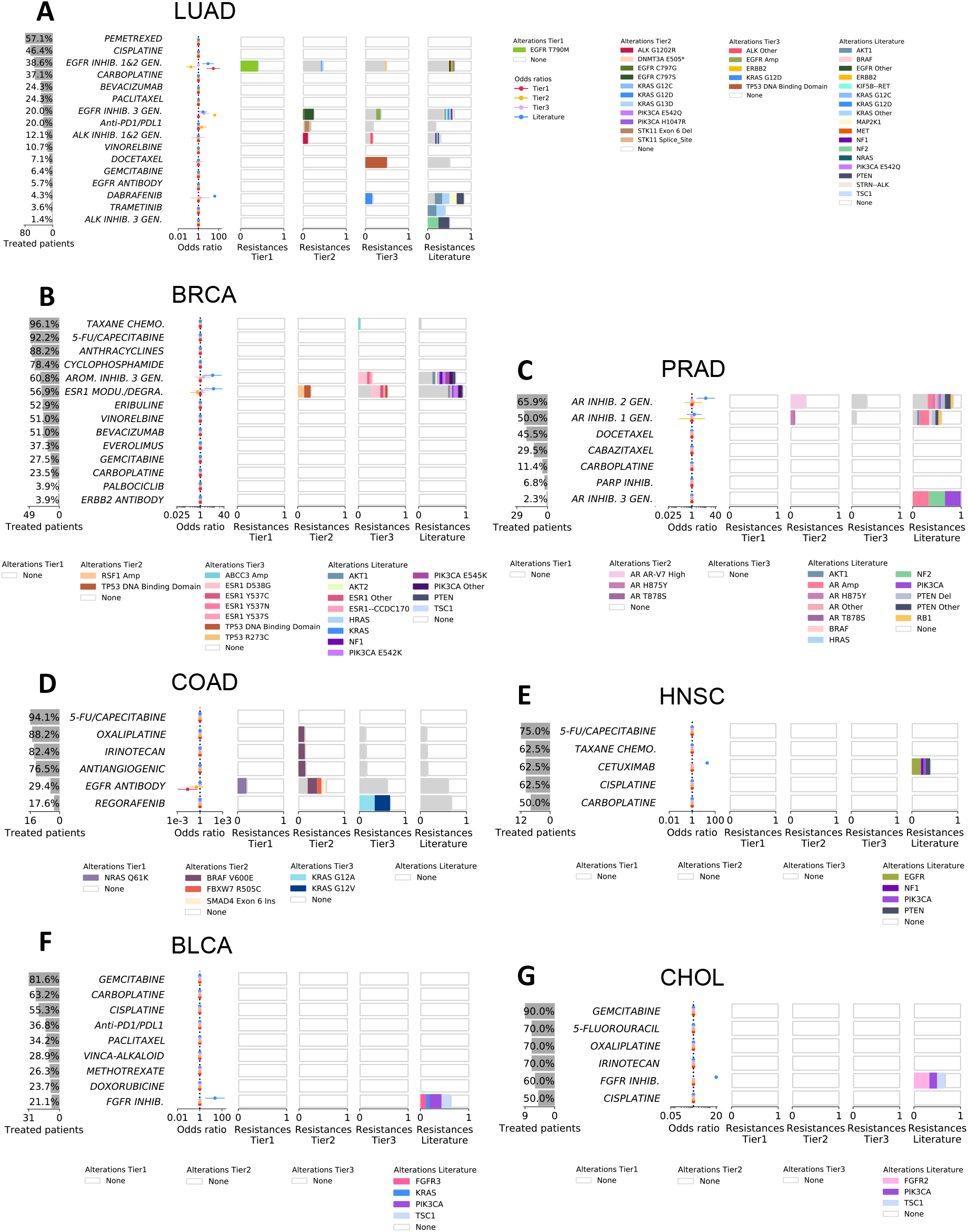
Associations between treatments received and molecular markers in META-PRISM. **A.** The left bar plot indicates the percentage of LUAD patients in META-PRISM WES & RNAseq subcohort that received each treatment or group of treatments. The middle error bar plot represents the odds ratio and 95% confidence interval estimates (Supplementary Methods) for the enrichment of resistance markers of Tier 1, Tier 2, and Tier 3 or extracted from the literature in the patients that received the corresponding treatment. The right group of four bar plots provides details about the identity of the markers associated with resistance in treated patients at each of the four confidence levels. **B.**, **C.**, **D.**, **E.**, **F.**, and **G.** Identical to **A.** for BRCA, PRAD, COAD, HNSC, BLCA, and CHOL patients, respectively.

Additionally, we were able to associate ‘novel’ resistance mechanisms with a considerable fraction of resistances to targeted and hormonal therapies. For example, 13%, 25%, and 67% of LUAD patients treated with first- or second-generation *EGFR* inhibitors, third-generation *EGFR* inhibitors, and *BRAF* inhibitors (dabrafenib), respectively, harbored hypothetical resistance mechanisms with literature support (Fig. 6A, Supplementary table S10). Interestingly, one out of the two patients that progressed on lorlatinib (third-generation *EGFR* inhibitor) harbored an *EML4-ALK* fusion and a double *ALK* mutation (p.F1174L. and p.G1202R) as previously described ^40^. In BLCA and CHOL, we were able to associate mutations in *FGFR1*, *FGFR2*, *PIK3CA*, *KRAS*, and *TSC1* with resistance to *FGFR* inhibitors (Fig. 6F, 6G). In HNSC, we associated driver mutations in *EGFR*, *NF1*, *PIK3CA*, and *PTEN* to *EGFR* inhibitor cetuximab (Fig. 6E).

The observations above show that SOC resistance biomarkers were associated with only 1.9% of received treatments, while investigational, hypothetical, and ‘novel’ mechanisms could further explain 2.9%, 2.3%, and 6.3% of resistances, respectively.

#### b. Survival analyses

The association of molecular markers with overall survival measured from the day of diagnosis has been the subject of extensive work ^19 20 21 22 23 24 25^, some of which has become SOC. However, the association of these markers with survival in metastatic settings has been scarcely explored, and it remains unclear how genotyping may inform decision-making in investigative trials for advanced tumors.

##### i. Univariate regressions

We investigated how driver genes are univariably associated with metastatic cancer patient survival in META-PRISM WES & RNAseq tumors. Twenty-one genes that were altered (SCNA, mutations, fusions) in more than 5% of tumors were considered for the analysis. Only *TP53* gene showed an association with survival (adjusted HR=1.44; CI 95% [1.01, 2.05]) after adjusting for the tumor type composition (Fig. 7A, upper left). Analyses per tumor type revealed an association of *STK11* events with poor prognosis in LUAD and of *ERG* events with favorable prognosis in PRAD (Fig. 7A, upper right and middle left).

**Fig 7.**
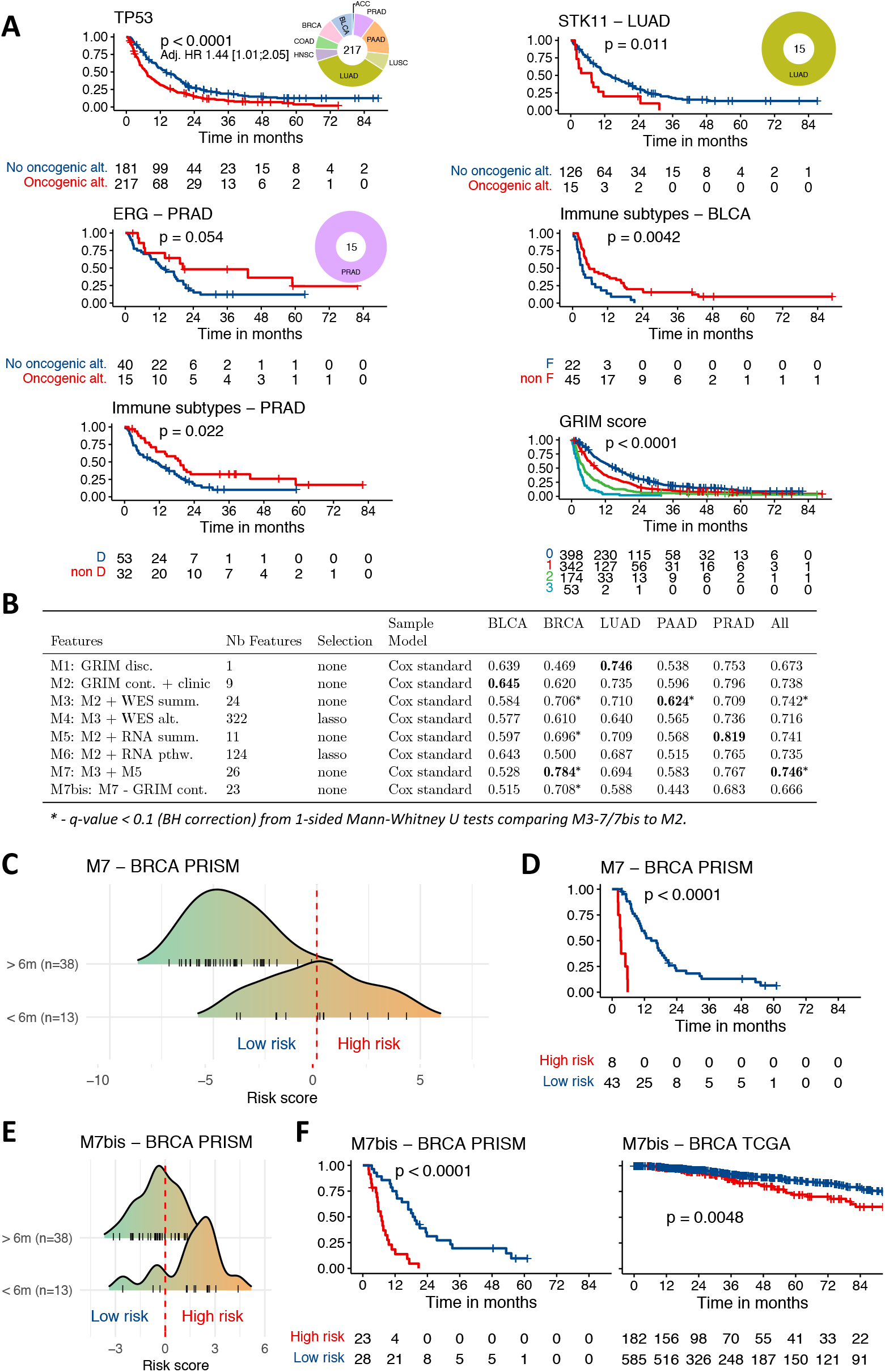
Kaplan-Meier curves for significant predictors and prediction of six-month survival using clinical characteristics and combinations of WES or RNAseq features. **A.** Kaplan-Meier curves of META-PRISM WES & RNAseq tumors with and without *TP53* oncogenic alterations (upper left), LUAD tumors with and without *STK11* oncogenic alterations (upper right), PRAD tumors with and without *ERG* oncogenic (middle left), BLCA tumors with fibrotic (F) and without fibrotic (non-F) tumor-microenvironments (middle right), PRAD tumors with depleted (F) and without depleted (non-D) tumor-microenvironments (bottom left), and all META-PRISM tumors classified into four categories as determined by the GRIM score (bottom right). All p-values are computed from log-rank tests. Hazard ratios are computed by fitting univariate Cox models except for *TP53*, where the cancer type was included in the model. **B.** Averaged C-indexes on the 1,000 cross-validation subsamples for each combination of features analyzed. Cox models were fit on META-PRISM patients with both WES and RNAseq data and from the five most frequent tumor types or a pan-cancer cohort composed of patients belonging to the ten most prevalent tumor types. M1: discrete GRIM score. M2: continuous components of GRIM score, age, tumor type, and gender are considered. M3: variables from M2 and WES summary statistics are considered. M4: variables from M2 and WES-derived alterations in driver genes (lasso reduction applied, Supplementary Methods). M5: variables from M2 and RNAseq-derived fusion burden and TMEs. M6: variables from M2 and RNAseq-derived gene expression signatures (lasso reduction applied). M7: variables included in M3 and M5 are considered. M7bis: variables M7 without the components of the GRIM score. **C.** Distribution of the predicted risk scores from the Cox model M7 in META-PRISM BRCA patients that had a survival time greater or lower than six months from the biopsy date. One patient that was censored before six months was included in the “> 6m” group. **D.** Kaplan-Meier curves for META-PRISM BRCA patients predicted to be high-risk (risk score > 0, Supplementary Methods) or low-risk (risk score < 0) using the M7 model. **E.** Identical to C. using the M7bis model. F. Kaplan-Meier curves for META-PRISM BRCA (left) and TCGA BRCA (right) patients predicted to be high-risk (risk score > 0) or low-risk (risk score < 0) using M7bis model.

TME classifications, which have already been shown to be prognostic^15 16^, were associated with survival in META-PRISM. Indeed, immune-cold subtypes (F and D) had worse survival compared to immune-enriched subtypes (IE/F and IE) (OR IE/F + IE vs. F + D). Strikingly, the F subtype in BLCA and D subtype in PRAD, both enriched in META-PRISM vs. TCGA, were also associated with the poorest prognosis in the corresponding tumor type (p=0.004, F vs. non-F BLCA; p=0.02, D vs. non-D in PRAD) (Fig. 7A, middle right and bottom left).

##### ii. Cox multivariate regressions combining clinical, genomic, and transcriptomic variables for prediction of six-month survival

Patients with advanced metastatic cancer are characterized by severe physiological deterioration as measured by lactate dehydrogenase levels, serum albumin, or neutrophil-to-lymphocyte ratio. The GRIM score ^13^, which combines these physiological markers, can predict six-month survival in the META-PRISM patients with an average concordance index of 0.67 (Fig. 7B; Supplementary Methods). We then investigated if the prediction of survival at this stage of the disease could be improved by considering genetic markers engineered from the previous analyses. The list of genetic markers retrieved from WES was composed of summary statistics, including SCNAs, WGD status, MSI score, TMB, and the presence of oncogenic alterations aggregated into genes or pathways discriminating between ESCAT levels. The list of markers retrieved from RNAseq included TME subtypes and gene expression signatures measuring the activity of immune pathways, activation of general pathways ^41^, or main transcription factors ^42^. In order to compare the added prognostic value from different categories of markers to the objective clinical markers, we ran multiple Cox proportional hazard regressions on the META-PRISM WES & RNAseq subcohort and each of the five main tumor types considering only the samples with both WES and RNAseq (Supplementary Methods).

As expected, the models’ performances based on discrete GRIM score (M1 model) were significantly improved by using the continuous metrics underlying it and by adding baseline clinical variables (age, gender, tumor type, clinical subtypes; M2 model). Nevertheless, the incorporation of WES-derived markers (M3 and M4 models), RNAseq-derived markers (M5 and M6 models), or both (M7 model) resulted in a further increase of the C-index over the continuous GRIM score model (M2) in all analyzed tumor types and for the full cohort except for LUAD where the discrete GRIM score classification was the most prognostic (Fig. 7B).

Strikingly in BRCA tumors, for which the GRIM score had no prognostic value in META-PRISM, the consideration of genetic markers engineered from WES or RNAseq was highly predictive of six-month survival. The best-performing model for BRCA utilized WES and RNA summary statistics (C-index increased by 16% in M7 vs. M2 model, reaching 0.784). The predicted risk scores from this model were significantly different between patients with a survival time greater or lower than six months from the date of the biopsy (Fig. 7C). By discriminating between patients having a positive risk score (poor prognosis, 8/51) and a negative risk score (good prognosis, 43/51), the model was able to split BRCA patients into two groups with very different clinical outcomes (p < 0.0001; Fig. 7D). Analysis of the M7 model coefficients in BRCA revealed the independent prognostic value of: (i) the total number of tier 1 alterations (LHR 3.75; CI 95% [1.07, 6.53]), the fraction of the genome covered by (ii) focal deletions (LHR -4.05; CI [-8.73, -0.34]), by (iii) focal amplifications (LHR 7.28; CI 95% [2.63, 13.55]), and by (iv) middle to low-level gains (LHR -3.65, CI 95% [-8.18, -1.22]), and (v) the TME subtypes (LHR -3.18 for F vs. D, -2.72 for IE vs. D, -4.80 IE/F vs. D) (Supplementary Fig. S13). In order to validate the survival model on an external cohort, we considered one extra model (M7bis) that was identical to M7 except that the components of the GRIM score were excluded. M7bis was also predictive of survival in META-PRISM BRCA (C-index=0.71) and in TCGA BRCA (C-index=0.64; Fig. 7E, 7F).

## Discussion

Our investigation of the META-PRISM cohort describes the genetic and transcriptomic variations across a wide range of refractory metastatic tumors from 39 cancer types. This cohort is characterized by a short survival time after the biopsy date and by a high proportion of multi-resistant tumors or rare tumors with no approved therapy options. Consequently, the genomes of these tumors represent a much-advanced evolutionary stage containing the footprints of mutagenic treatments and therapeutic pressure. To characterize the genetic traits specific to this cohort and assess how these may inform the tumor’s aggressiveness and resistance to therapies, we compared META-PRISM to > 10,000 primary untreated tumors from TCGA ^43 1^ and validated all results using an external cohort of 500 metastatic tumors ^3^.

Our analysis reveals that several types of genomic instability were strongly enriched in refractory cancers, particularly the mutation rate, the frequency of whole-genome duplications, and the fraction of the genome covered by focal copy number alterations. These results are consistent with previous studies that reported increased genomic instability and mutational burden in metastases of different cancer types ^44 5 2 45 46 47^. Correlative analyses between the mutational profiles of tumors and the history of treatments received have shown increased activity of signatures SBS31 and SBS35 in tumors treated with cisplatin and to a lesser extent with carboplatin. The high mutation rate of these two signatures supports previous observations of strong mutational footprints caused by platinum treatment in metastatic cancers ^12^ and relatively low mutagenic effects of oxaliplatin ^48^. The driver fusions and SCNAs represented 9.4% and 21% of all detected variation in driver genes, respectively, and were strongly enriched in META-PRISM tumors compared to TCGA tumors. However, these two types of events were associated with treatment resistance in 0% and 6.2% of META-PRISM patients, respectively, reflecting their scarce resistance annotations in OncoKB and CIViC.

Our research also reveals that current annotations of resistance mechanisms can only account for a small proportion of all observed treatment resistances, as standard-of-care resistance markers were found in 73.4% of patients but could only explain 1.9% of all resistances. Investigational, hypothetical, and ‘novel’ mechanisms could further explain 2.9%, 2.3%, and 6.3% of resistances. These low percentages were observed even though patient inclusion in MOSCATO and MATCHR trials was driven by the possibility of using innovative treatments. This data highlights the unmet need for large-scale efforts that combine molecular profiling with exhaustive clinical annotations to fill our current lack of understanding of resistance mechanisms in cancer.

Interestingly, MSI and high TMB were detected in 3% and 13% of META-PRISM tumors, respectively, suggesting that a considerable fraction of metastatic cancers refractory to conventional treatments could benefit from immune checkpoint inhibitor therapies, such as pembrolizumab^49 50^. Additionally, a higher fraction of pathogenic cancer-risk germline variants was observed in META-PRISM than in the general population or the comparison cohort of primary tumors. The increased incidence of predisposing mutations suggests that metastatic cancer patients could benefit from germline testing and genetic counseling.

Markers of physiological deterioration, such as levels of albumin, neutrophils, lymphocytes, and LDH, are used in objective risk scoring systems to predict patient survival, which is essential for assessing the eligibility of these patients to phase I clinical trials ^51 52^ This study shows that tumor genomic and transcriptomic features can be used to improve the accuracy of predictions based on objective risk factors at this stage of the disease. The added value of genetic markers to current prognostic scores is currently limited at the pan-cancer level, likely due to the high heterogeneity of mechanisms driving tumorigenesis in each tumor type. However, models incorporating genetic markers are significantly more accurate in predicting six-month survival in BRCA refractory tumors, showing that whole-exome and RNA sequencing are important for accurately establishing the individual risk profile of late-stage cancer patients.

This study validates the previous combined genomic and transcriptomic descriptions from pretreated pan-cancer metastatic diseases ^53 54^. It also highlights the feasibility of precision medicine in clinical routine and the benefit of access to new drug protocols for patients without standard treatment^54 55^.

The present cohort advances translational cancer genomics by providing a unique resource combining detailed clinical data with exome and transcriptome profiling.

## Methods

### 1. META-PRISM patient cohort

Patients 18 years or older with unresectable or metastatic cancer that had progressed during at least one line of prior therapy or lacked treatment options, that had at least one site accessible to biopsy, and that were considered non-curable by a multidisciplinary board were included as part of the MATCH-R (NCT02517892) and MOSCATO (01/02) (NCT01566019) clinical studies. These studies were approved by the Institutional Review Board (IRB) of Gustave Roussy and were executed in accordance with the Declaration of Helsinki and Good Clinical Practice guidelines. In this study, we included biopsies of patients enrolled in MATCH-R, MOSCATO, or both (302, 697, and 32 biopsies, respectively). In cases where multiple biopsies were available (84 cases), the latest sequenced biopsy was selected for the study (with the exception that preference was given to samples with DNA and RNA data). All patients have given consent for the trial, genomic/transcriptomic analyses, and data sharing for cancer research purposes. Needle biopsies were sampled from a metastatic lesion whenever possible or the primary tumor otherwise and were frozen in liquid nitrogen. A senior pathologist assessed the tumor cellularity on the hematoxylin and eosin (H&E) slide from the biopsy that was used for DNA and RNA extraction. Samples with tumor cellularity >30% were selected for the genetic analysis.

Clinical variables were manually collected by clinicians or extracted from electronic health records stored on the Dr. Warehouse database (PMID: 29501921) using semi-automatic methods. Semi-automatic data collection consisted in extracting text chunks containing specific patterns of interest that were further processed by automatic scripts and subjected to manual inspection and correction by three different oncologists. Through these techniques, we were able to collect data regarding patients’ demographics (age at biopsy, gender), tumor characteristics (primary tumor site, tumor histology, date of first cancer diagnosis, metastatic status at diagnosis, metastatic sites number, and localization at the time of MOSCATO/MATCH-R inclusion), blood test results performed within one month of the tumor biopsy date, and history of treatments received before the biopsy. All anti-cancer drugs were classified into either conventional chemotherapy (further subdivided into platinum salts, alkylating agents, and non-alkylating agents), targeted therapies (further subdivided on the basis of the drug target; e.g., EGFR inhibitors, ALK inhibitors, BRAF inhibitors, etc.), hormone therapies, or immunotherapies (further subdivided into immune checkpoint inhibitors and non-immune checkpoint inhibitors). Patients were considered resistant to a systemic treatment if they had progressed under or after the treatment and before the biopsy date. All clinical variables are tabulated in Supplementary table S1.

The Gustave Roussy Immunoscore (PMID: 32047540) was calculated based on the blood test results measured within one month of the biopsy date (lactate dehydrogenase levels, albumin concentration, neutrophil counts, and lymphocyte counts). Survival time was calculated from the date of biopsy to the date of death from any cause. Out of 1,031 patients, dates of death and dates of last news were available for 1,029 patients (99.81%). At the time of the present study, 909 patients had died (88.17%). Follow-up time ranged from 2 days to 2667 days (Q1 127-Q3 616 days).

### 2. DNA library preparation and exome sequencing

Tumor DNA and whole-blood germline DNA were extracted using AllPrep DNA/RNA Mini Kit (Qiagen) and DNeasy Blood and Tissue Kit (Qiagen) according to the manufacturer’s instructions. Library preparation, exome capture, sequencing, and data analysis have been performed by IntegraGen SA (Evry, France).

Genomic DNA was captured using Agilent Human All Exon V5, CR, and CR2 capture kits (for samples processed before the year 2019) or Twist Human Core Exome Enrichment System (Twist Bioscience (for samples processed after the year 2019). Sequence capture, enrichment, and elution were performed according to the manufacturer’s instructions and protocols without modifications. For library preparation, at least 150 ng of each genomic DNA was fragmented by sonication and purified to yield fragments of 150-200 bp. Paired-end adaptor oligonucleotides from the NEB kit were ligated to repaired and a-tailed fragments, then purified and enriched by 4-7 PCR cycles. 500ng of these purified libraries were then hybridized to the SureSelect or Twist oligo probe capture libraries. After hybridization, washing, and elution, the eluted fraction was PCR-amplified with 8 -10 cycles, purified, and quantified by qPCR to obtain enough DNA templates. Each DNA sample was then sequenced on an Illumina HiSEQ 2000 as paired-end 75bp reads (before the year 2019) or on an Illumina NovaSeq as paired-end 100bp reads (after the year 2019). Image analysis and base calling were performed using Illumina Real Time Analysis with default parameters. Supplementary table S2 provides details about the kits used for each DNA sample.

Quality control of paired-end reads was accomplished using FastQC v0.11.8^1^. Fastp v0.20 (PMID: 30423086) was subsequently employed for trimming adaptors and polynucleotide tracts from reads which were longer than 25 nucleotides. Afterward, the resulting cleaned FASTQ files were aligned to the reference human genome GRCh37 using BWA-MEM v0.7.17 (PMID: 19451168). Intermediate BAM files were further processed for deduplicating reads using MarkDuplicates from Picard v2.20.3^2^, sorting coordinates using SAMtools v1.9 (PMID: 19505943), and finally recalibrating their base qualities using BaseRecalibrator and ApplyBQSR. All these tools are included in the GATK bundle v4.1.8.1 (PMID: 21478889). Alignment quality was controlled using three different algorithms: mosdepth v0.2.5 (PMID: 29096012), flagstat from SAMtools v1.9 (PMID: 19505943), and CollectHsMetrics from GATK v4.1.8.1.

### 3. RNA library preparation and transcriptome sequencing

Tumor RNA was extracted using AllPrep DNA/RNA Mini Kit (Qiagen) according to the manufacturer’s instructions. RNA libraries were prepared with TruSeq Stranded mRNA kit (for samples processed before the year 2019) or with NEBNext Ultra II Directional RNA Library Prep Kit for Illumina protocol (for samples processed after the year 2019) according to supplier recommendations.

The purification of PolyA-containing mRNA molecules was performed using oligo(dT) coupled to magnetic beads from 1µg total RNA (with the Magnetic mRNA Isolation Kit from NEB). Fragmentation was performed using divalent cations under elevated temperature to obtain 300-400bp fragments, followed by double-strand cDNA synthesis, Illumina adapters ligation, and cDNA library amplification by PCR for sequencing. Paired-end read sequencing was then carried out on Illumina NextSeq 500 (75bp; before the year 2019) or on Illumina NovaSeq (100bp; after the year 2019). Image analysis and base calling were performed using Illumina Real Time Analysis (3.4.4) with default parameters. Supplementary table S2 provides details about the kits used for each RNA sample.

Quality control of reads was performed with Trim galore^3^ (v0.4.4). Reads were then pseudo-aligned to the human transcriptome from GENCODE v27 using Kallisto (v0.44.0, PMID: 27043002), and transcript-level estimates were aggregated to the gene-level (58,288 genes) using TxImport (v1.16.0, PMID: 26925227).

### 4. Germline and somatic calling of point mutations and small indels

Germline single nucleotide variants (SNVs) and indels were called using HaplotypeCaller (DOI: 10.1101/201178). After calling, putative germline variants were filtered using hard thresholding on QualByDepth (QD > 2), genotype quality (QUAL > 30), FisherStrand (FS < 60 for SNV, < 200 for indels), ReadPosRankSumTest (ReadPosRankSum > -8 for SNVs, > - 20 for indels), RMSMappingQuality (MQ > 40 for SNVs only), and MappingQualityRankSumTest (> - 12.5 for SNVs only) as recommended in GATK best practices^4^. Variants that passed all filters were annotated using ANNOVAR (PMID: 20601685).

Somatic point mutations and small indels were detected using Mutect2 (PMID: 23396013). In order to remove artifacts and false positives, a panel-of-normal was created from normal samples and used at the Mutect2 calling step as specified in the GATK best practices^5^. Putative variants were then analyzed for read orientation artifact and sample contamination by running GATK LearnReadOrientationModel and CalculateContamination again as recommended in the best practices.

The following set of filters was applied

- Not filtered by Mutect2 (MUTECT_FILTERS).
- Minimum VAF of 5% (LOW_VAF).
- Minimum sequencing coverage of 20X in the tumor sample (LOW_COVERAGE_TUMOR).
- Minimum sequencing coverage of 10X in the normal sample (LOW_COVERAGE_NORMAL).
- Located inside exonic regions as defined by the set of canonical transcripts used by VEP v104 on GRCh37 assembly (NOT_EXONIC).
- Allele frequency across all gnomAD v2.1 exome subpopulations is < 0.04% (COMMON_VARIANT). This rule is not applied for variants annotated by OncoKB (see section 12).
- Localized within the META-PRISM target region, which is defined by the intersection of the capture regions of all four different kits used (OFF_TARGETS_INTERSECTION). This region spans approximately 36.6 Mb.

A total of 118,216 somatic point mutations and small indels were used for analysis. Supplementary Methods Fig. 1 summarizes the effect of each filter individually and in combination with other filters.

All PASS mutations were annotated using VEP release 104 (PMID: 27268795) on canonical transcripts. We took advantage of the plugin feature of VEP to enhance the annotations with different metrics from the dbNSFP v4.2 database.

### 5. Microsatellite Instability (MSI) detection

MSI analysis was performed with MANTIS v1.0.3 following the same procedure that was described in the original study (PMID: 27980218). In order to run MANTIS, a list of microsatellite loci has to be compiled in a 6-column BED file, which includes not only the genomic coordinates but a column with the motif and another column with its count on the reference genome. In our analysis, we employed the same BED file^6^ that was used in the TCGA study (PMID: 29850653) and includes 2530 loci. The method scrutinizes repetitive regions in aligned reads from the tumor and normal BAM files, each locus at a time. Per locus read count for each repeat motif is calculated in the tumor and normal and used for computing an instability score. Subsequently, the average of all locus instability scores is computed to produce a final score for the tumor/normal pair. Scores reported range from 0.0 (entirely stable) to 2.0 (entirely unstable). MSI-H was called in samples that had a final score exceeding the default threshold of 0.4.

### 6. Somatic copy number alterations (SCNA) and whole-genome duplications (WGD) calling

SCNAs, tumor purity, and average tumor ploidy were identified with the FACETS R package v0.5.14 (PMID: 27270079) run with parameters cval_pre=25 and cval_pro=500. Lower copy number (LCN) and total copy number (TCN) are assessed by FACETS for each copy number segment identified.

The number of WGDs was determined to be the lowest positive integer k such that at least 11 autosomes have undergone k duplications. An autosome is identified to have undergone k duplications if the major allele ploidy (calculated as TCN-LCN) is strictly superior to 1.5*2^k-1^ on at least half of the chromosome length. If this condition cannot be fulfilled with k=1, we assume that no WGD has occurred. This rule mirrors the simple heuristic used in Priestley et al. (PMID: 31645765) for identifying WGD-positive samples (regardless of the number k of duplications).

The ploidy of SCNAs in all chromosomes was then categorized into six classes (similarly to what was done in PMID: 31645765) taking into account the estimated number of WGDs (0 or k>=1) and the patient’s gender, as detailed below.

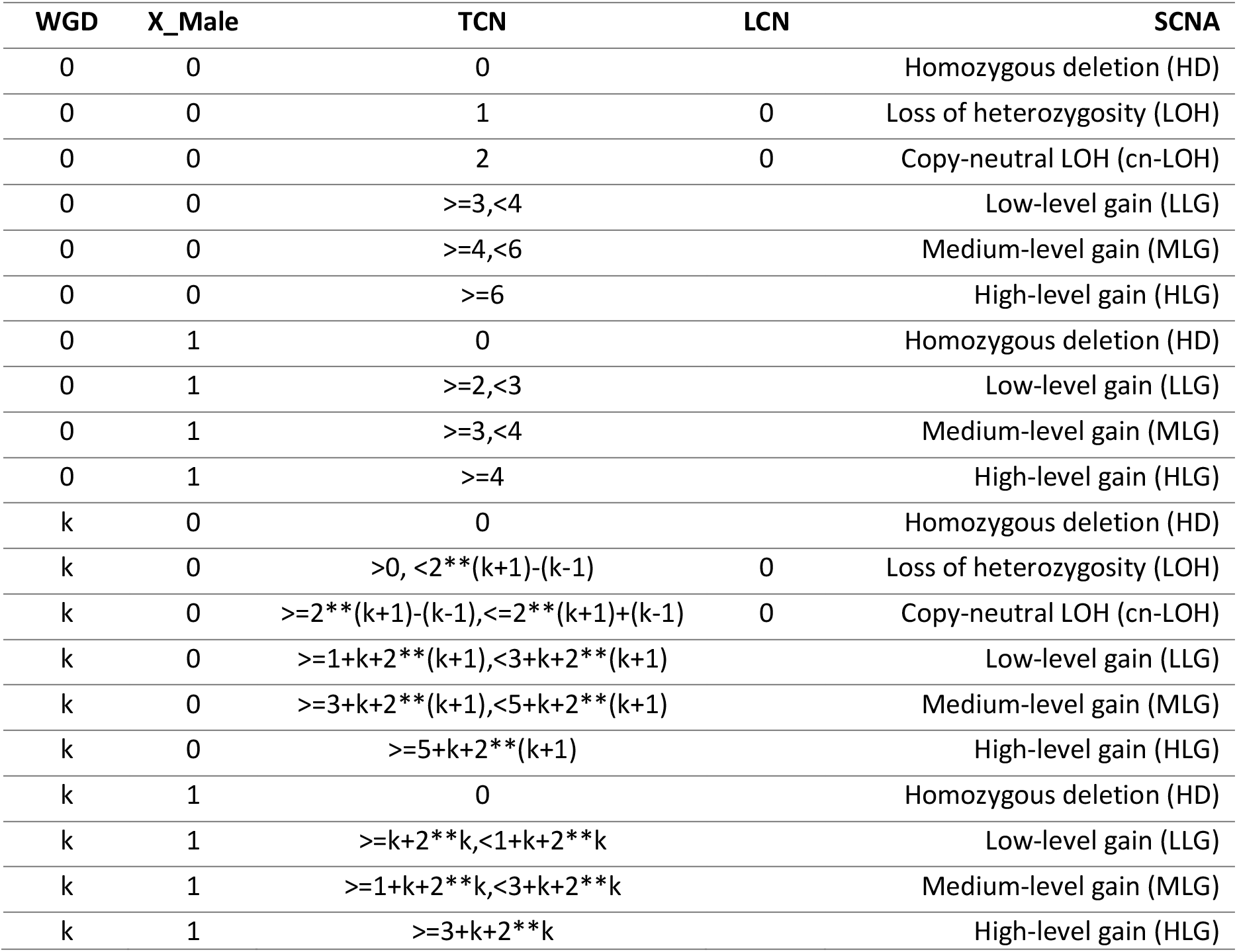

As WGD is a strong confounding factor for the accurate calling of low-level and middle-level SCNAs, these events were only retained in samples without WGD. Consequently, the fractions of low and middle-level SCNAs at each locus in Figure 3 were estimated only among nearly diploid tumors, while high-level SCNAs were computed considering the full cohort.

Additionally, only high-level gene SCNAs (HD and HLG) from focal events (segments smaller than 10 Mb) were considered further for the description of driver events (see section 12).

### 7. Mutational signature analysis

Only samples with at least 50 detected somatic point mutations were used for the mutational signature analysis, resulting in the selection of 452 out of 571 samples with WES data available. Point mutations from these samples were summarized into a matrix of mutation counts with 96 rows for the 96 tri-nucleotide mutation categories (disregarding the strand) and 452 columns, totaling 105,606 point mutations. Mutational signatures activities in each sample were determined by projecting this matrix onto the database of SBS signatures COSMIC v3.2^7^ (78 signatures, PMID: 32025018) using the MutationalPatterns R package v3.2.0. We then made an effort to mirror the original algorithm used for deconvoluting mutational signatures (SigProfiler MATLAB package^8^, also available in Python on Alexandrov’s lab Github page^9^). For that, we assessed the individual contribution of each signature to the mutation profile of each sample and removed signatures that contributed to less than 1% of the profile as measured by the decrease in cosine similarity by removing the signature. Mutations from removed signatures were redistributed among the other signatures using a non-negative least-squares procedure. We used the part of the Julia code^10^ (that reimplements the SigProfiler algorithm) from Pich et al. 2019 (PMID: 31740835) to run this sparsity-inducing step.

### 8. Identification of gene fusions

Gene fusions were called on the GRCh38 reference genome by filtering and combining fusions predicted by Arriba v1.2.0 (PMID: 21450565), EricScript v0.5.5 (PMID: 23093608), Pizzly v0.37.3 (PMID: 28422140), and StarFusion v1.8.1 (PMID: 31639029), all included in the nf-core/rnafusion pipeline v1.2.0^11^.

For each individual caller, the set of putative gene fusions was limited to exclude fusions that have been previously reported in studies of normal tissues (blacklists) and to retain only the fusions that have been reported in studies of cancer tissues (whitelists) or that involve a cancer driver partner (see section 15.b.6 for all the details of the lists used). After filtering, only gene fusions seen by both Arriba and EricScript or by both Pizzly and StarFusion, independently of the predicted breakpoints, were included (see section 15.b.6 for detailed explanations about this specific combination).

A total of 851 gene fusions (disregarding breakpoints, see Supplementary Methods Fig. 2) were detected in 445 out of 944 samples with RNAseq data and successfully processed by all fusion-calling algorithms (3 samples repeatedly failed on one or more algorithms). Details for the 767 gene fusions from the 796 samples included in RNAseq analyses and successfully processed (2 samples repeatedly failed) are available in Supplementary table S8.

### 9. Validation of gene fusions

RNA from tumor samples was extracted using AllPrep DNA/RNA mini kit (Qiagen) combined with Trizol LS (ThermoFisher). cDNA was synthesized with 50ng of RNA using High-Capacity cDNA Reverse Transcriptase kit (LifeTechnologies). PCR amplification was performed using Fast Start PCR Master (Roche). We used the following program PCR amplification: hot start 95°C for 2mn, then 38 cycles of 95°C for 30sec., 58°C for 1mn and 72°C for 1mn. A final extension was done at 72°C for 7mn (VeritiPro Thermal Cycler, Thermo Fisher Scientific). Electrophoretic migration was performed in TBE1x 2% agarose gel. PCR products were purified according to size with a QIAquick PCR purification kit for single bands or a QIAquick Gel Extraction kit (Qiagen) for multiple bands. Eurofins Genomics synthesized the primers and did the sequencing. Out of 18 gene fusions tested, 16 exhibited a positive PCR (Fig. 6E).

### 10. Germline cancer-risk variant calling

To find germinal cancer-predisposing variants, we used a conservative approach aiming to maximize the representation of variants with strong effects in our dataset. After annotation with ANNOVAR, we extracted variants that met all the following criteria:

- the variant is in a curated list of 130 cancer-predisposing genes. The list was established starting from the list of 152 genes curated by Huang et al. 2018 (PMID:29625052), from which we additionally excluded 22 genes after literature curation showing that these genes have only weak support for being cancer-predisposing. These 22 excluded genes are *ABCB11*, *ELANE*, *FAH*, *FH*, *GBA*, *GJB2*, *HFE*, *HMBS*, *HNF1A*, *ITK*, *MTAP*, *PHOX2B*, *PMS1*, *PRF1*, *SBDS*, *SERPINA1*, *SLC25A13*, *SRY*, *STAT3*, *TNFRSF6*, *UROD*, *WAS*
- the variant has a maximum general population allele frequency of 5% in the gnomAD v2.1.1 exome database (PMID: 32461613).
- the variant is annotated in ClinVar database (PMID:29165669) as “Pathogenic”, “Likely_pathogenic”, “Pathogenic/Likely_pathogenic” or is located in a tumor suppressor gene and has protein-disrupting role (“splicing”, “frameshift deletion”, “frameshift insertion”, “stopgain”, or “stoploss”) except for the POLD1 and POLE tumor suppressors for which only variants annotated by ClinVar were retained.

Candidate variants were then manually reviewed in the BAM files. A total of 98 cancer-predisposing variants were detected in 75 out of 571 META-PRISM samples with germline data available and included in analyses (14 samples harbored two or more such variants).

### 11. Discovery of cancer driver genes from the cohort

MutPanning v2.0 (PMID: 32015527) was run on the full META-PRISM with the specification of the tumor types and the default options.

### 12. Cancer gene list and catalog of oncogenic events

We compiled a list of 360 cancer driver genes by intersecting the list of driver genes (tiers 1 and 2) from COSMIC census v92 and the list of genes annotated in the OncoKB database as of July 2021 (PMID: 28890946).

Oncogenic events were identified by intersecting point mutations, indels, high-level gains (HLG), homozygous deletions (HD), and gene fusions with the OncoKB and CIViC (PMID: 28138153) databases. OncoKB constitutes a literature- and knowledge-based database that is accessible through the web API after having registered an account and requested a token. In accordance with the type of event to be annotated, different scripts from oncokb-annotator^12^ have to be employed when operating with the OncoKB API:

- MafAnnotator.py for point mutations and indels.
- CnaAnnotator.py for SCNAs.
- FusionAnnotator.py for gene fusions.

CIViC is also a literature and knowledge-based database, but no API was available yet at the time of our analyses. As a consequence, we have developed in-house scripts to annotate point mutations, indels, gene fusions, HLGs, and HDs using the table of clinical evidence summaries 01-Jan-2022-ClinicalEvidenceSummaries.xlsx from the January 2022 release^13^. A minor number of errors were manually curated from the table. Additionally, missing genomic coordinates for point mutations and indels were manually filled where possible.

Importantly, the majority of OncoKB and CIViC annotations are tumor-type specific. Consequently, the annotation algorithms require an extra file containing the tumor type of each sample to allow for precise on-label annotations. OncoKB uses the MSKCC’s oncotree nomenclature, while CIViC uses designations that do not follow specific rules and have a varying degree of specificity. A thorough work of tumor type matching was performed by a panel of oncologists to navigate between TCGA, MSKCC’s oncotree, and CIViC designations.

All HLGs, HDs, and gene fusions found in either OncoKB or CIViC databases were retained. However, point mutations and indels that intersected with the CIViC database but not the OncoKB database were discarded, given that manual inspection of these CIViC-only events revealed false matching arising from an unspecific variant description in CIViC. Among the point mutations and indels that were annotated by oncokb-annotator, all events with MUTATION_EFFECT as “Likely Neutral”, “Neutral”, or “Unknown” were discarded unless the ONCOGENIC field was “Likely Oncogenic” or “Predicted Oncogenic”. Lastly, only point mutations and indels that were classified as either “Missense_Mutation, “Frame_Shift_Del”, “Frame_Shift_Ins”, “In_Frame_Del”, “In_Frame_Ins”, Nonsense_Mutation”, “Splice_Site” or “Translation_Start_Site” were retained.

### 13. Clinical applicability of oncogenic events

After the catalog of oncogenic events was defined as described in the previous section, we took advantage of these databases to assess how these events may inform treatment decisions. OncoKB provides indications of how events may be used to inform treatment indications (sensitivity) or counter-indications (resistance) on 5-level and 3-level confidence scales (version from July 2021), respectively. Similarly, CIViC provides the same type of indications but uses a 5-level confidence scale for both types. Sensitivity and resistance confidence scales were harmonized against a common 3-level reference scale following ESCAT guidelines (PMID: 30137196).

ESCAT Tier 1 level (standard-of-core or SOC) is for biomarkers that are used in clinical practice. These were identified as:

- CIViC Level A (proven/consensus association in human medicine).
- OncoKB Level 1 (FDA-recognized biomarker predictive of response to an FDA-approved drug in this indication).
- OncoKB Level 2 (standard-of-care biomarker recommended by the NCCN or other professional guidelines predictive of response to an FDA-approved drug in this indication).
- OncoKB Level R1 (standard-of-care biomarker predictive of resistance to an FDA-approved drug in this indication).

ESCAT Tier 2 level (investigational) encompasses investigational targets that likely define a patient population that benefits from a given drug but for which additional data are needed. These were identified as:

- CIViC Level B (clinical trial or other primary patient data support association).
- OncoKB Level 3A (compelling clinical evidence supports the biomarker as being predictive of response to a drug in this indication).
- OncoKB Level R2 (compelling clinical evidence supports the biomarker as being predictive of resistance to a drug).

ESCAT Tier 3 level (hypothetical) includes all targets that have demonstrated a clinical impact on other tumor types or that are supported by scarce data (often from case reports only). These were identified as:

- CIViC Levels C, D, and E (supported by case-study, preclinical and inferential data, respectively).
- OncoKB Level 3B (standard-of-care or investigation biomarker predictive of response to an FDA-approved drug in another indication).
- OncoKB Level 4 (compelling biological evidence supports the biomarker as being predictive of response to a drug).

### 14. RNAseq-based immune and transcription factor pathways analyses

#### 14.a Tumor microenvironment (TME)

The transcriptomic profile of each tumor was used to assess the expression level in a set of 29 functional gene expression signatures representing the functional pathways or cell-type composition (immune, stromal, and other cellular populations) of the tumor micro-environment (PMID: 34019806). The list of genes defining each of these signatures was downloaded from the Github repository of the paper^14^. The ssGSEA activity score of each signature was computed from the gene-level TPM tables (Kallisto-TxImport pipeline, see section 3) using the Python function “ssgsea_formula” defined in the Github. These scores were then used to classify tumors into one of the four tumor TME subtypes defined in the paper.

In order to assign a TME to a new transcriptomic profile, we trained a model using TCGA scores and labels provided by the authors in the Github^15^. To begin with, we checked that we were able to reproduce the normalized signature scores for the same TCGA dataset as used by Bagaev et al. For that purpose, signature scores of TCGA samples were recomputed starting from the TPM expression tables (see section 15.b.5) and were then median-centered and scaled by the mean-absolute deviation independently for each tumor, mirroring the method described in the paper. Normalized scores were compared to the scores provided by the authors and showed very high concordance, as depicted in Supplementary Methods Fig. 3.

We next trained a multiclass classification model on TCGA data to predict to which TME each sample belongs using the 29 normalized scores as predictors. We compared the accuracy of different machine learning models trained on the normalized scores recomputed by us and the normalized scores provided in the paper. Best hyperparameters were fit using a five-fold cross-validation procedure and the scores reported in the table below represent the average of the five test scores from internal cross-validation splits.

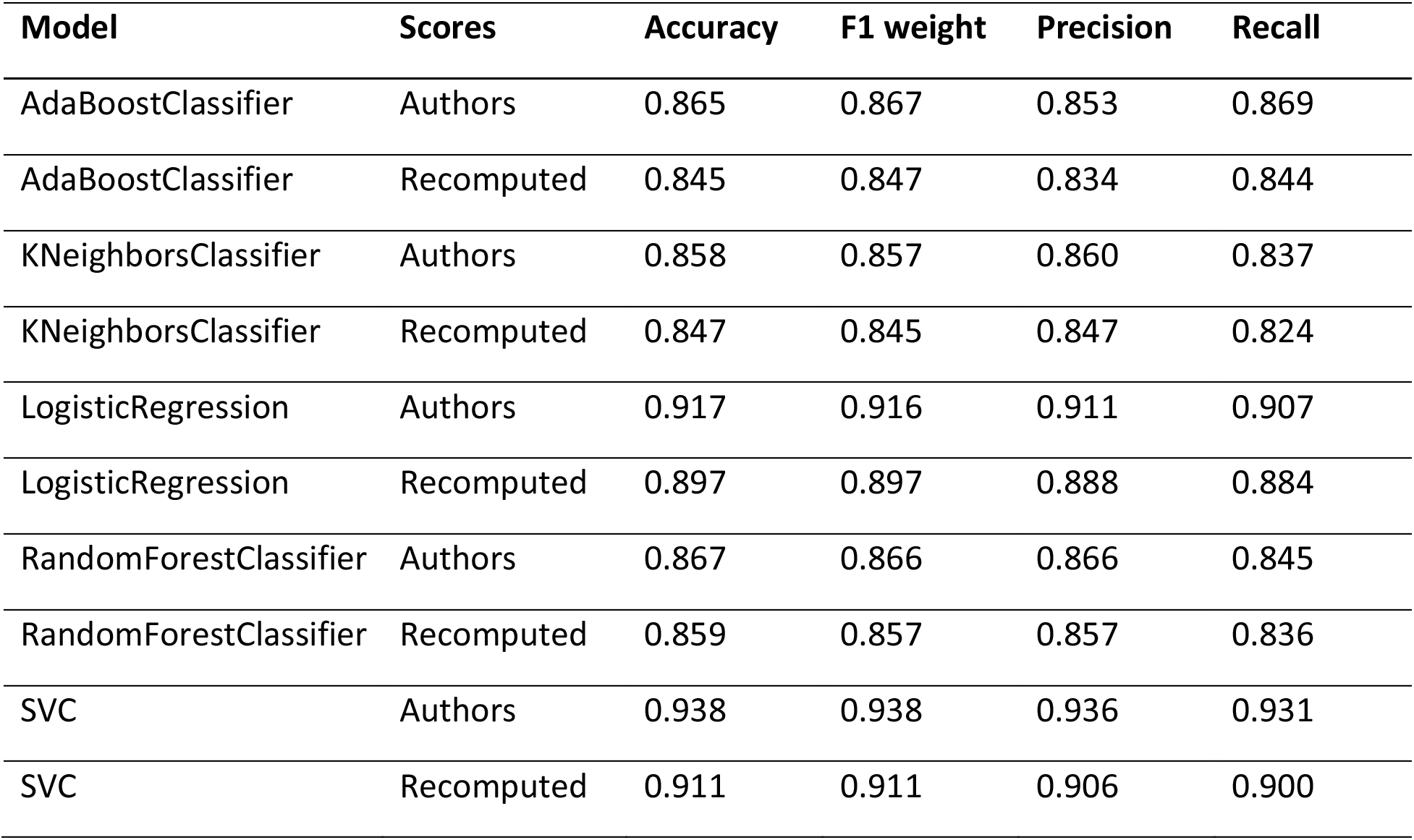

| **Model** | **Scores** | **Accuracy** | **F1 weight** | **Precision** | **Recall** |
| --- | --- | --- | --- | --- | --- |
| AdaBoostClassifier | Authors | 0.865 | 0.867 | 0.853 | 0.869 |
| AdaBoostClassifier | Recomputed | 0.845 | 0.847 | 0.834 | 0.844 |
| KNeighborsClassifier | Authors | 0.858 | 0.857 | 0.860 | 0.837 |
| KNeighborsClassifier | Recomputed | 0.847 | 0.845 | 0.847 | 0.824 |
| LogisticRegression | Authors | 0.917 | 0.916 | 0.911 | 0.907 |
| LogisticRegression | Recomputed | 0.897 | 0.897 | 0.888 | 0.884 |
| RandomForestClassifier | Authors | 0.867 | 0.866 | 0.866 | 0.845 |
| RandomForestClassifier | Recomputed | 0.859 | 0.857 | 0.857 | 0.836 |
| SVC | Authors | 0.938 | 0.938 | 0.936 | 0.931 |
| SVC | Recomputed | 0.911 | 0.911 | 0.906 | 0.900 |

Overall, we saw a very small decrease in classification performance when substituting the scores computed by the authors with the scores recomputed by us, in line with the high concordance described above. In the Github, the authors recommend the usage of KNeighbors classifiers with k=35, but these models were the worst-performing in all five models that we have tested (with best hyperparameters [tested values ranged from 2 to 80] at k=37 and k=36 for scores provided by the authors and recomputed by us, respectively). For our analyses, we used predictions from the logistic regression model trained on the scores recomputed by us as it showed excellent performance (90% weighted F1-score) and was the easiest to interpret.

Per tumor type normalization parameters learned from the full TCGA dataset were reapplied when performing predictions for the transcriptomic profiles used in our analysis. This normalization procedure allows for the use of the model in a predictive setting and for comparison between different cohorts. For that reason, only tumor types that were analyzed by Bagaev et al. could have a TME assignment. Most notably, the SARC tumor type was excluded by the authors, and therefore we did not analyze SARC tumors, nor did we analyze rare tumors.

#### 14.b Transcription factors (TFs)

To profile the relative activity of transcription factors across the samples, we filtered out lowly-expressed genes from the raw read counts table (median number of reads less than 10 per gene) and used the TMM method of edgeR package (PMID:19910308) to normalize the counts and extract logCPM values. The logCPM values were used as input to the DoRothEA regulons package (PMID: 31340985) based on the statistical method VIPER (PMID:<u>27322546</u>) to get the relative activity of each TF in every sample as a normalized enrichment score (*nes=T*) taking into account pleiotropy of the gene activity (*pleiotropy=T*). The DoRothEA regulons is a database of direct targets of transcription factors with different levels of evidence (A-E). We used only transcription factors with at least ten targets with a high level of evidence (A, B, and C) which were annotated as important parts of oncogenic signaling pathways (*FOXA1, AR, MYC, MYCN, GLI1, GLI2, LEF1, TCF7L1, TCF7L2, TCF7, SMAD2, SMAD3, TEAD1, TEAD2, TEAD3, TEAD4, E2F1, E2F2, FOXO1, FOXO3, FOXO4, FOXO6, STAT1, STAT2, STAT3, STAT4, STAT5A, STAT5B, STAT6, ELK1, ELK4, ETS1, ETS2, FOS, JUN, RBPJ, RBPJL, NFKB, RELA, RELB, TP53, HIF1A, ARNT, MYB*) (PMID: 12360277,<u>29625050)</u>.

### 15. Publicly available cancer cohorts and data reanalysis

#### 15.a MET500 data

##### 15.a.1 Patients and samples attributes

Patients and sample attributes from the MET500 study were downloaded from the dbGaP study id phs000673.v4.p1 with permission and from the supplementary tables 1 and 2 of Robinson et al. 2017 (PMID 28783718). All relevant variables were merged into curated tables that were used for analysis.

##### 15.a.2 Somatic mutations

Raw sequencing files were downloaded with permission and processed with our internal pipelines, as described in the previous sections. Only samples included from patients in the initial MET500 article were used. A total of 106,341 somatic mutations and small indels were used for analysis after applying the filtering procedure described in section 4. The filtering that resulted in this list of somatic calls is summarized in Supplementary Methods Fig. 4.

NOTE: the “OFF_TARGETS_INTERSECTION” does not appear in the Supplementary Methods Fig. 4 because for these samples, the bed file containing only the positions in the intersection of all capture kits used on META-PRISM samples was provided as input to the --intervals parameter of Mutect2.

##### 15.a.3 Germline mutations

Raw sequencing files were processed with our internal pipelines, as described in the previous sections. A total of 71 germline cancer-predisposing variants were detected in the 500 MET500 samples after applying the filtering procedure described in section 10.

##### 15.a.4 Somatic copy-number alterations (SCNAs)

Raw sequencing files were processed with our internal pipelines and SCNAs were identified using our internal SCNA pipeline.

##### 15.a.5 Gene expressions

RNA-seq data files were downloaded for 497 samples. The majority of samples had multiple files available produced from either polyA+ selection, hybridization capture, or both. As all RNA-seq files were produced from polyA+ in META-PRISM, we prioritized polyA+ whenever possible, resulting in 386 polyA+ and 111 hybridization capture RNA-seq libraries.

Gene expressions were quantified using the Kallisto + Tximport pipeline as used on META-PRISM samples.

##### 15.a.6 Gene fusions

Raw sequencing files were processed with our internal pipelines and gene fusions were identified using our internal gene fusion pipeline. A total of 731 gene fusions (disregarding breakpoints, see Supplementary Methods Fig. 5) were detected in 308 out of 497 samples with RNAseq data and successfully processed by all fusion-calling algorithms.

#### 15.b TCGA data

##### 15.b.1 Patients and samples attributes

Patients and samples attributes for the TCGA study were downloaded from the GDC data portal using the R package GenomicDataCommons and from supplementary tables publicly available on the PanCanAltas page https://gdc.cancer.gov/about-data/publications/pancanatlas. In short, attributes from all 11,315 patient samples and all 34,815 aliquots (23,346 whole-exome and 11,469 RNA-seq) of interest for our study were downloaded. Considering the data that was available to us and the latest update of patients’ annotations, a total of 554 patients were excluded for the following reasons:

- 155 patients for which we had no associated molecular data.
- 392 patients for which annotations in the aliquot attributes files or in merged_sample_quality_annotations.tsv file (available at https://gdc.cancer.gov/about-data/publications/pancanatlas) provided a basis for exclusion.

- Unacceptable prior treatment: 216
- Excluded upon AWG pathology review: 126
- Does not meet study protocol: 20
- Does not meet study protocol & Excluded upon AWG pathology review: 14
- Subject withdrew consent: 4
- Duplicated subject: 4
- Qualified in error: 3
- Qualified in error & Excluded upon AWG pathology review: 3
- Unacceptable prior treatment & Excluded upon AWG pathology review: 1
- Genotype/gender mismatch: 1
- 7 patients for which we both had no molecular data and annotations providing a basis for exclusion.

A total of 10,761 patients for which we had one or multiple types of molecular data available and found no reason for exclusion were considered for analysis.

##### 15.b.2 Somatic mutations

TCGA somatic mutation catalog was taken from the controlled-access MAF file mc3.v0.2.8.CONTROLLED.maf.gz available at https://gdc.cancer.gov/about-data/publications/pancanatlas. All filters described in the “FILTER” column were applied as well as the following additional filters:

- Minimum VAF of 5% (LOW_VAF).
- Minimum sequencing coverage of 20X in the tumor sample (LOW_COVERAGE_TUMOR).
- Minimum sequencing coverage of 10X in the normal sample (LOW_COVERAGE_NORMAL).
- Localized within the META-PRISM target region (OFF_TARGETS_INTERSECTION).
- Located inside exonic regions as defined by the set of canonical transcripts used by VEP v104 on GRCh37 assembly (NOT_EXONIC).
- Allele frequency across all gnomAD v2.1 exome subpopulations is < 0.04% (COMMON_VARIANT). This rule is not applied for variants annotated by OncoKB (see section 12).
- Only SNPs and MNPs mutations identified by at least two of the five callers used by the MC3 consortium were retained. Likewise, only INDELs identified by Indelocator or Varscan (INDELOCATOR or VARSCANI tags in the “CENTERS” column from the controlled-access MAF file) were retained (INDEL/SNP_CALLING_ALIGNMENT).

These filters were determined after running our internal pipeline on a test set of 58 TCGA raw whole-exome sequencing files downloaded with permission from the GDC data portal. Agreement results from the comparison of the variants detected using our pipeline with the variants reported in the mc3.v0.2.8.CONTROLLED.maf.gz file after applying the set of optimal filters are summarized in the table below, where DSC stands for Dice-Sorensen coefficient, TP true-positives, and FP false-positives.

| Mutations | CENTERS | DSC | TP/# True | FP/# True | # Pred | # True |
| --- | --- | --- | --- | --- | --- | --- |
| SNPs/MNPs | 2 or more | 0.90 | 0,91 | 0,121 | 5686 | 5860 |
|  | INDELOCATOR |  |  |  |  |  |
| INDELs | or VARSCANI | 0.84 | 0,864 | 0,193 | 228 | 241 |

True mutations designate mutations from the MC3 MAF file. DSCs of 0.89 for SNPs/MNPs and 0.84 for INDELs indicate very good agreement. Supplementary Methods Fig. 6 (resp. Fig. 7) provide details about the number of predicted SNP (resp. INDEL) vs true SNP (resp. INDEL) for each of the 58 samples analyzed.

A total of 2,097,573 somatic mutations and small indels were used for analysis in the 8,974 patients for which somatic mutation data was available out of the 10,761 patients considered for analysis. The filtering that resulted in this list of somatic calls is summarized in Supplementary Methods Fig. 8. All PASS mutations from the refiltered MC3 MAF file were split into individual VCF files for each tumor/normal pair and were subsequently annotated using the same annotation pipeline as used on META-PRISM data (see section 4).

##### 15.b.3 Germline mutations

The list of cancer-risk germline variants detected in TCGA samples was taken from the file PCA_pathVar_integrated_filtered_adjusted.tsv available at https://gdc.cancer.gov/about-data/publications/PanCanAtlas-Germline-AWG (PMID: 29625052). Reducing the original list of 1,393 mutations to only mutations in the 10,761 patients considered for analysis results in 1,342 pathogenic germline variants detected in 1,253 patients. After running the filtering procedure described in section 10, 828/1,253 variants were retained and used for analysis.

##### 15.b.4 Somatic copy-number alterations (SCNAs)

SCNAs in TCGA were identified through a reanalysis of all available TCGA WES files (21,987 BAM files from 10,332 patients with gender available and at least one pair tumor/normal WES files; 12,129 pairs in total) by FACETS tool (PMID: 27270079). This reanalysis was a necessary step to avoid large batch effects arising when comparing data generated from different copy-number variation assessment techniques. Execution of FACETS and SCNA calling from segments and allele-specific copy numbers were performed via the Google Cloud Engine with support from Institute for Systems Biology Cancer Genomics Cloud (ISB-CGC).

SCNA categories for TCGA data were identified using the exact same rules as used for META-PRISM samples (see section 6).

##### 15.b.5 Gene expressions

Gene and transcript expression tables for all TCGA RNAseq samples were retrieved from the supplementary data of Zheng et al. 2019 (PMID: 31808800) available at https://stanfordmedicine.app.box.com/s/lu703xuaulfz02vgd2lunxnvt4mfvo3q. In short, these tables were obtained by running a Kallisto + Tximport (Gencode v27) pipeline^16^ on TCGA raw sequencing files. This pipeline is identical to the pipeline that we have employed on META-PRISM samples, thereby minimizing technical differences between the cohorts.

##### 15.b.6 Gene fusions

Three independent and publicly available lists of TCGA gene fusions were retrieved from the following sources:

- Hu et al. 2018 (PRADA, PMID: 29099951) supplementary table nar-02671-data-e-2017-File007.xlsx.
- Gao et al. 2018 (StarFusion, PMID: 29617662) supplementary table S1.
- Dehghannasiri et al. 2019 (DEEPEST, PMID: 31308241) supplementary table pnas.1900391115.sd01.xlsx.

In order to adjust the filtering criteria and compare the nf-core fusion pipeline (see section 8) used for META-PRISM fusions to the three different pipelines used for the aforementioned lists of TCGA fusions, we downloaded FASTQ files for 69 TCGA samples from the GDC data portal and analyzed them using the nf-core fusion pipeline. All 69 samples were successfully processed by the six different callers available in nf-core fusion pipeline, namely: Arriba (“AR”, v1.2.0), EricScript (“ES”, v0.5.5), FusionCatcher (“FC”, v1.20), Pizzly (“PZ”, v0.37.3), Squid (“SQ”, v1.5), StarFusion (“SF”, v1.8.1). All detected fusions were then annotated with FusionAnnotator^17^ that connects datasets of fusions detected in cancer or normal tissues^18^.

Supplementary Methods Fig. 9 summarizes the overlap between the fusions detected by these six algorithms. The fusions predicted by Squid were very different from that predicted by any of the five other callers (see figure above) and were therefore removed from all analysis. Additionally, even though FusionCatcher was successful on all 69 TCGA samples, it repeatedly failed on some META-PRISM samples and was therefore not considered further. Consequently, only fusion calls from Arriba, EricScript, Pizzly, and StarFusion were used.

As our study only aimed at describing the variations relevant to cancer, only fusions known in cancer or involving at least one oncogenic partner (see section 12) were analyzed. We, therefore, limited the lists of fusions as detailed hereafter.

Firstly, we removed all fusions that have been previously reported in studies of normal tissues. More specifically, fusions were removed if they met any of the following criteria:

- are in Babiceanu_Normal list (PMID: 26837576).
- are in ChimerSeq_Normal_v4.0 list (available upon request to the authors, PMID: 31680157), which was established from the analysis of 1,144 TCGA normal samples and curated in order to remove well-known fusions (e.g., TMPRSS2-ERG) sometimes seen in normal samples.
- are in GTEX_V6 supplementary table S3 (PMID: 31965184).
- have at least one “Red Herring” flag (FusionAnnotator annotations) among the following: GTEx_recurrent_StarF2019, BodyMap, DGD_PARALOGS, HGNC_GENEFAM, Greger_Normal, ConjoinG.
- one of the partners is not protein-coding.

Secondly, only fusions that met one of the following criteria were retained:

- are in COSMIC v95 list of fusions^19^.
- are in ChimerKB v4.0 list (PMID: 31680157).
- are in Chitars Cancer v5.0 list (PMID: 31747015, available upon request to the authors).
- are in TIC v3.3 list^20^.
- one of the partners is a cancer driver (see section 12).

Lastly, after filtering the fusions calls for the 69 samples in each of the three TCGA published lists and in each of the lists of fusions predicted by the four callers considered, we looked for the combinations of calls that showed the best agreement. The best agreement was obtained between the following combinations:

- for the three published TCGA lists: fusions seen by StarFusion or by both DEEPEST and PRADA.
- for the four callers in our pipeline: fusions seen by both Arriba and EricScript or by both Pizzly and StarFusion.

Agreement results are summarized in the table below, where DSC stands for Dice-Sorensen coefficient, TP true-positives, and FP false-positives.

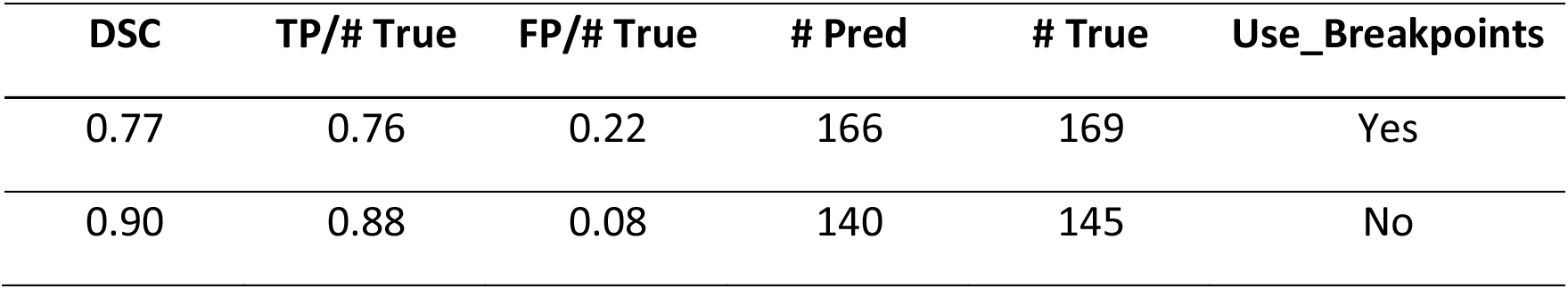

| DSC | TP/# True | FP/# True | # Pred | # True | Use_Breakpoints |
| --- | --- | --- | --- | --- | --- |
| 0.77 | 0.76 | 0.22 | 166 | 169 | Yes |
| 0.90 | 0.88 | 0.08 | 140 | 145 | No |

True fusions designate fusions from the combination of published TCGA lists. The DSC was 0.90 if fusion calls were assessed to be concordant regardless of the breakpoint prediction and was 0.80 if the predicted breakpoint was required to be identical.

To summarize, we kept for TCGA the fusions seen by StarFusion alone (8,194 fusions), by DEEPEST and PRADA exactly (2,604 fusions), or by all three methods (9,989 fusions). Supplementary Methods Fig. 10 represents the overlap between these three gene fusion lists and highlights in red the fusions that we have retained.

#### 15.c ExAC data

ExAC release 1.0 vcf file was downloaded from the gnomAD download page^21^ and annotated with ANNOVAR using the same pipeline as used for META-PRISM vcf files. The same filtering procedure as described in section 10 was applied, resulting in 2,002 cancer-predisposing germline variants among the Non-Finnish European population.

### 16. Tumor purity

The tumor purity of DNA samples for META-PRISM, MET500, and TCGA tumors was estimated using FACETS. We observed a very good tumor purity in all three cohorts, with an average purity above 50% in each of them. Moreover, the average tumor purity of META-PRISM samples was lower than that of TCGA, which rules out the possibility that the observed increased variation in META-PRISM is due to better tumor purity.

### 17. Survival analyses

#### 17.a Univariate analyses

All univariate analyses are based on Kaplan-Meier estimates comparing two or more groups of patients defined by values of a categorical variable. The survival curves and log-rank tests comparing these groups of patients were all obtained using the survminer R package. Univariate survival analyses were applied to assess the association with survival of

1. the most frequently altered driver genes (only genes altered in at least 12 patients if considering only one tumor type, 20 patients otherwise)
2. the tumor microenvironment subtypes
3. the number of metastatic sites and the number of resistant drugs before biopsy
4. the GRIm score

in the five most represented tumor types in our cohort (BLCA, BRCA, LUAD, PAAD, PRAD) and in the META-PRISM WES & RNAseq cohort (ten tumor types).

#### 17.b Multivariate analyses

All multivariate analyses are based on Cox multivariate regressions incorporating parts of all possible covariates depending on the effects that we were trying to measure. To provide a comparative assessment of the added and independent value of biomarkers derived from WES or RNAseq as compared to clinical biomarkers (age, gender, global tumor characteristics, blood test results, treatment history), we experimented with multiple models that incorporated different combinations of predictors. The baseline model (M2) to be improved upon was composed of all standard clinical variables plus the continuous components of the GRIm score (lactate dehydrogenase, albumin, neutrophil, and lymphocyte levels). A separate model was run for each of the five most represented tumor types in our cohort (BLCA, BRCA, LUAD, PAAD, PRAD) and the META-PRISM WES & RNAseq cohort (ten tumor types). In order to ensure a fair comparison with more complex models, only samples with complete molecular profiles (WES & RNAseq) were used in the different Cox models.

For each combination of predictors and samples, we repeated the feature selection and coefficients estimation steps 1,000 times to assess the selection procedure’s stability, if any, and to provide robust estimates and confidence intervals for the effect of each variable on survival. Each of the 1,000 repeats consisted in running the selection procedure, if any, and training the model on a random 80% subsample. The remaining 20% were used to assess the model quality (C-index). The C-indices values reported in Figure 7B are averages of the 1,000 estimates of the C-index on the 20% test subsamples. Estimates of the covariates’ coefficients were computed by averaging the coefficient fitted on each 80% subsample across the 1,000 repeats. In case a selection procedure is active at each repeat, covariates were assigned an estimated coefficient of zero every time the selection procedure did not retain them. Confidence intervals at the 95% level were estimated by computing the empirical 2.5% and 97.5% quantiles from the 1,000 estimates.

A selection procedure was applied in case the number of covariates was unreasonably high given the number of available observations. We experimented with univariate – fit univariate Cox model for each candidate predictor and select only predictors for which adjusted p-values fall below a certain threshold - and multivariate selection procedures – fit Cox model with lasso penalization and select only predictors with non-zero coefficients to identify a small set of predictors that, hopefully, is a superset of all important predictors. The Cox regressions on predictors selected by the lasso were always more prognostic (higher value of C-index) than when predictors were selected through the univariate procedure.

Each set of 1,000 repeats of Cox regressions was preceded by preprocessing steps (run once for each combination of samples and features) in which covariates were formatted, imputed, min-max transformed, and analyzed for redundancy. The imputation relied on the MICE R package (PMID: 26889483) that performs multiple imputations of the same dataset by iteratively learning a predictive model of each covariate with missing data from all other covariates and randomly sampling from observed values through the predictive mean matching procedure. Data tables with at least one missing value were imputed five times, and coefficient estimates across each of the 1,000 repeats were calculated by averaging estimates across the five imputed tables.

The C-index was estimated in two different ways, either using the C-statistic proposed by Harrell et al. (PMID: 8668867) or using the inverse probability weighting estimate presented by Uno et al. (PMID: 21484848) to account for the fact that the C-statistic proposed by Harrell et al. depends on the censoring distribution which in practice is rarely independent of the covariates used in the Cox regression. As survival times are subject to censoring, and because we were mainly interested in discriminating between patients having a survival longer or shorter than six months, we used an estimate of the C-index truncated at six months by considering only pairs of patients for which one of the two patients had a survival shorter than six months. The estimator proposed by Uno et al. (see formula 2.3), as well as the C-statistic proposed by Harrell et al., were implemented in C using a truncation time of six months. A comparison of the estimated values from both estimators showed very little difference due to the fact that only a minority of patients were right-censored in our cohort (< 10%). Values reported in the main text use the C-statistic from Harrell et al. truncated at six months.

### 18. Statistical tests

For each analysis where the frequency of a binary event was compared at the cohort level, we used the stratified Cochran-Mantel-Haenszel test stratifying by tumor type to control for different tumor type composition between the cohorts or a simple Fisher exact test for the specific case where the tumor type compositions were already aligned (whole-genome duplication frequency at cohort level, Fig 3a). In cases where we analyzed the distribution of a continuous variable, the cohort-level comparison was made through a standard Mann-Whitney U test, regardless of the tumor type composition.

For analyses performed for each individual tumor type, we used the Fisher-Boschloo test when comparing the frequency of a binary event (e.g., chromosome arm gains/losses, whole-genome duplication status, mutation status, etc.) and the Mann-Whitney U test when comparing the medians of a continuous variable (tumor mutational burden, number of driver events, number of mutations contributed by each mutational signature, genome fraction covered by each type of SCNA).

Log odd ratios estimates and confidence intervals measuring the associations between one or multiple predictors and a binary outcome were computed by fitting standard logistic regression models (statsmodels python package and stats R package).

Comparisons of Kaplan-Meier survival curves were performed using the standard log-rank test implemented in the ggsurvplot and survdiff R packages. Adjusted hazard ratios were estimated by multivariate Cox regression modeling incorporating the variable of interest and the variable to adjust for (e.g., absence/presence of oncogenic alterations adjusted for tumor type in Extended Data Fig. 9).

Chromosome regions with SCNAs enriched in META-PRISM vs. TCGA480 were identified using the non-parametric method IWTomics (PMID: 29474526). The statistical significance was inferred through the comparison of the SCNA fraction distributions by 1Mb intervals. We have chosen the quantile statistics (statistics=’quantile’, probs=c(0.25,0.75)) for the IWTomic test, as the most robust to zero-inflated data. The resampling procedure used in IWTomics allows not to apply the multiple testing correction and to accurately identify the boundaries of significantly different SCNAs.

Whenever multiple independent tests were performed within a given analysis, we applied the Benjamini-Hochberg multiple testing correction controlling for the false-discovery rate and only reported adjusted p-values < 0.05 (so-called “q-values”) unless explicitly mentioned otherwise.

### 19. Code used

All the code supporting the analysis, numbers, and figures presented in this study is available at https://github.com/gustaveroussy/MetaPRISM_Cohort.

### 20. Resources and databases

A list of all external data sources and files used for this study is available in Supplementary table S11.

## Data Availability

Experimental data generated in this study have been deposited to the European Genome-Phenome Archive (EGA) under the accession number EGAD00001009684.

All the other data supporting the findings of this study are available within the article and its supplementary information files or from the corresponding authors upon reasonable request. A reporting summary for this article is available as a Supplementary Information file.

## Data Availability

Experimental data generated in this study have been deposited to the European Genome-Phenome Archive (EGA) under the accession number EGAD00001009684.
All the other data supporting the findings of this study are available within the article and its supplementary information files or from the corresponding authors upon reasonable request.

## Authors’ Contributions

MD, AH, YL, BB, and FA provided sequencing data. SN and FA designed the project. YP, PHC, DG, and SN designed the analyses. YP, JV, MD, LC, GG, LV, SM, GJC, YL, LM, BB, DG, and FA retrieved data. YP, LC, GG, and JV curated clinical data. YP, MD, IP, and LP performed data preprocessing. YP, KG, AY, AL, and DG performed NGS data analyses. YP, MD, IP, KG, DG, and LP performed bioinformatic analyses. VS performed validation experiments. YP performed survival analyses. YP and SN wrote the manuscript. JV, LC, SM, AH, LF, YL, FA, and DG commented on the manuscript.

## Acknowledgements

This work is supported by Prism – National Precision Medicine Center in Oncology funded by the France 2030 programme and the French National Research Agency (ANR) under grant number ANR-18-IBHU-0002. The authors are thankful to Fabian Seidl, Bill Longabaugh, and David Pot from the Institute for Systems Biology Cancer Genomics Cloud (ISB-CGC) for providing technical and financial support in the analysis of TCGA WES files.

## Supplementary Figures

**Figure S1.**
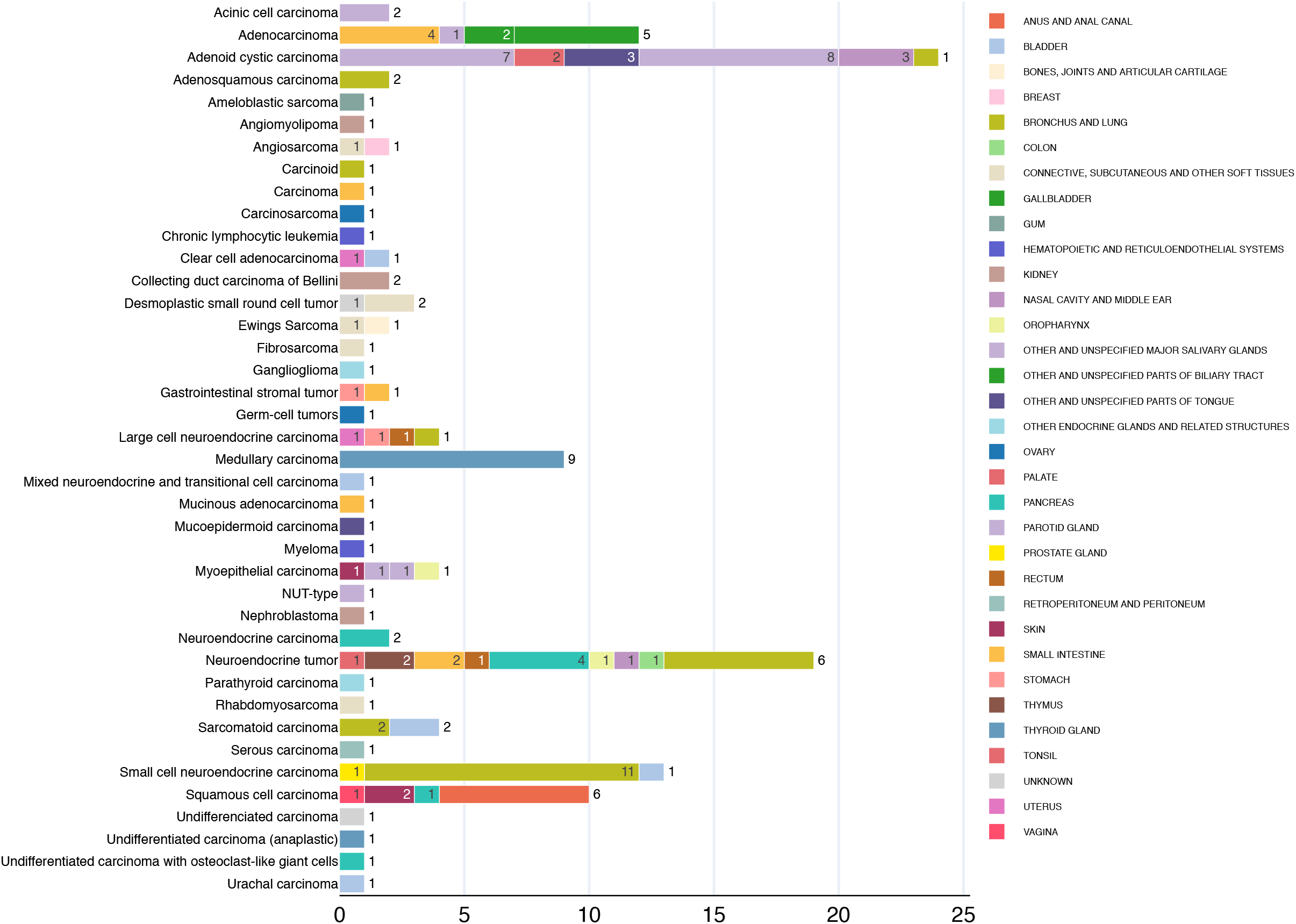
Histological subtypes and primary sites of META-PRISM rare tumors. Primary site and histological type of tumors from 140 patients having a rare tumor (not classifiable in the tumor types analyzed by TCGA). Electronic health records of all 140 patients were manually reviewed by a specialist.

**Figure S2.**
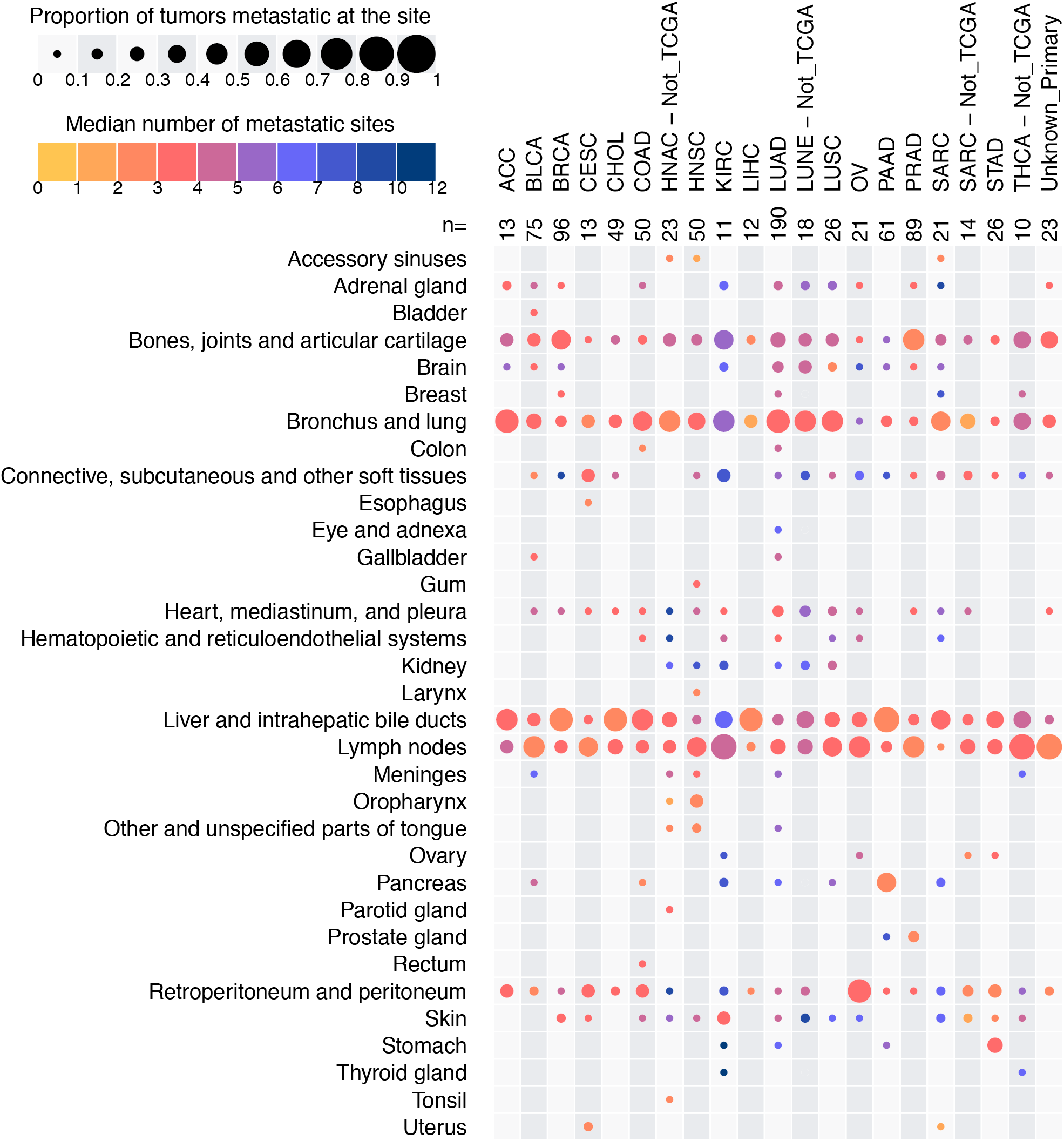
Localization and number of metastases in META-PRISM. Heatmap showing the localization of metastatic sites (rows) per tumor type (columns). The circle size encodes the percentage of patients harboring a metastasis at the corresponding site and tumor type. The circle color encodes the median number of metastases of these patients. The site of the primary tumor was added added to the list of metastases in case it was not resectable.

**Figure S3.**
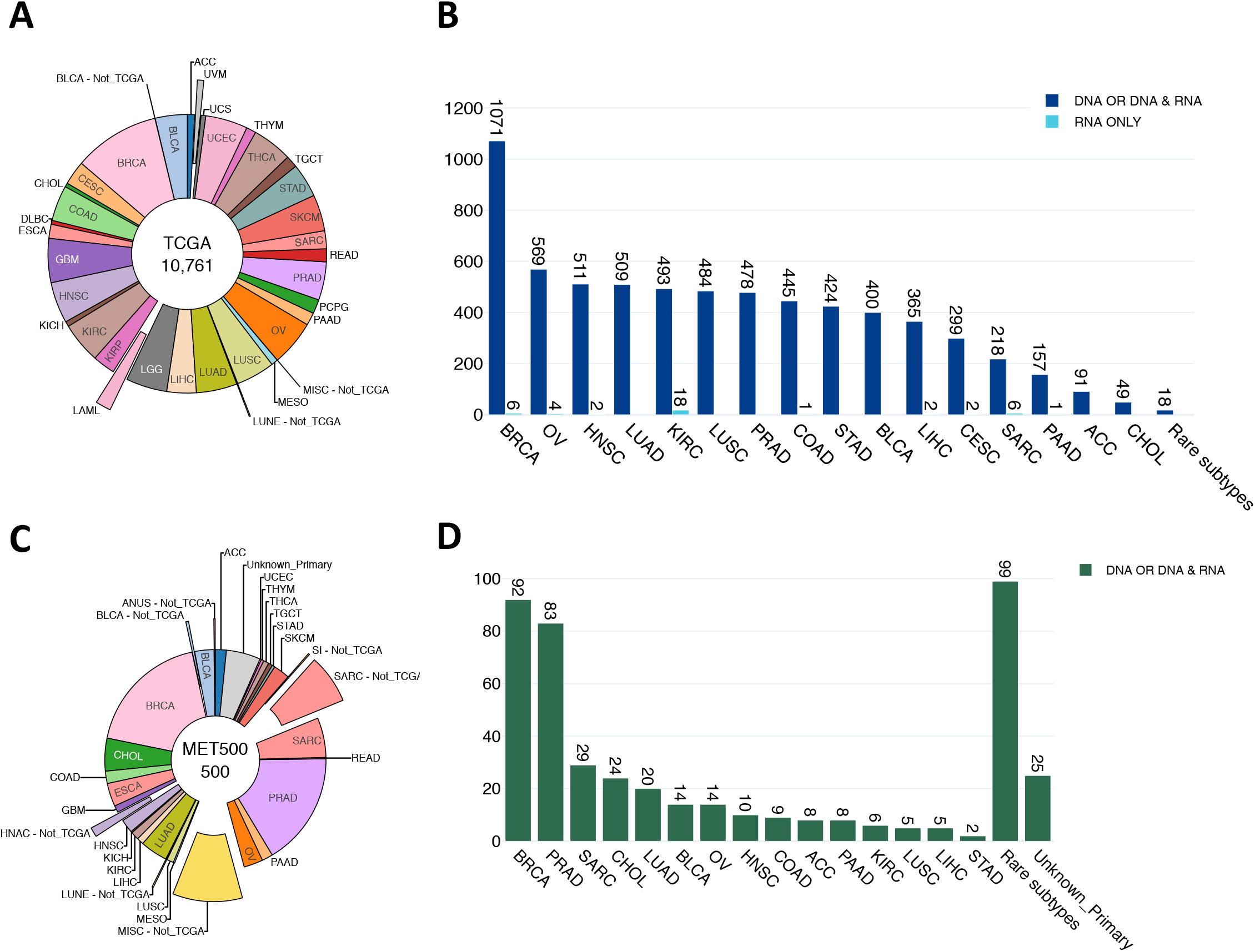
Tumor types and sample types for TCGA and MET500. **A.** Distribution of the TCGA tumors according to TCGA classification of tumor types. Tumors that could not fit into this classification were classified into tumor types suffixed by “Not_TCGA” and are shown as exploded slices as are tumor types with no representant in META-PRISM. **B**. Numbers of patients with either DNA only or DNA and RNA samples, or RNA only samples for each tumor type. Only tumor types represented by at least 10 tumors in META-PRISM are shown. **C. D.** Identical to **A. B.** but for MET500.

**Figure S4.**
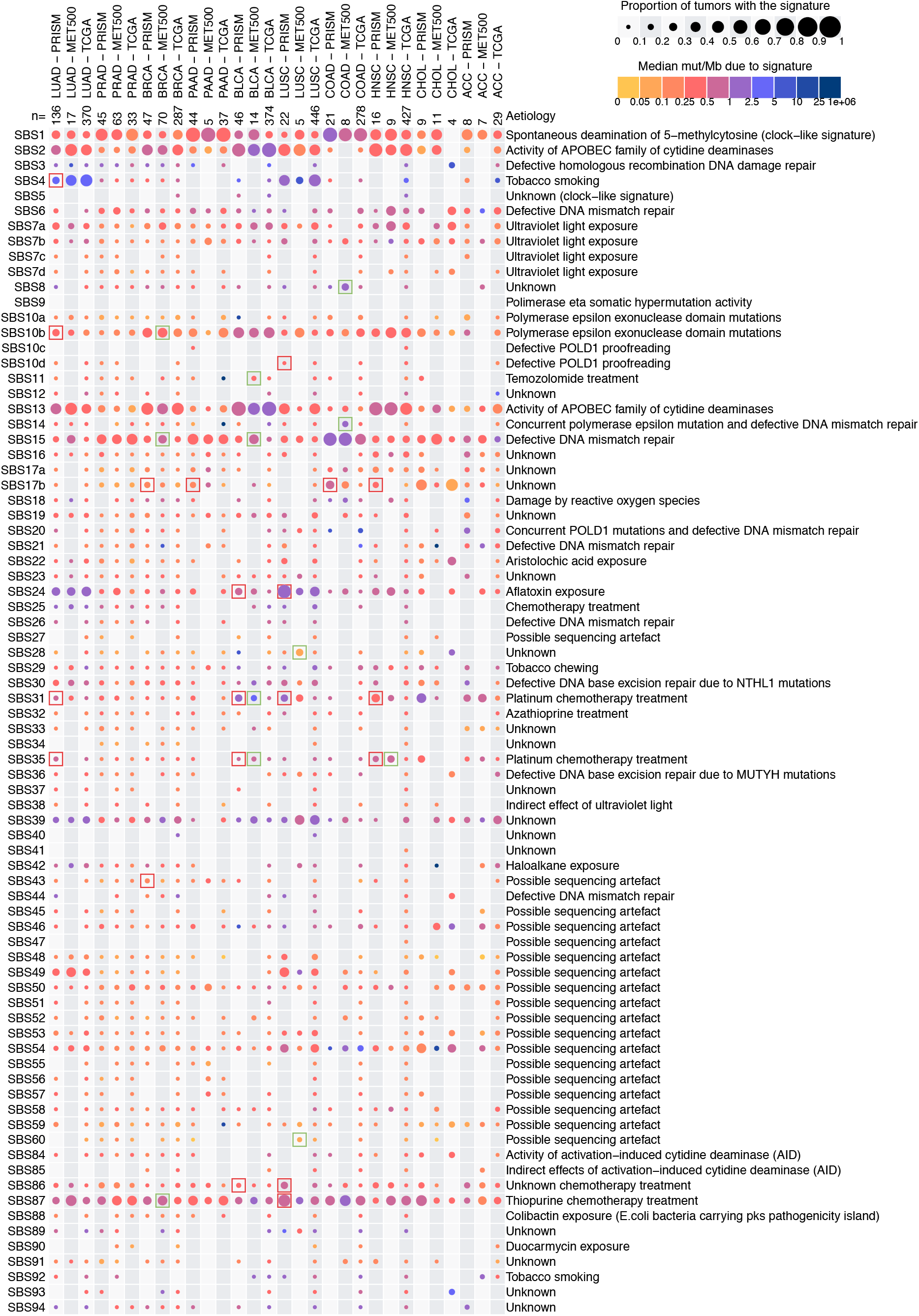
Mutational signatures in META-PRISM, MET500 and TCGA tumors. Activities of mutational signatures (rows) in each tumor type (columns) are encoded in the circle sizes and colors. The circle size is proportional to the fraction of tumors in which the signature was detected while the circle color represents the median number of mutations contributed by the signature in these tumors. Red and green frames indicate a significant difference (FDR < 0.1) between META-PRISM and TCGA or MET500 and TCGA respectively, as quantified by Mann-Whitney U tests corrected for multiple testing through the Benjamini-Hochberg procedure. Only tumors with at least 50 somatic mutations and from the ten tumors types of META-PRISM WES subcohort are displayed. Signatures are from the catalog COSMIC SBS v3.2 (78 signatures).

**Figure S5.**
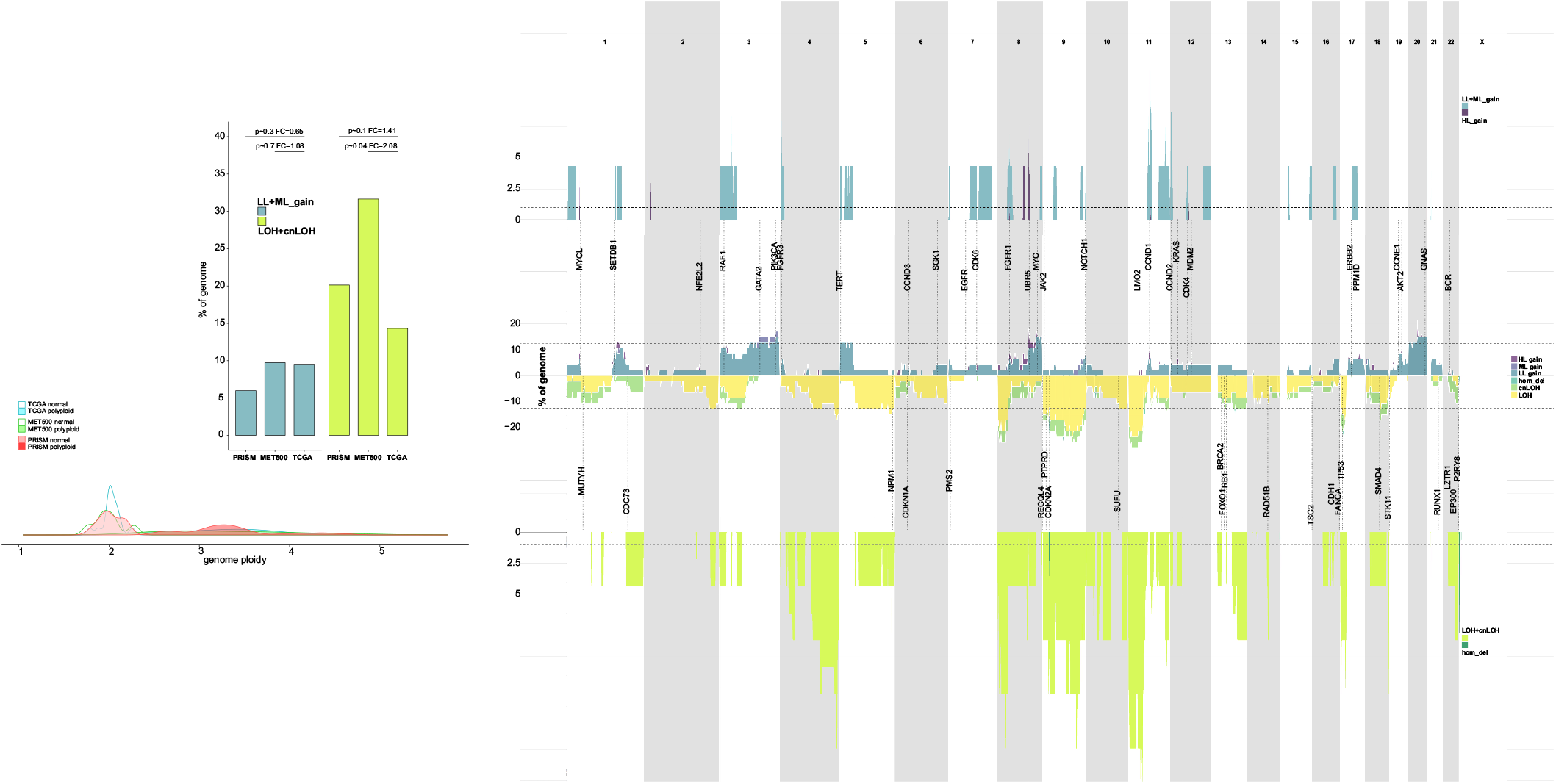
Genomic landscape of somatic copy-number alterations in BLCA tumors.

**Figure S6.**
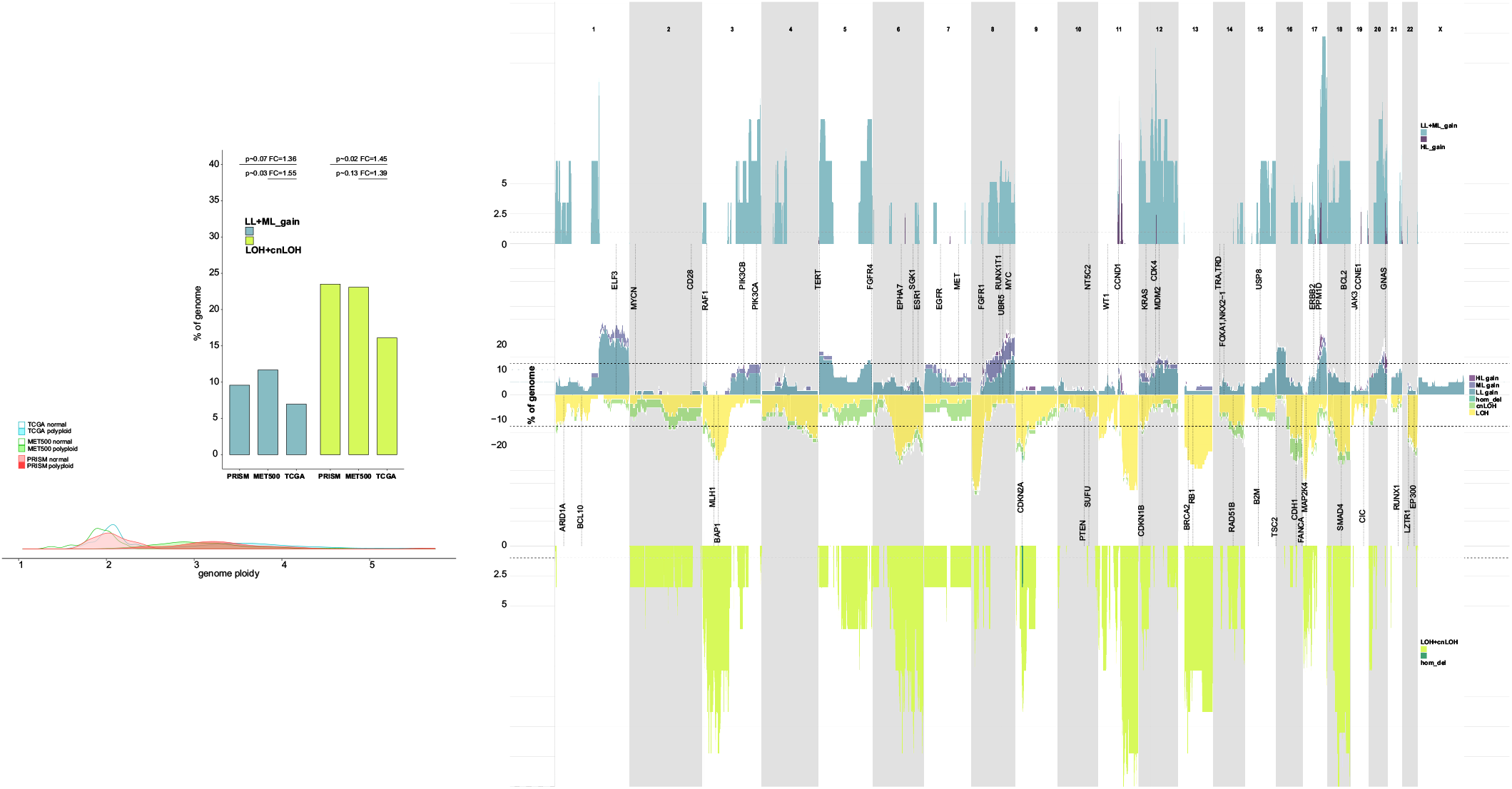
Genomic landscape of somatic copy-number alterations in BRCA tumors.

**Figure S7.**
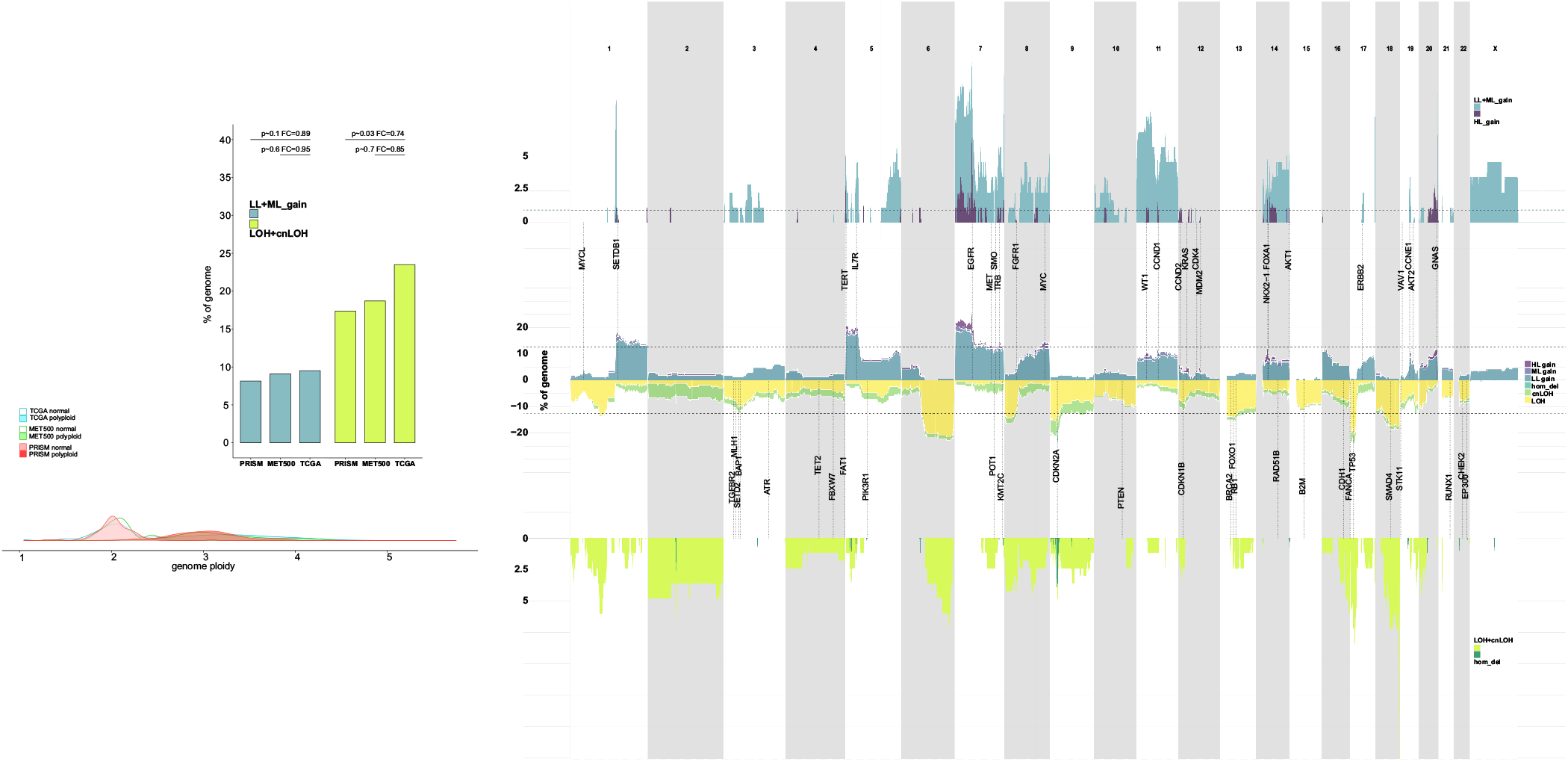
Genomic landscape of somatic copy-number alterations in LUAD tumors.

**Figure S8.**
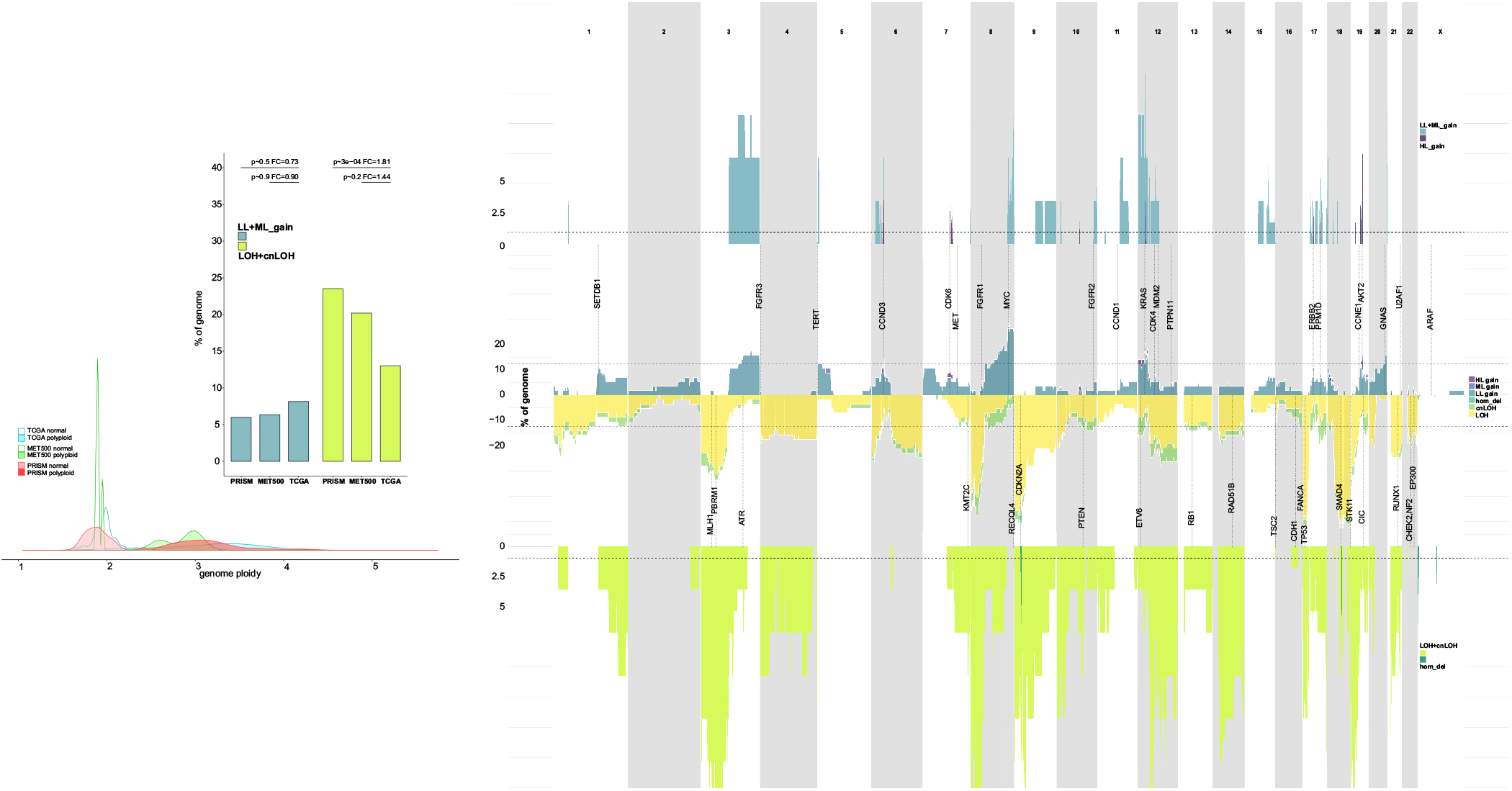
Genomic landscape of somatic copy-number alterations in PAAD tumors.

**Figure S9.**
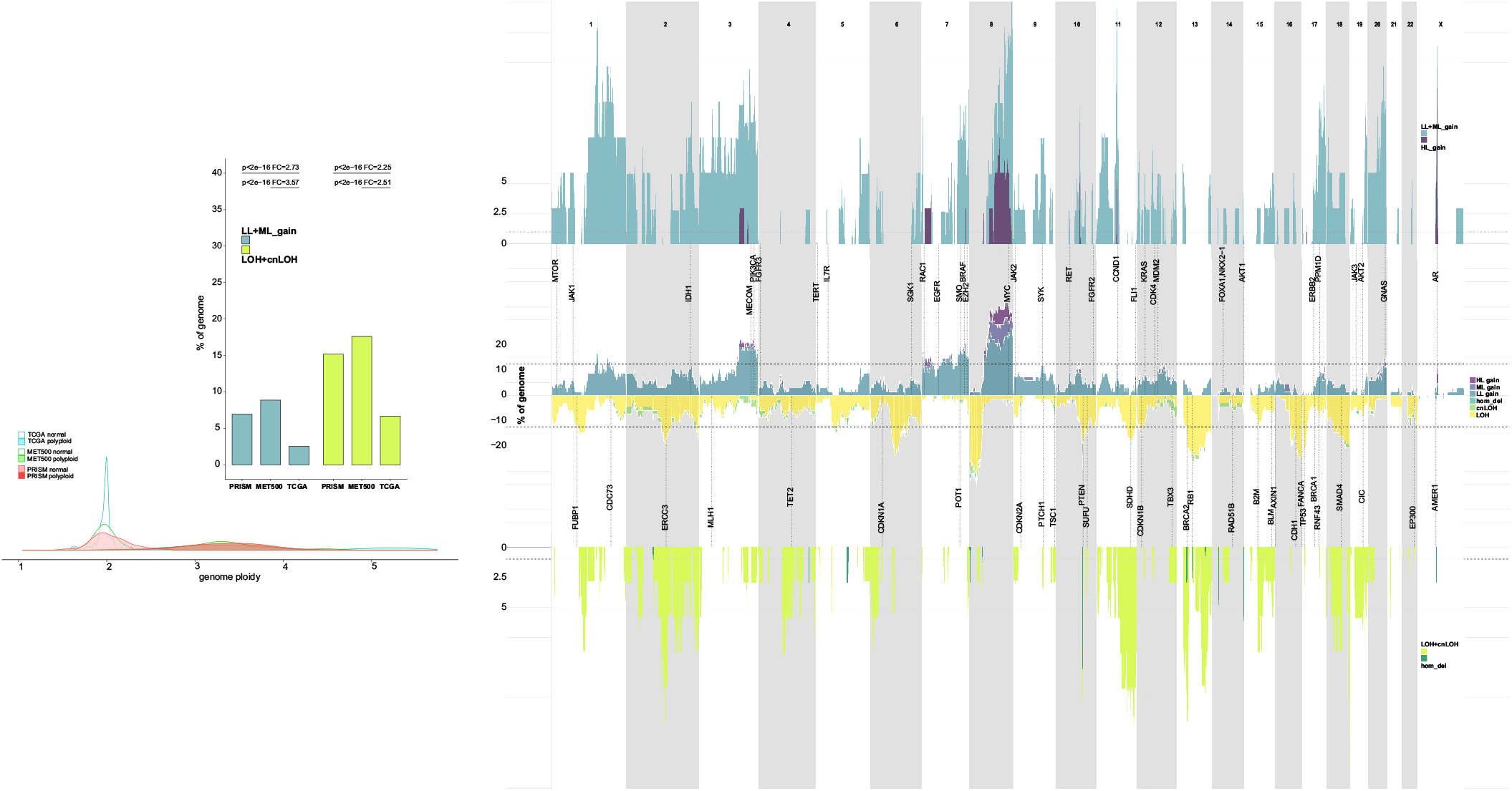
Genomic landscape of somatic copy-number alterations in PRAD tumors.

**Figure S10.**
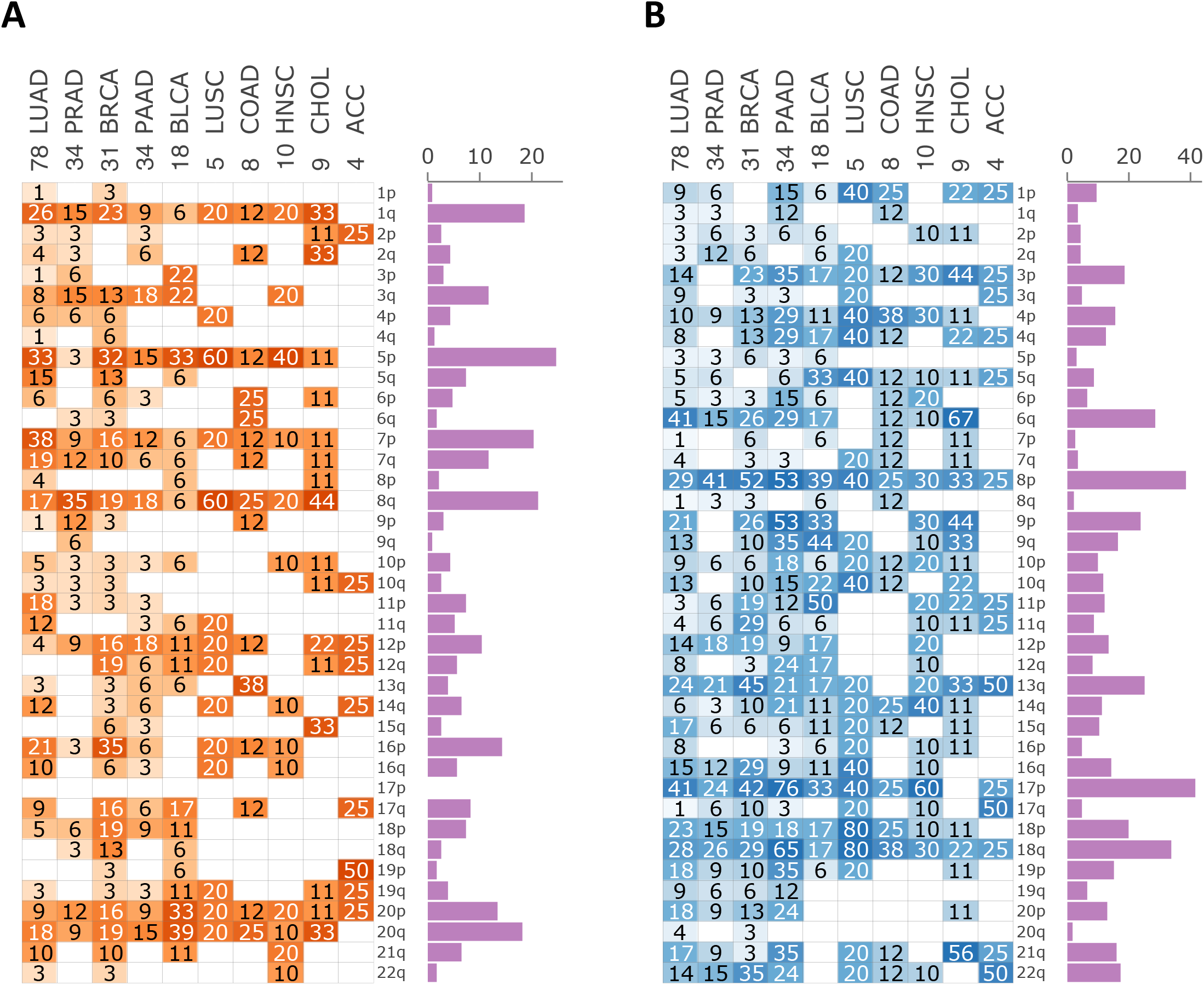
The landscape of chromosome arm SCNAs in non-WGD META-PRISM tumors. Chromosome arm **A.** copy-gains and **B.** copy-losses in the tumor types of META-PRISM WES subcohort (10 tumor types). Heatmaps show the percentages of tumors affected by corresponding chromosome arm SCNAs in each tumor type. The absolute bar plots show the percentage of tumors in META-PRISM harboring the chromosome arm SCNA. After correction of p-values from Fisher-Boschloo tests, there was no significant change in any of the chromosome arms and tumor types.

**Figure S11.**
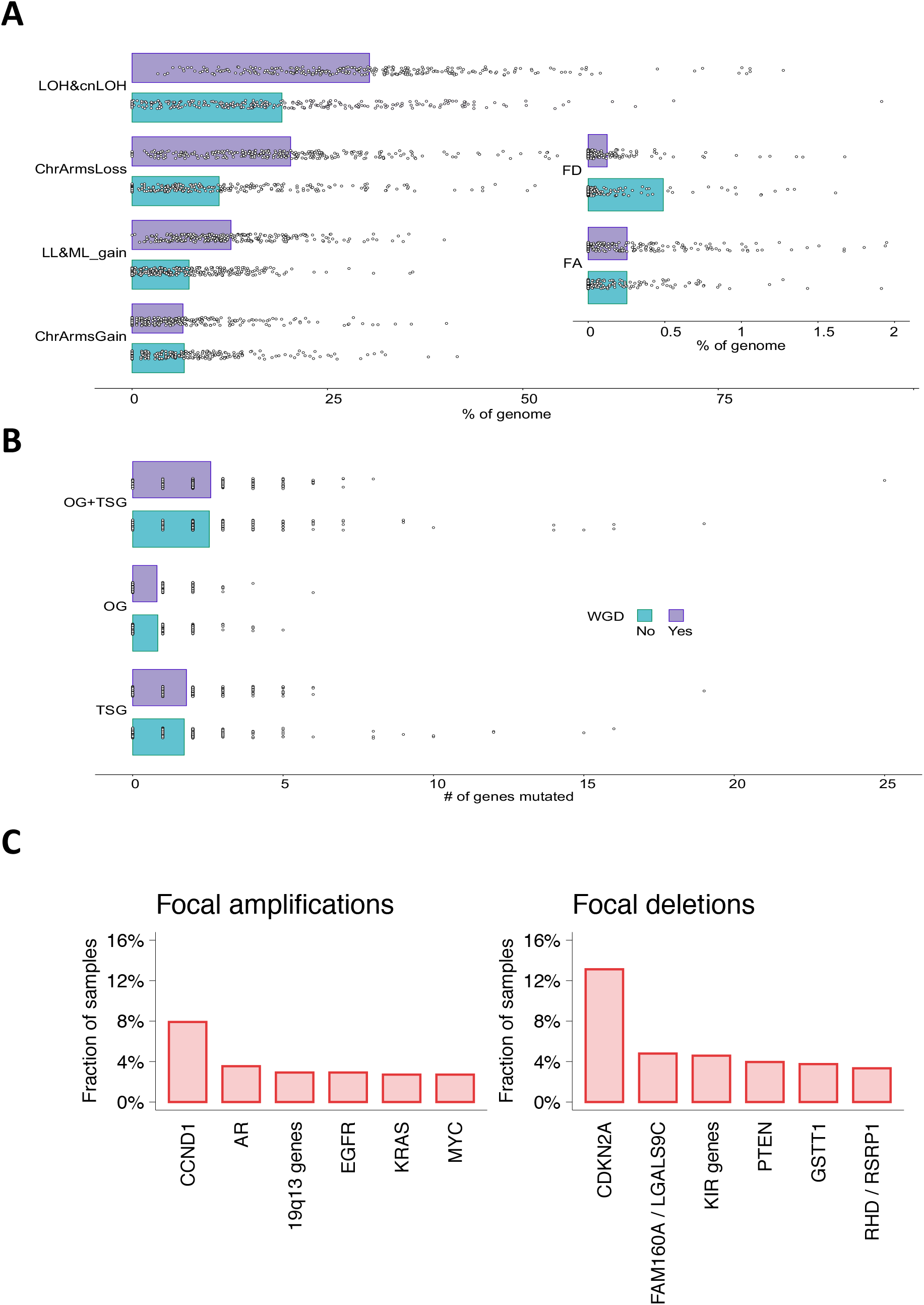
SCNAs and gene mutations according to WGD status and most frequently amplified/deleted genes in META-PRISM WES cohort. **A.** Bar plot of the fraction of the genome covered by different types of SCNAs in META-PRISM WES tumors according to the WGD status. **B.** Similar to **A.** but consideringthe number of driver oncogenes (OG) and tumorsuppressor genes (TSG) altered. **C.** Fraction of META-PRISM WES tumors harboring high-level gains (left) and homozygous deletions (right) for the genes most frequently involved in SCNAs. Only genes or gene groups altered in at least 2.5% of samples are shown.

**Figure S12.**
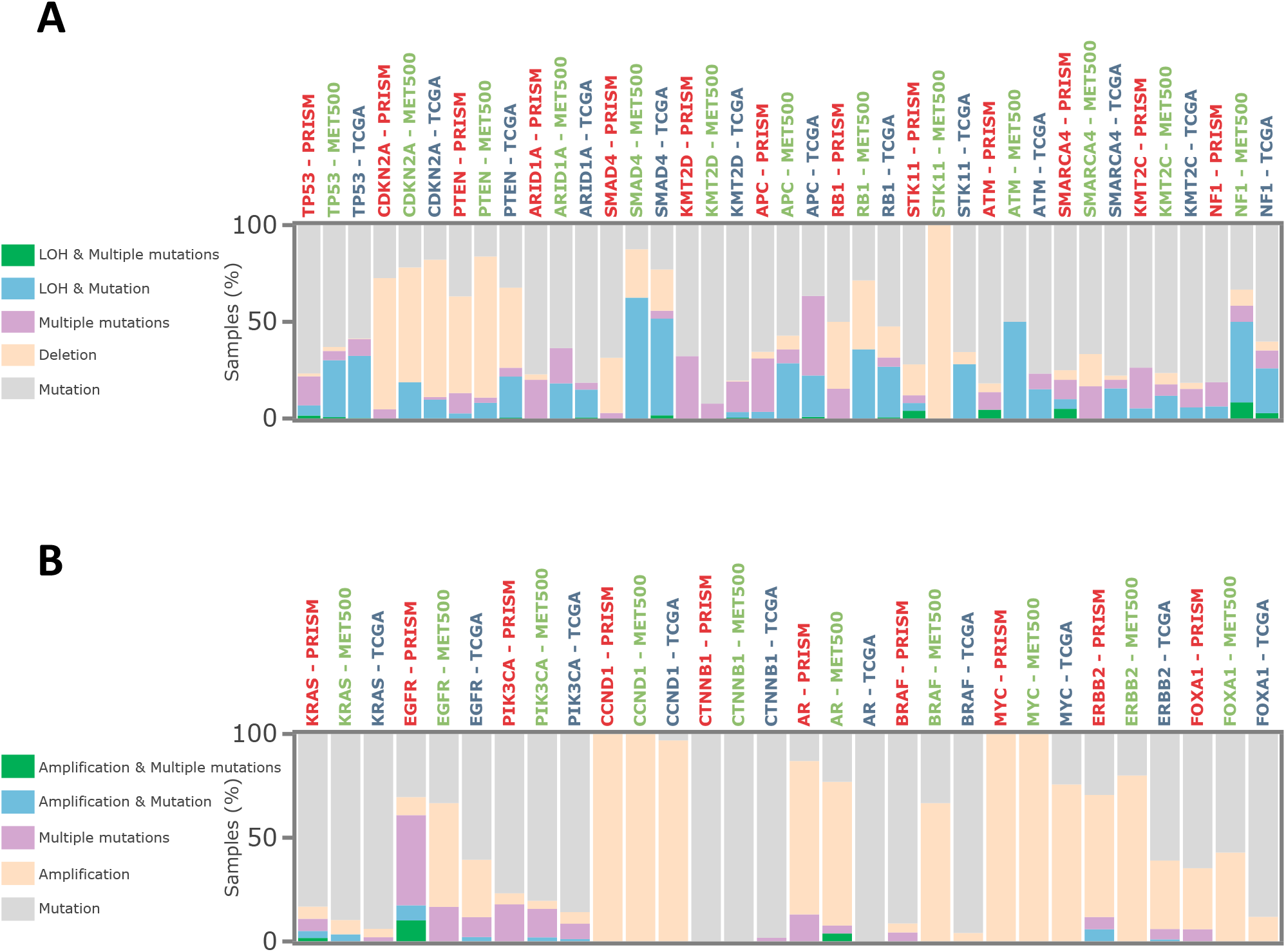
Multi-hit events in main driver genes in META-PRISM WES tumors. Relative bar plots showing the relative frequency of single-hit and different types of multi-hit events in the main **A.** tumor suppressor genes and **B.** oncogenes.

**Figure S13.**
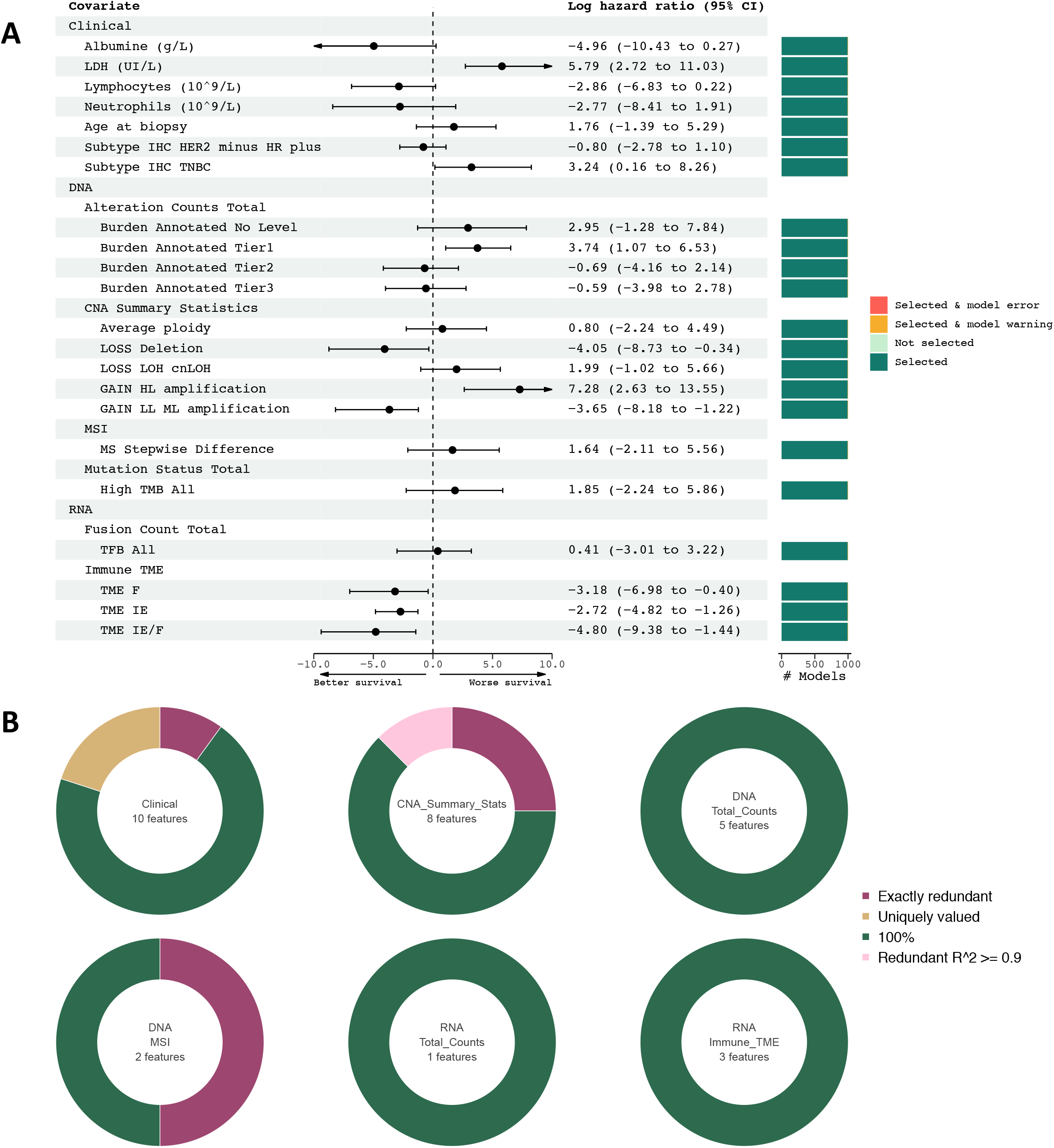
Cox models M7 coefficients in META-PRISM WES & RNAseq, BRCA tumors. **A.** Coefficients and 95% confidence intervals estimates from 1,000 Cox models (see Supplementary Methods) using covariates from M7 model on BRCA tumors from META-PRISM WES & RNAseq subcohort. **B.** Number of features from each category that were removed or selected during the modeling preprocessing and fitting steps. Green colors indicate the selection frequency of selected features, while non-green colors indicate reasons for removal.

## Supplementary Methods Figures

**Supplementary Methods Fig. 1.**
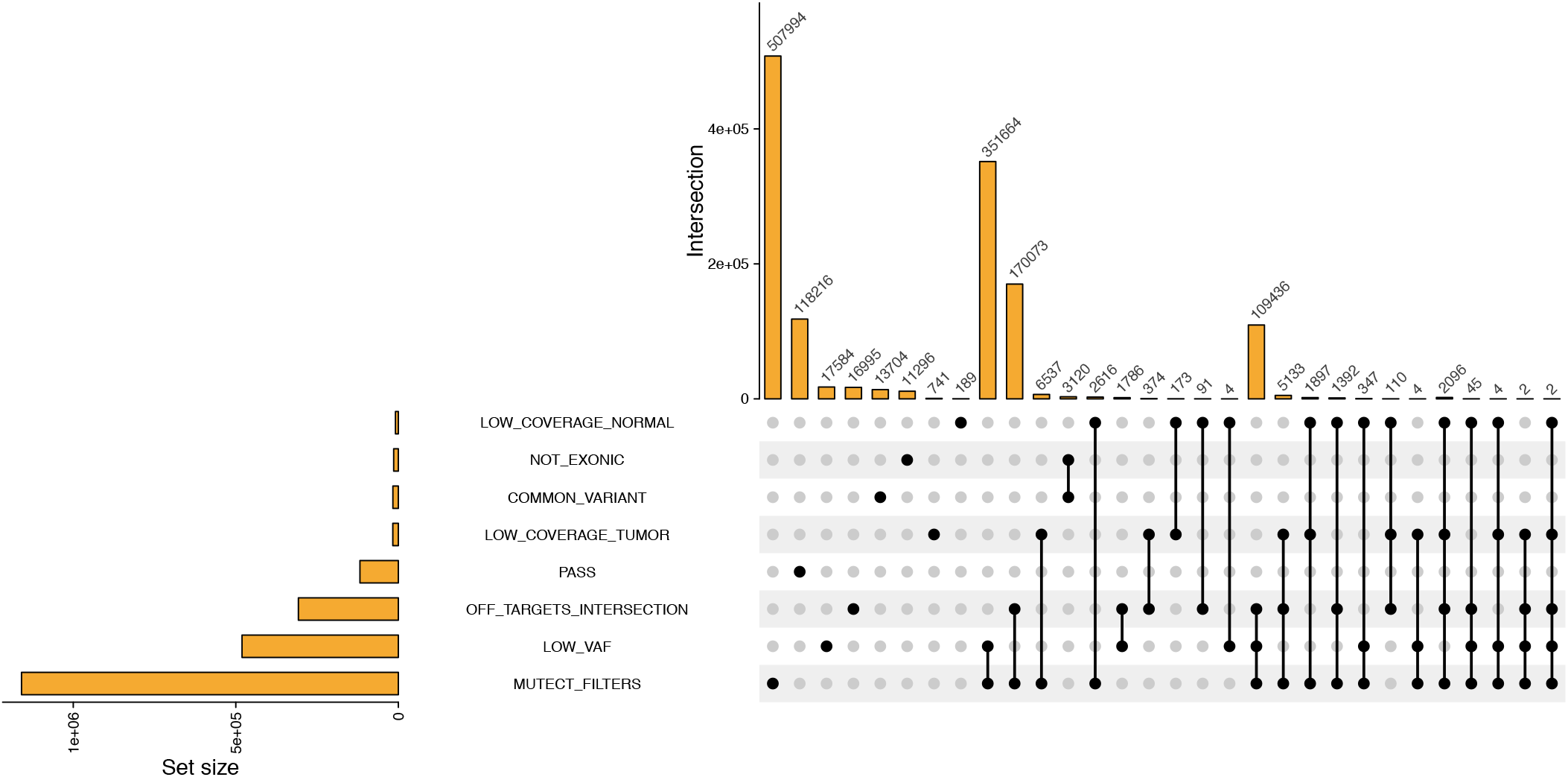
Mutation filtering on META-PRISM WES samples.

**Supplementary Methods Fig. 2.**
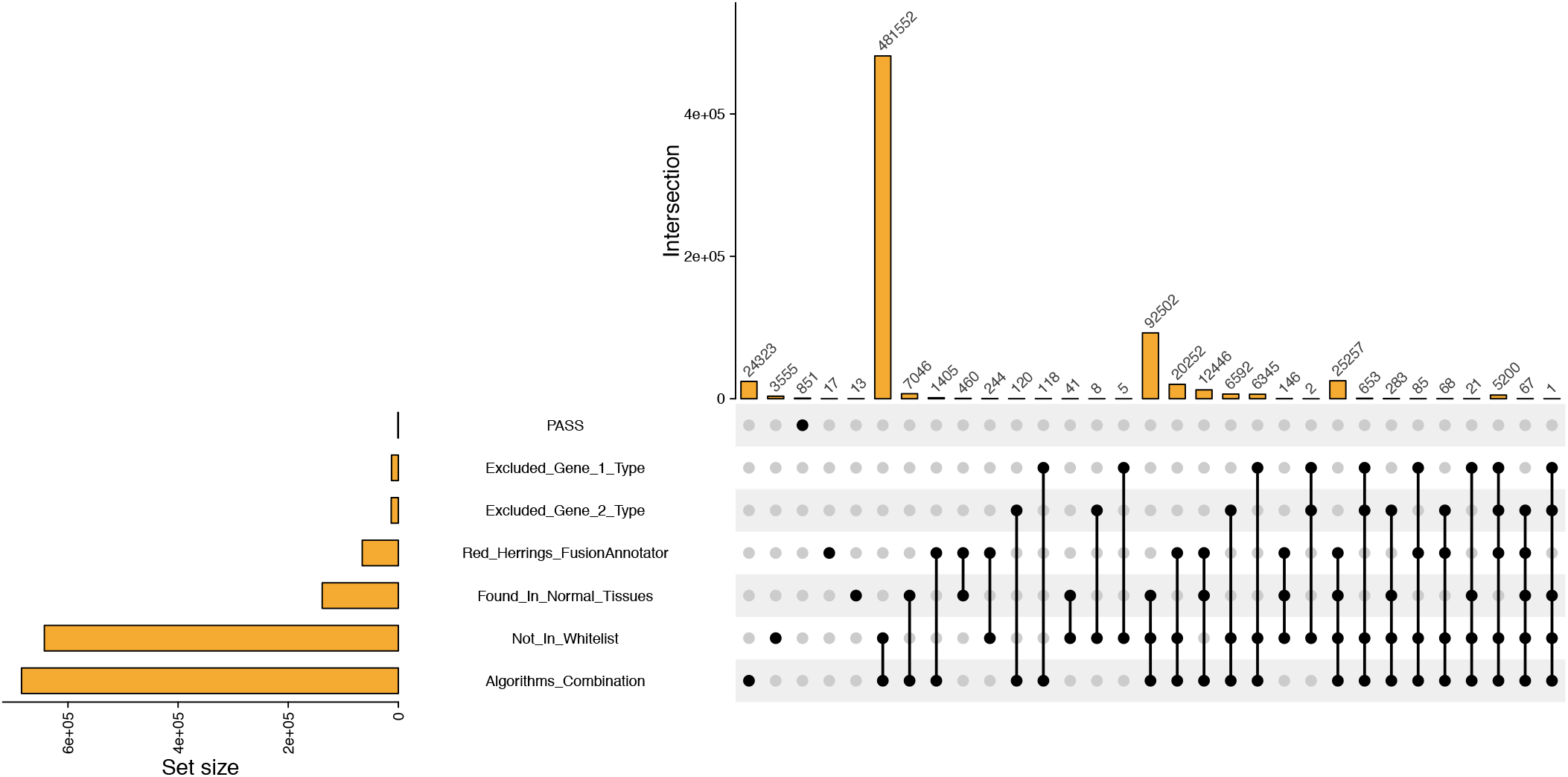
Gene fusion filtering on META-PRISM RNAseq samples.

**Supplementary Methods Fig. 3.**
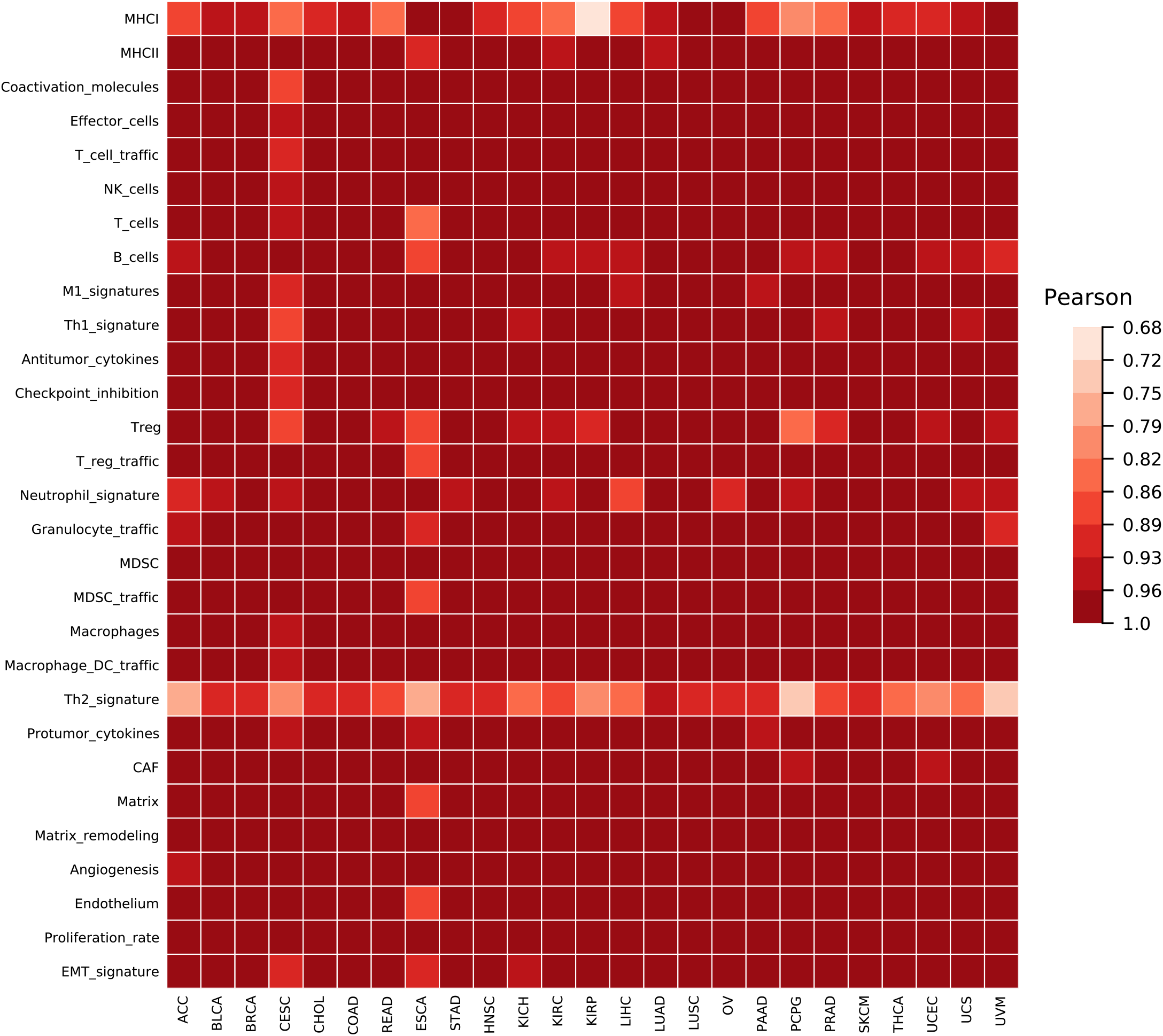
Signatures scores concordance between TCGA Bagaev 2021 data and TCGA META-PRISM data.

**Supplementary Methods Fig. 4.**
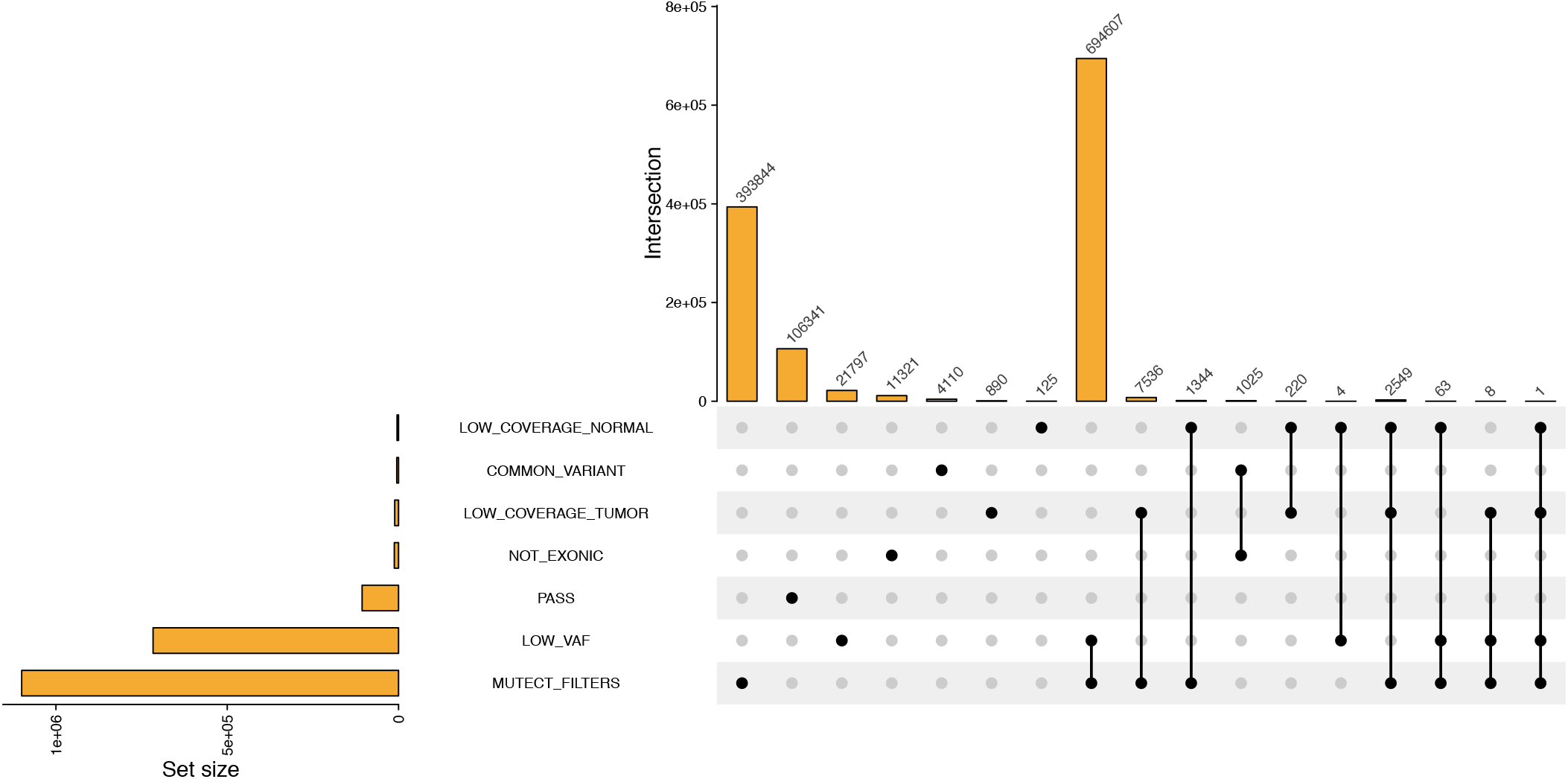
Mutation filtering on MET500 WES samples.

**Supplementary Methods Fig. 5.**
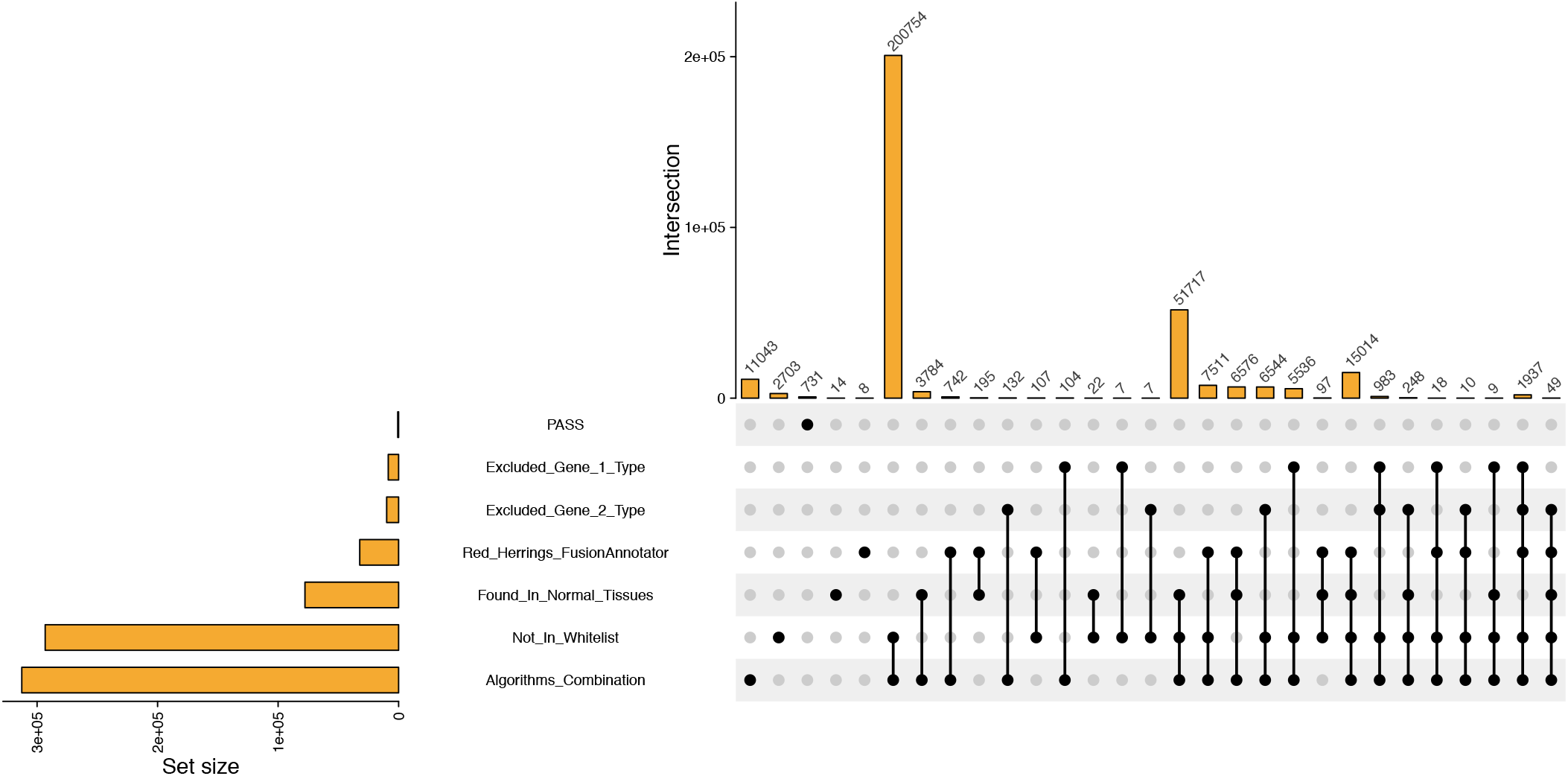
Gene fusion filtering on MET500 RNAseq samples.

**Supplementary Methods Fig. 6.**
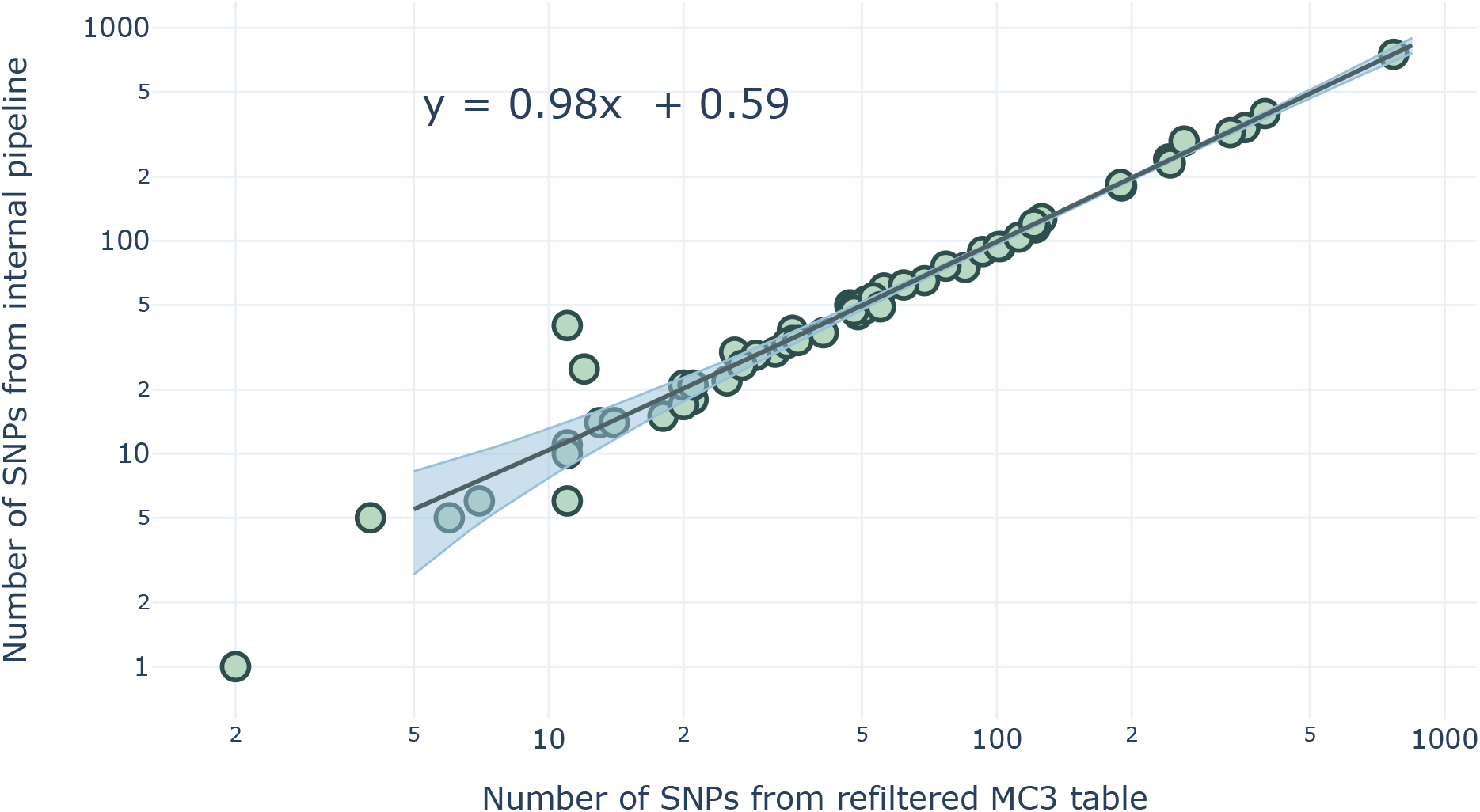
SNP-calling pipeline alignment.

**Supplementary Methods Fig. 7.**
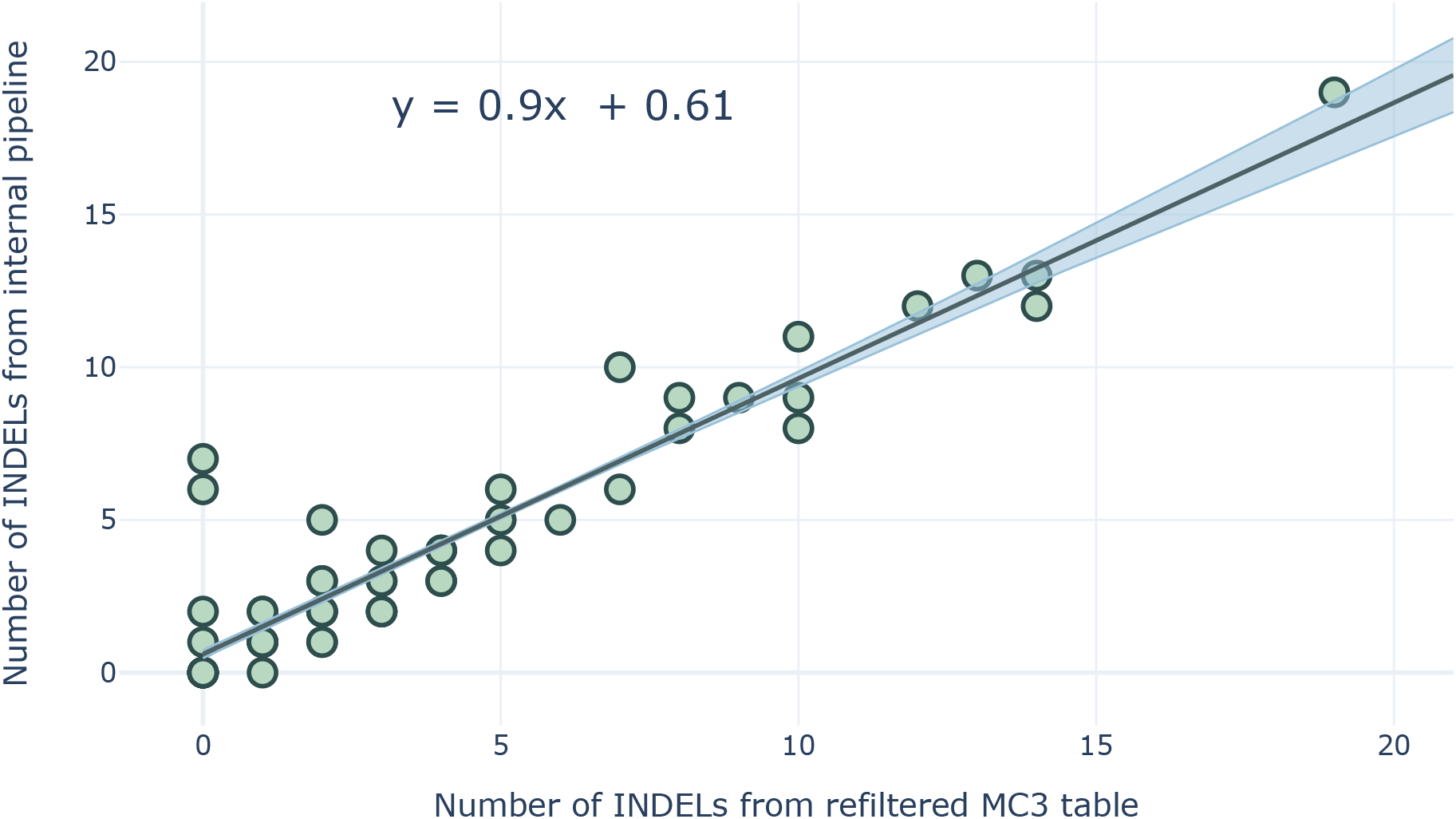
INDEL-calling pipeline alignment.

**Supplementary Methods Fig. 8.**
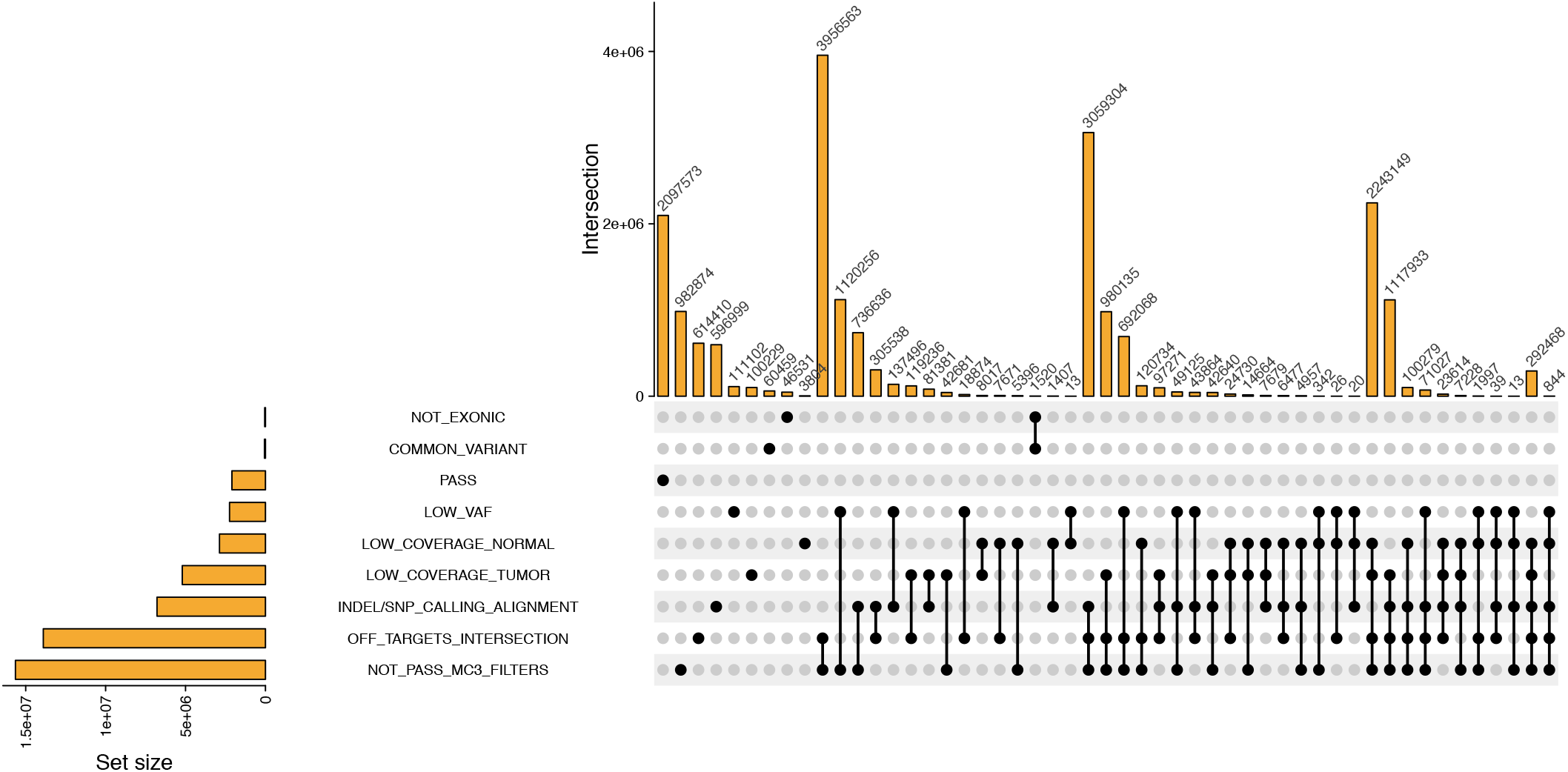
Mutation filtering on TCGA WES samples.

**Supplementary Methods Fig. 9.**
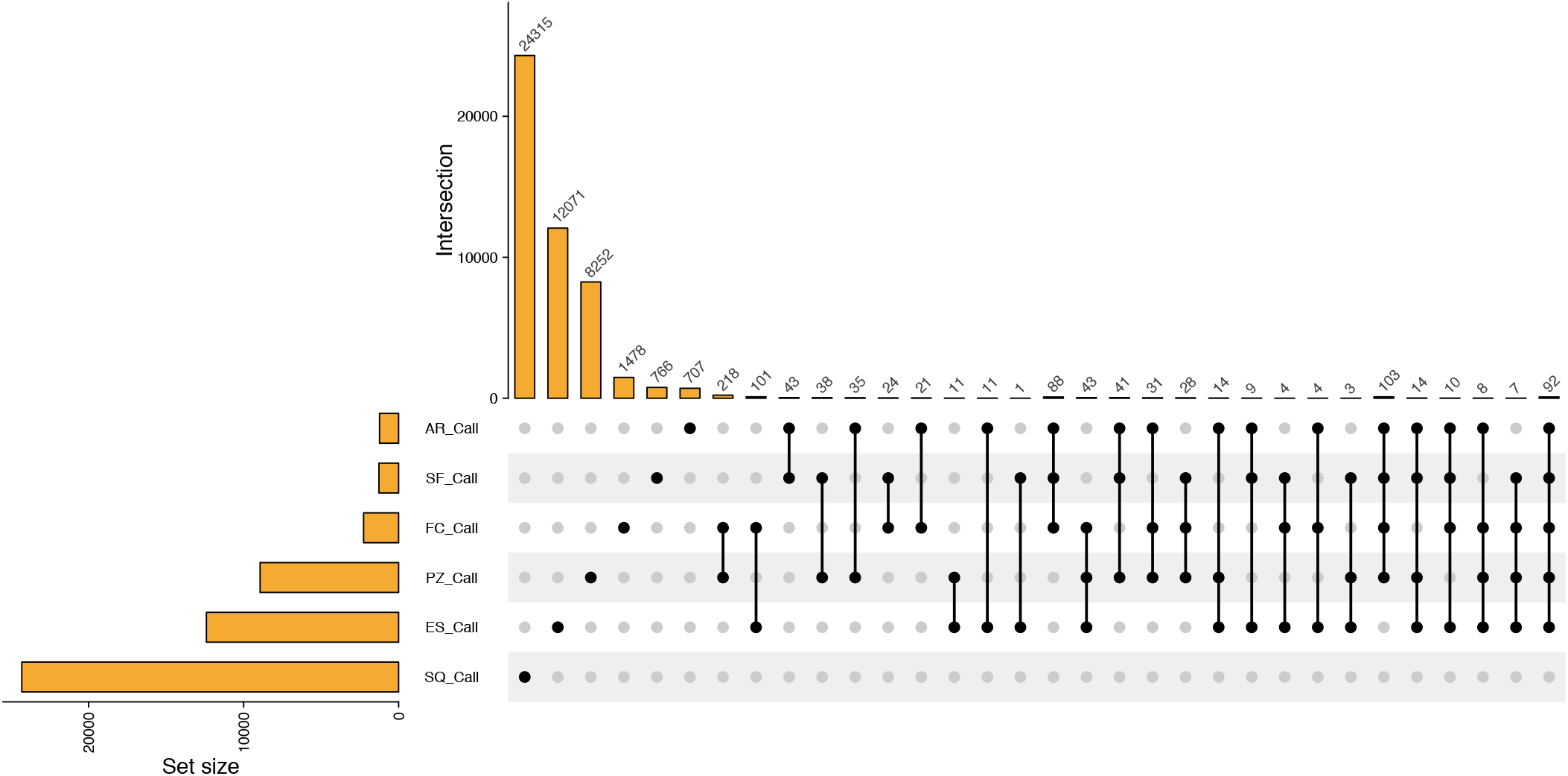
Gene fusion calls by six candidate algorithms on 69 TCGA RNAseq samples.

**Supplementary Methods Fig. 10.**
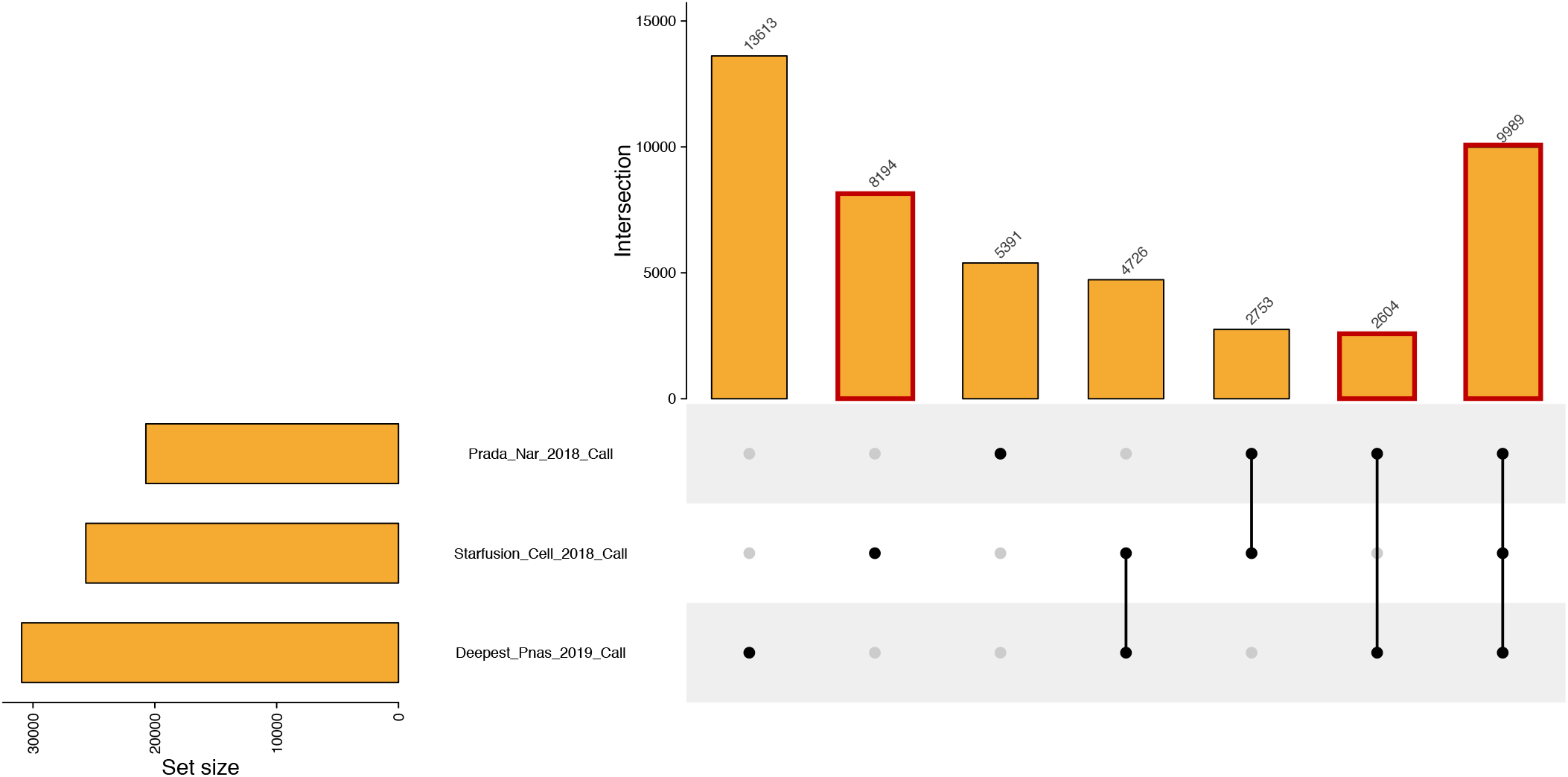
Gene fusion filtering on TCGA RNAseq samples.

## Footnotes

1 https://www.bioinformatics.babraham.ac.uk/projects/fastqc/

2 http://broadinstitute.github.io/picard/

3 https://github.com/FelixKrueger/TrimGalore

4 https://gatk.broadinstitute.org/hc/en-us/articles/360035890471-Hard-filtering-germline-short-variantsh

5 https://gatk.broadinstitute.org/hc/en-us/articles/360035890631-Panel-of-Normals-PON-

6 https://ascopubs.org/doi/suppl/10.1200/PO.17.00073/suppl_file/ds_PO.17.00073-2.xlsx

7 https://cancer.sanger.ac.uk/signatures/sbs/

8 https://fr.mathworks.com/matlabcentral/fileexchange/38724-sigprofiler

9 https://github.com/AlexandrovLab/SigProfilerExtractor

10 https://bitbucket.org/bbglab/sigprofilerjulia

11 https://github.com/nf-core/rnafusion

12 https://github.com/oncokb/oncokb-annotator

13 https://civicdb.org/releases

14 https://github.com/BostonGene/MFP/tree/master/signatures

15 https://github.com/BostonGene/MFP/tree/master/Cohorts/Pan_TCGA

16 https://github.com/zhengh42/RNASeq_pipeline

17 https://github.com/FusionAnnotator/FusionAnnotator

18 https://github.com/FusionAnnotator/CTAT_HumanFusionLib/wiki

19 https://cancer.sanger.ac.uk/cosmic/fusion

20 https://genetica.unav.edu/TICdb/

21 https://gnomad.broadinstitute.org/downloads#exac-variants

